# Predicting thoracic aortic dissection in a diverse biobank using a polygenic risk score

**DOI:** 10.1101/2024.09.01.24312895

**Authors:** John DePaolo, Siavash Zamirpour, Sarah Abramowitz, Gina Biagetti, Renae Judy, Cameron Beeche, Jeffrey Duda, James Gee, Walter R. Witschey, Julio A. Chirinos, Nicholas J. Goel, Nimesh Desai, Wilson Y. Szeto, Dongchuan Guo, Dianna M. Milewicz, Michael G. Levin, James P. Pirruccello, Scott M. Damrauer

## Abstract

**Background:** Thoracic aortic dissection is a life-threatening condition that often occurs in the presence of aortic dilation. Despite a known association between ascending aortic diameter (AscAoD) and dissection risk, predicting dissection risk remains challenging.

**Objectives:** Determine whether common variant genetics can be used to improve identification of individuals most at risk for dissection.

**Methods:** A genome wide association study (GWAS)-by-subtraction was performed to characterize the diameter-independent genetics of thoracic aortic dissection by subtracting a GWAS of aortic diameter (AoD) from a GWAS of thoracic aortic aneurysm and dissection (TAAD). A polygenic risk score (PRS) was calculated using the PRS-Continuous Shrinkage statistical package and evaluated for its ability to predict aortic dissection. The primary analytic cohort was Penn Medicine Biobank (PMBB) which comprises volunteers consenting to linkage of health records with biospecimens, including DNA which has undergone genome-wide genotyping; additional analyses were performed in the National Institutes of Health All of Us (AoU) cohort.

**Results:** We identified 43 significant genetic risk loci in our GWAS-by-subtraction and derived a “Dissection-PRS.” In the PMBB, the Dissection-PRS associated with an increased risk of prevalent dissection (odds ratio [OR]=2.13 per 1 standard deviation [sd] increase in Dissection-PRS, 95% confidence interval [CI] 1.91 to 2.39, *P*<0.001), These results were consistent when excluding individuals with pathogenic or likely pathogenic variants in established aortopathy genes. When adjusting for clinical risk factors including ascending aortic diameter, the association of the Dissection-PRS with prevalent dissection attenuated but remained significant (OR=1.62 per 1 sd increase in PRS, 95% CI 1.36 to 1.94, *P*<0.001). The addition of the PRS to a model containing age, sex, clinical risk factors, and ascending aortic diameter substantially improved model discrimination (base model area under the receiver operator characteristic curve [AUROC]=0.676, 95% CI 0.651 to 0.702; with addition of PRS AUROC=0.723, 95% CI 0.702 to 0.744). Analysis in AoU demonstrated similar findings.

**Conclusions:** A common-variant PRS can predict aortic dissection in a diverse population.

## Introduction

Thoracic aortic dissection is characterized by tearing of the intimal layer of the thoracic aorta, more commonly in the ascending portion (Stanford Type A dissection), leading to formation of a false lumen resulting in antegrade and/or retrograde malperfusion.^1^ Dissection has an incidence rate of approximately 3-10 per 100,000 individuals annually,^2–5^ and although rare, acute dissection can have devastating health consequences with 30-day mortality rates ranging from 17% to more than 50% when considering out-of-hospital deaths.^6–11^ Currently, individuals with dilated or aneurysmal thoracic aortas are treated with aggressive blood pressure management and serial imaging with elective surgical repair at ascending thoracic aortic diameters (AscAoD) ≥ 5-5.5 cm.^12^ However, it has become increasingly evident by the prevalence of dissection among individuals with AscAoD < 5.5 cm that this size cutoff alone is insufficient to guide clinical decision making.^13^

Dissection, along with ascending and descending thoracic aortic aneurysms, are included in the group of related conditions referred to as thoracic aortic aneurysm and dissection (TAAD). Rare pathogenic (P) or likely pathogenic (LP) variants in several Mendelian aortopathy genes associated with hereditary TAAD (HTAAD) are causally linked to familial and/or syndromic disease.^14,15^ Notably, due to overwhelming genetic risk, the size threshold for surgical intervention to prevent dissection is lower among individuals with P/LP rare variants in a subset of aortopathy genes associated with familial or syndromic disease.^12^ Despite TAAD being historically thought of as a Mendelian disease, approximately 80% of cases of TAAD occur among individuals without evidence of heritable disease.^16^ Recent genome-wide association studies (GWAS) have identified dozens of common single nucleotide variants (SNVs) associated with increased thoracic aortic diameter (AoD),^17,18^ and TAAD.^19^ To characterize common variant risk, polygenic risk scores (PRS) have been constructed from these GWAS data using different methodologies and have shown consistent association with prevalent and incident thoracic aortic disease in different biobank cohorts.^17–19^

In this study, we utilized Genomic-Structural Equation Modeling (Genomic-SEM) to perform a GWAS-by-subtraction to determine the diameter independent common genetic variants associated with dissection. This procedure revealed 43 genome-wide significant genetic risk loci associated with diameter-independent thoracic aortic disease. From this GWAS we generated a “Dissection-PRS” that better associates with prevalent dissection compared to existing PRSs. Using this novel Dissection-PRS, we were able to predict individuals who are most at risk of dissection independent of other clinical factors, including aortic size.

## Methods

### Study Population

#### PMBB

The Penn Medicine BioBank is a genomic and precision medicine cohort comprising participants who receive care in the Penn Medicine health system and who consent to linkage of electronic health records with biospecimens, including 43,731 with DNA which has undergone whole exome sequencing and 43,623 which has undergone genome-wide genotyping (pmbb.med.upenn.edu).^20^ Among these individuals, 7,947 have had at least one transthoracic echocardiography (TTE) study with a recorded measurement of AscAoD or a computed tomography (CT) scan with an interpretable aortic diameter. Details of the genetic acquisition, quality control, and assignment of population groups are included in the **<u>Supplemental Methods</u>**.

#### All of Us

The *All of Us* (AOU) research program is a multi-site, prospective cohort study in the United States.^21^ The enrollment process included a physical examination and biospecimen collection, with follow-up based on electronic health record (EHR) records and surveys. All participants provided written, informed consent. Analysis of AOU was considered exempt by the UCSF IRB (#22-37715). Further information regarding genetic information and data analysis of the *AOU* cohort are provided in the **<u>Supplemental Methods</u>**.

### Primary outcome in the PMBB

The primary outcome of our study was thoracic aortic dissection. Dissection was defined either as ≥2 outpatient or ≥1 inpatient encounters coded with *International Classification of Diseases, 10^th^ Revision* (ICD10) diagnosis codes I71.01 or I71.03, or *International Classification of Diseases, Ninth Revision* (ICD9) codes 441.01 or 441.03.

### Clinical Covariate Selection

Previous studies have identified several clinical variables as risk factors for ascending thoracic aortic dilation and/or dissection.^22–25^ From these studies, we selected those covariates that are most easily and universally assessed, referred to as clinical risk factors (CRF), including height, weight, body mass index (BMI), body surface area (BSA), systolic blood pressure (SBP), diastolic blood pressure (DBP), and heart rate (HR). For each individual, the mean value of each covariate taken over time was used in the analysis. In a subset of the analyses, we also used AscAoD as measured by either TTE, or computed tomography (CT). Participant TTE measurements were derived from clinical echocardiography reports recorded in EHR. CT measurements were derived using automated segmentation of the ascending aorta as previously described.^26^

### Genome Wide Association Study

GWAS-by-subtraction was performed using Genomic-Structural Equation Modeling (SEM), as previously described.^27^ Genomic-SEM is a flexible modeling framework that allows a user to create systems of equations to model relationships between the genetic components of observed traits and related latent traits. Because TAAD comprises both aortic aneurysm (i.e. large aortic diameter) and dissection, we posited that subtracting the genetic contribution of aortic size from TAAD would isolate the genetics of aortic dissection. Using publicly available summary statistics from GWAS of TAAD,^19^ ascending aortic diameter (AscAoD),^17^ and descending aortic diameter (DesAoD)^17^ (summary information in **Supplemental Table 1**) we employed the genomic-SEM framework to perform GWAS-by-subtraction based on a Cholesky decomposition model that subtracts out the genetic covariance contributed by multiple related traits resulting in a novel GWAS of latent traits.^28^ This allowed us to identify variants uniquely associated with TAAD and independent of AscAoD or DesAoD.^29^

The objective of this method was to estimate the association with TAAD independent of that variant’s association with AscAoD and DesAoD for each genetic variant identified in the TAAD parent GWAS. To do this, the Cholesky decomposition model (**Supplemental Figure 1**) regressed each genetic variant on three latent factors: one defined as all aortic diameter plus TAAD (AscAoD + DesAoD + TAAD), a second defined as descending aortic diameter + TAAD – ascending aortic diameter [(DesAoD + TAAD) – AscAoD], and a third defined as diameter-independent TAAD (TAAD – (AscAoD + DesAoD)), which we posit represents dissection alone. In creating such a model, we assume that all genetic variants that affect thoracic aortic diameter also affect TAAD. The analysis allows for three paths by which a genetic variant can affect TAAD: first, mediated through aortic diameter + the TAAD latent trait (Latent Trait 1); second, mediated through just the DesAoD + TAAD) – AscAoD latent trait (Latent Trait 2), and finally mediated through the diameter-independent TAAD latent trait (Latent Trait 3), our latent trait of focus. Each genetic variant was regressed on each latent trait to determine the effect size specific to the latent trait of interest. Independent lead loci for the diameter-independent TAAD latent trait were then selected using *P*<5×10^−8^, a radius of 250kb, and an LD threshold of *r*^2^<0.001.

### Polygenic risk score creation

PRS-CSx-auto^30^ was used to construct a PRS for each of the following traits: 1) AscAoD from a GWAS performed among 38,694 UKB participants;^17^ 2) TAAD from a GWAS among a diverse population of 461,669 MVP participants (8,626 cases, 453,043 disease-free controls);^19^ 3) thoracic aortic dissection (“Dissection-PRS”) using our GWAS-by-subtraction summary statistics described above. These are described further in the **<u>Supplemental Methods</u>**.

To apply each PRS in the PMBB, individual participant scores were calculated using pgsc_calc, a pipeline developed by the PGS Catalog that computes individual scores by combining imputed genotypes with PRS weights.^31^ As allele frequency differences across diverse populations can influence PRS distribution and limit accuracy, pgsc_calc allows a genetic principal component (PC)-based method to normalize scores across populations by adjusting the mean and variance relative to the 1000 Genomes Project and Human Genomes Diversity Project (HGDP).^32^ In this approach, a PC space is created using reference populations and a PRS is modeled as a linear function of the PCs. Score residuals are calculated as the difference between the observed and predicted PRS, and then divided by the standard deviation of the residuals. This ensures that the adjusted PRS values have a mean of 0 when considering the influence of population diversity. To adjust for the variations in PRS range, the variance of the residuals is modeled as a function of the PCs, and then is used to normalize the residual PRS. The variance of the final PRS values distribution is therefore approximately 1 across all populations.

### Prediction model creation

Using the selected clinical covariates and the Dissection-PRS we created, we derived the following six regression models to predict dissection in the PMBB:

1. Age + Sex + genetic PC1-PC5 (Age + Sex model)
2. Age + Sex + genetic PC1-PC5 + Clinical Risk Factors (Age + Sex + CRF model)
3. Age + Sex + genetic PC1-PC5 + Clinical Research Factors + Dissection-PRS (Age + Sex + CRF + PRS model)
4. Age + Sex + genetic PC1-PC5 + Clinical Research Factors + AscAoD (Age + Sex + CRF + AscAoD model)
5. Age + Sex + genetic PC1-PC5 + Clinical Research Factors + AscAoD + Dissection-PRS (Age + Sex + CRF + AscAoD + PRS model)

Prediction model cross-validation and statistical testing is described in the

## Supplemental Methods

### Statistical Analysis

Multivariable logistic regression was employed to evaluate the risk of prevalent dissection associated with the Dissection-PRS adjusting for age, sex, and genetic PCs 1-5. Incident dissection was modeled with Cox proportional hazards model with the R survival package,^33^ and including the same covariates as above. Where described, other covariates were added to specific analyses. All statistical analyses were performed in R version 4.3.2 (R Foundation for Statistical Computing, Vienna, Austria).

## Results

### Study population for PRS Analysis

The primary analysis was performed in the Penn Medicine BioBank (PMBB). There were 43,249 participants in the analytic cohort (**Table 1**), of which 262 individuals had a diagnosis of aortic dissection. Of those with a dissection 86 (33%) were female, 76 (29%) were genetically similar to the 1000 Genomes Project (1000G) African reference population (AFR), and the median age at analysis of individuals was 63.4 years (interquartile range [IQR]: 53.9-71.1 years). Among those without a diagnosis of dissection, 21,576 (50%) were female, 11,035 (26%) were genetically similar to the 1000G AFR reference population, and the median age at analysis was 57.2 years (IQR: 42.5-67.2 years). Overall, individuals with a diagnosis of dissection had greater median height (175.3 cm compared to 170.2 cm), weight (86.2 kg compared to 83.0 kg), body mass index (BMI) [28.5 compared to 28.2], body surface area (BSA) [2.02 m^2^ compared to 1.96 m^2^], and systolic blood pressure (127.5 mmHg compared to 126.3 mmHg). Concurrently, individuals with a diagnosis of dissection had a lower median diastolic blood pressure (70.7 mmHg compared to 74.0 mmHg) and heart rate (73.9 beats per minute [bpm] compared to 77.2 bpm).

**Table 1:** Clinical characteristics of Penn Medicine Biobank individuals with and without thoracic aortic dissection.

| **Penn Medicine Biobank Participant Characteristics** |  |  |
| --- | --- | --- |
|  | Non-Dissection | Dissection |
| Cohort Size, N | 42987 | 262 |
| Sex |  |  |
| Female (%) | 21576 (50%) | 86 (33%) |
| Male (%) | 21411 (50%) | 176 (67%) |
| Population |  |  |
| AFR (%) | 11035 (26%) | 76 (29%) |
| EUR (%) | 29770 (69%) | 177 (68%) |
| Other (%) | 2182 (5.1%) | 9 (3%) |
| Age in years, median (IQR) | 57.2 (42.5-67.2) | 63.4 (53.9-71.1) |
| Height in cm, median (IQR) | 170.2 (162.6-177.8) | 175.3 (166.8-182.9) |
| Weight in kg, median (IQR) | 83.0 (70.1-97.9) | 86.2 (75.1-98.7) |
| Body mass index, median (IQR) | 28.2 (24.7-33.0) | 28.5 (25.5-31.7) |
| Body surface area in meters squared, median (IQR) | 1.96 (1.78-2.13) | 2.02 (1.84-2.20) |
| Blood pressure |  |  |
| Systolic, median (IQR) | 126.3 (117.9-134.8) | 127.5 (120.6-135.3) |
| Diastolic, median (IQR) | 74.0 (69.0-79.0) | 70.7 (65.6-76.4) |
| Heart rate, median (IQR) | 77.2 (70.3-84.5) | 73.9 (65.7-82.1) |

Replication analyses were performed in the AOU cohort. There were 245,149 participants in this cohort, 133,444 of whom were genetically similar to the 1000G European reference population (EUR) and 56,870 genetically similar to the 1000G AFR reference population. Of the 205 individuals with diagnosis of dissection, 114 were genetically similar to the 1000G EUR reference population and 65 were genetically similar to the 1000G AFR reference population.

### Genome-wide association study-by-subtraction

Previously, we published a GWAS of TAAD performed among a diverse population in the Million Veterans Program (MVP) that identified 21 genetic risk loci associated with TAAD.^19^ Similarly, we published a GWAS of AscAoD and DesAoD in the UK Biobank that identified 82 (AscAoD) and 47 (DesAoD) independent loci, respectively.^17^ In an attempt to better characterize the pathologic variants associated with dissection but independent from thoracic AoD, we performed a GWAS-by-subtraction^27^ using the Genomic-SEM framework^29^ to model the genetics of a novel latent trait referred to as “diameter-independent dissection”. Using this methodology, we identified 43 independent risk loci that reached genome-wide significance and were primarily associated with thoracic aortic dissection independently of their effects on aortic diameter, (**Figure 1, Supplemental Tables 2-3, Supplemental Figures 2-5**) described further in the **<u>Supplemental Results</u>**.

**Figure 1:**
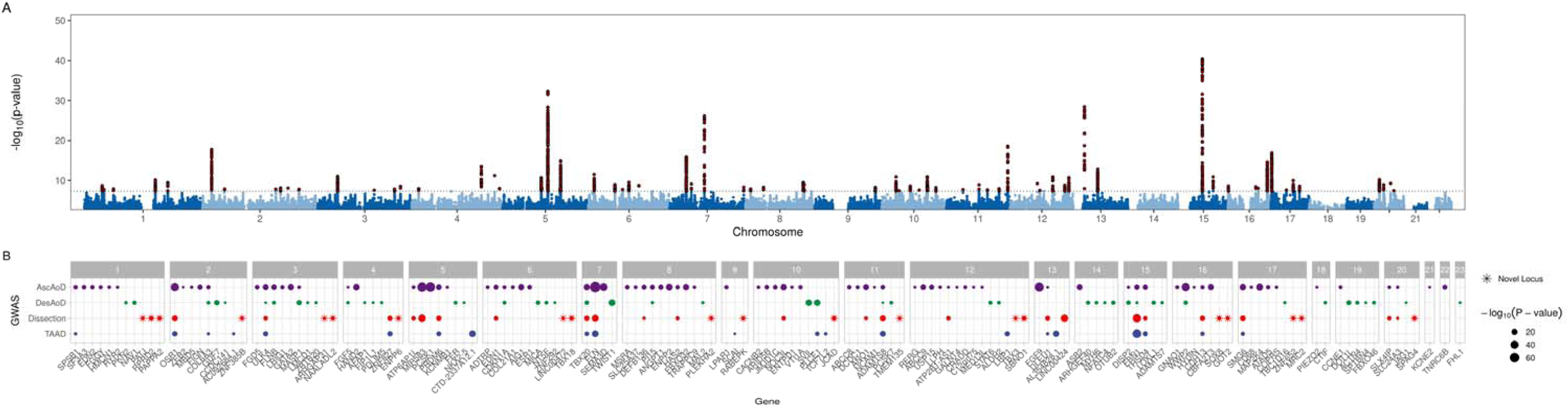
Genome-wide association study results. **A)** Manhattan plot of multi-trait genome-wide association study-by-subtraction results subtracting AscAoD and DesAoD from TAAD. Each point represents a genetic variant. Genome-wide significant (*P*<5×10^−8^) loci are represented by peaks of red points that associate with diameter-independent dissection. The X-axis represents genomic position by chromosome and the y-axis represents the strength of association by −log_10_(*P*-value). **B)** Candidate genes, grouped by chromosome, were assigned to each genome-wide significant (*P*<5×10^−8^) locus in the diameter-independent “dissection” (red) and compared to the previously published TAAD GWAS (blue), ascending aortic diameter GWAS (purple), and descending aortic diameter GWAS (green) with previously unreported candidate genes denoted by stars. The size of each point corresponds to the strength of the association represented by −log_10_(*P*-value).

### Polygenic risk score association with prevalent thoracic aortic dissection

To determine whether we could use genetic information to better predict dissection, we used our GWAS-by-subtraction summary statistics to create a diameter-independent dissection PRS, referred to as the “Dissection-PRS.” Individuals in the PMBB, an independent cohort of patients separate from the parent GWAS included in the GWAS-by-subtraction, were then used to assess the PRS association with prevalent thoracic aortic dissection (**Central Illustration**).

**Central Illustration.**
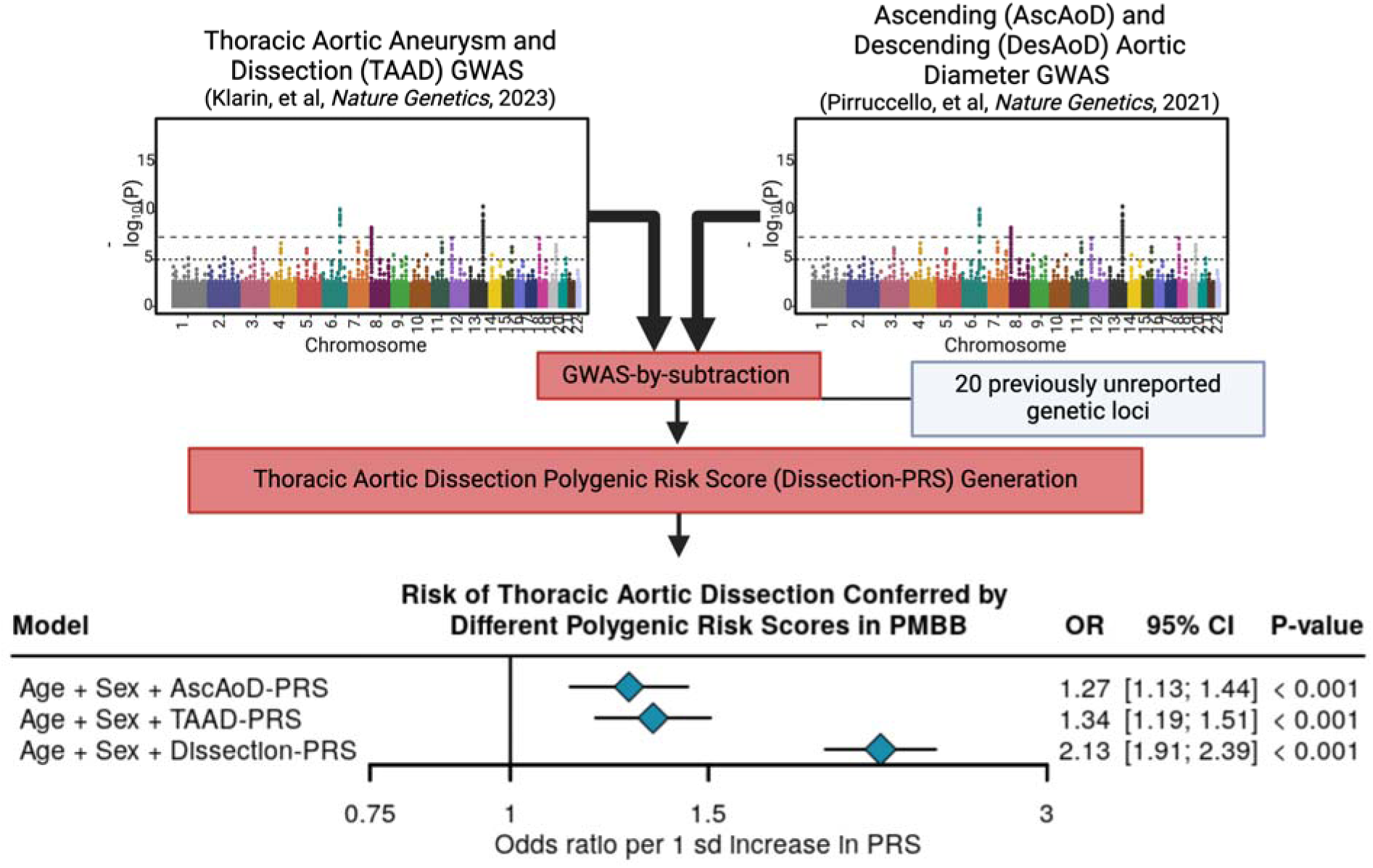
GWAS-by-subtraction, polygenic risk score creation, and association of different PRS with prevalent thoracic aortic dissection in the Penn Medicine Biobank. Analytic approach to the GWAS-by-subtraction of diameter-independent dissection followed by polygenic risk score creation and multivariable logistic regression analysis of the Dissection-PRS, TAAD-PRS, and AscAoD-PRS among all individuals in the PMBB to determine respective association with prevalent thoracic aortic dissection, adjusting for age, sex, and the first five genetic principal components. AscAoD-PRS = Ascending aortic diameter polygenic risk score; CI = confidence interval; OR = Odds ratio; sd = standard deviation; TAAD-PRS = thoracic aortic aneurysm and dissection polygenic risk score; Dissection-PRS = thoracic aortic dissection polygenic risk score.

Compared to individuals without a diagnosis of dissection (median PRS = 0.03, 95% CI = 0.01 to 0.04), individuals with a diagnosis of dissection had a significantly higher median PRS (0.88, 95% CI 0.68 to 1.06, P=1.07×10^−30^) [**Supplemental Figure 6**]. To further characterize the strength of the Dissection-PRS, we compared its association with prevalent dissection in the PMBB to the association of either a TAAD-PRS or AscAoD-PRS. Compared to the AscAoD-PRS (OR=1.27 per 1 sd increase in PRS, 95% CI 1.13 to 1.44, *P*<0.001) and the TAAD-PRS (OR=1.34 per 1 sd increase in PRS, 95% CI 1.19 to 1.51, *P*<0.001), the Dissection-PRS had a profoundly stronger association with prevalent dissection (OR=2.13 per 1 sd increase in PRS, 95% CI 1.91 to 2.39, *P*<0.001) [**Central Illustration**]. This was consistent when individuals were stratified by genetically similar population group (EUR: OR=1.93 per 1 sd increase in PRS, 95% CI 1.67 to 2.23, *P*<0.001; AFR: OR=2.61 per 1 sd increase in PRS, 95% CI 2.14 to 3.18, *P*<0.001; Meta: OR=2.14 per 1 sd increase in PRS, 95% CI 1.91 to 2.41, *P*<0.001) [**Supplemental Figure 7**], and when individuals carrying pathogenic/likely pathogenic (P/LP) HTAAD gene variants were excluded from the analysis (OR=2.15 per 1 sd increase in PRS, 95% CI 1.91 to 2.42, *P*<0.001) [**Supplemental Figure 8**].

To validate our findings, analyses were replicated in the AOU cohort with genetic data (N=245,149, including 132 participants with prevalent dissection). In this cohort, the Dissection-PRS remained robustly associated with prevalent dissection (OR=1.48, 95% CI 1.27 to 1.74, *P*<0.001). When stratified by population group, similar results were observed in the EUR population that included 73 cases of prevalent dissection (OR=1.89, 95% CI 1.52 to 2.34, *P*<0.001), however the association was not statistically significant among AFR population that included 38 cases of dissection (OR=1.09, 95% CI 0.82 to 1.45, *P*=0.57). We conclude that the Dissection-PRS strongly associates with prevalent dissection in PMBB across diverse populations, has been externally validated in AOU though with an attenuated effect in the AFR population, and is independent of P/LP HTAAD gene variants.

### Effect of Dissection-PRS when adjusting for known risk factors

To determine if the Dissection-PRS was independently associated with prevalent dissection independent of other risk factors, we performed multivariable logistic regression analysis adjusting for CRF including age, sex, height, weight, BMI, BSA, SBP, DBP, and HR. When we adjusted for these variables, the effect of the Dissection-PRS remained consistent (OR=2.29 per 1 sd increase in PRS, 95% CI 1.97 to 2.65, *P*<0.001) [**Figure 3**].

**Figure 3:**
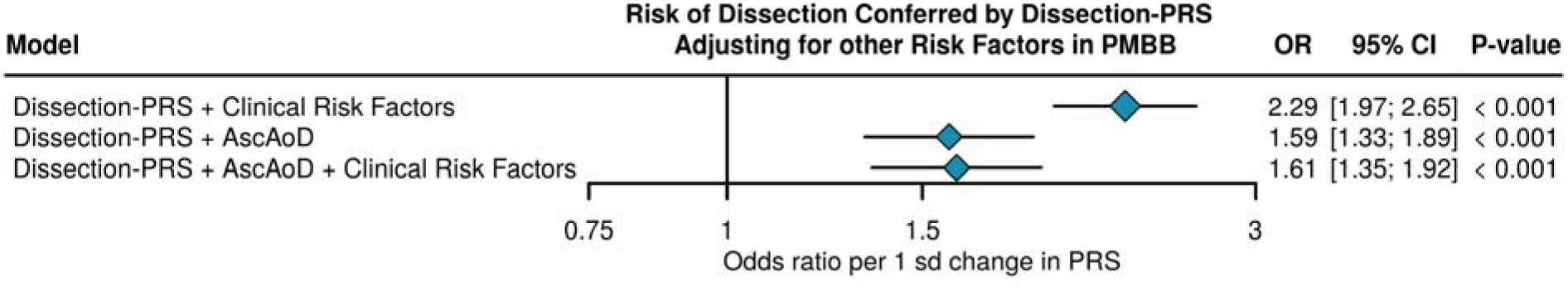
Association of Dissection-PRS with prevalent dissection in multivariable logistic regression analysis adjusting for different clinical risk factors. Multivariable logistic regression analysis of the effect of Dissection-PRS on prevalent dissection adjusting for clinical risk factors, measured AscAoD, or clinical risk factors and measured AscAoD. Each logistic regression analysis was also adjusted for age, sex, and the first five genetic principal components. AscAoD = Ascending aortic diameter measured by TTE or CT; CI = confidence interval; OR = Odds ratio; sd = standard deviation; Dissection-PRS = thoracic aortic dissection polygenic risk score.

As AscAoD is the most critical clinical factor in determining when to offer surgical intervention,^12,13^ we repeated our multivariable logistic regression to also adjust for AscAoD among 7,974 individuals with AscAoD measured by TTE (7,414) or CT (560) [**Supplemental Table 4**]. When adjusting for AscAoD alone (OR=1.59 per 1 sd increase in PRS, 95% CI 1.33 to 1.92, *P*<0.001), or AscAoD and CRF (OR=1.61 per 1 sd increase in PRS, 95% CI 1.35 to 1.92, *P*<0.001), the effect estimate of the Dissection-PRS was attenuated but remained statistically significant [**Figure 3**]. These results suggest that the risk of dissection associated with the Dissection-PRS is completely independent of CRFs and largely independent of measured AscAoD.

### PRS effect on predictive model calibration

To determine if the Dissection-PRS could improve identification of individuals at increased risk of dissection, we integrated the Dissection-PRS in prediction models and compared model performance with and without the PRS. To determine the effect of the Dissection-PRS on model calibration, we assessed differences in model log-loss. Log-loss is a measure of how close a predictive probability is to the corresponding actual value with a lower log-loss equating to model improvement; a lower log-loss connotes improved model calibration. A prediction model constructed with just age, sex, and the first 5 genetic PCs (Age + Sex) had a log-loss of 0.075 (95% CI 0.070 to 0.080) [**Supplemental Figure 9**]. Compared to an Age + Sex + CRF model that integrated weight, BMI, height, BSA, SBP, DBP, and HR (log-loss=0.074, 95% CI 0.069 to 0.079), an Age + Sex + CRF + Dissection-PRS had an improved model log-loss (log-loss=0.071, 95% CI 0.066 to 0.076). Similarly, compared to an Age + Sex + CRF + AscAoD model (log-loss=0.071, 95% CI 0.066 to 0.076), an Age + Sex + CRF + AscAoD + Dissection-PRS had an improved log-loss (log-loss=0.069, 95% CI 0.064 to 0.074). To determine if the addition of the Dissection-PRS meaningfully improved model calibration, we employed cross-model Bayesian ANOVA analysis of log-loss. The addition of the Dissection-PRS to the Age + Sex + CRF model (mean log-loss difference = −0.002, 95% credible interval −0.004 to 0.000; >95% probability of a practical improvement in log-loss) and the Age + Sex + CRF + AscAoD (mean log-loss difference = −0.002, 95% credible interval −0.004 to 0.001; >88% probability of a practical improvement in log-loss) [**Supplemental Figure 10, Supplemental Table 5**], demonstrated that the Dissection-PRS meaningfully improved model log-loss. These results were also supported in analyses using a frequentist approach employing the likelihood-ratio test (**Supplemental Table 6**). Finally, model calibration improvement with the addition of the Dissection-PRS was consistent when stratifying by either sex (**Supplemental Figures 11-13, Summary Tables 7-8**) or genetically similar population group (**Supplemental Figures 14-16, Summary Tables 9-10**). These results lead us to conclude that the Dissection-PRS consistently enhances clinical model calibration.

### PRS effect on model discrimination

The ability to determine patients most at risk of dissection, especially among individuals with ascending thoracic aortic dilation, remains elusive. To investigate whether the Dissection-PRS could meaningfully improve clinical prediction model discrimination between individuals with and without dissection, we tested whether its inclusion increased model area under receiver operator characteristic curve (AUROC). The Age + Sex model had an AUROC=0.572 (95% CI 0.547 to 0.597) [**Figure 4**]. The Age + Sex + CRF model had an AUROC=0.634 (95% CI 0.610 to 0.657), whereas the Age + Sex + CRF + Dissection-PRS had an AUROC=0.700 (95% CI 0.677 to 0.722). Similarly, the Age + Sex + CRF + AscAoD model had an AUROC=0.676 (95% CI 0.651 to 0.702), whereas the Age + Sex + CRF + AscAoD + Dissection-PRS model had an AUROC=0.723 (95% CI 0.702 to 0.744). To determine if the addition of the PRS meaningfully improved model discrimination, we once more employed cross-model Bayesian analysis of AUROC. The addition of the Dissection-PRS to the Age + Sex + CRF model (AUROC mean difference = 0.066, 95% credible interval 0.057 to 0.075; >95% probability of a practical improvement in AUROC) and the Age + Sex + CRF + AscAoD (AUROC mean difference = 0.047, 95% credible interval 0.037 to 0.056; >95% probability of a practical improvement in AUROC) [**Supplemental Figure 17, Supplemental Table 11**], demonstrated that the Dissection-PRS meaningfully improved model AUROC. These results were supported with frequentist analysis of differences in AUROC using the DeLong method (**Supplemental Table 12**). Models were once more compared when stratifying the cohort by either sex (**Supplemental Figures 18-20, Supplemental Tables 13-14**) or genetically similar population group (**Supplemental Figure 21-23, Supplemental Tables 15-16**) with consistent results. These results lead us to conclude that the Dissection-PRS consistently enhances clinical model discrimination.

**Figure 4:**
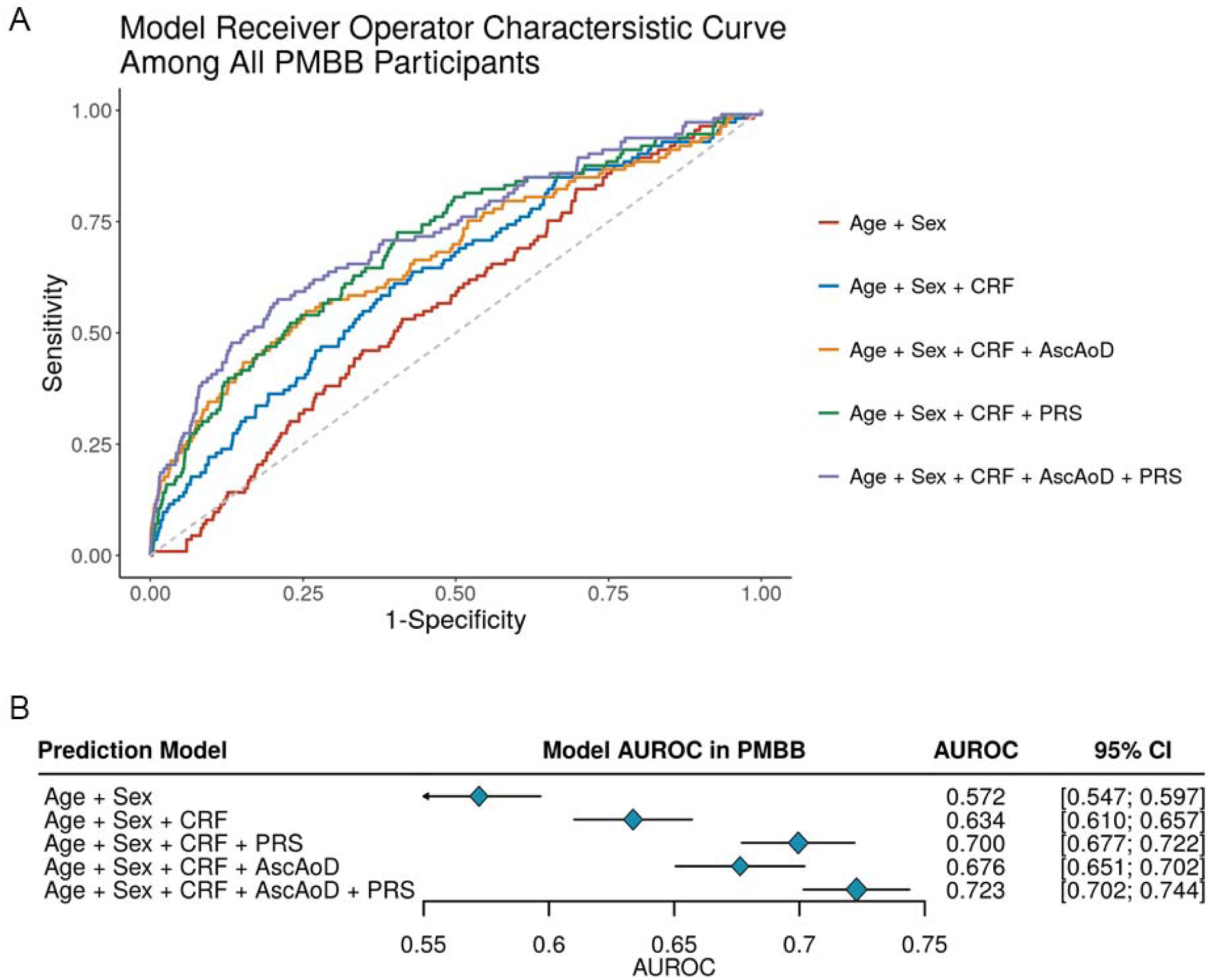
Logistic regression model receiver operator characteristic curve (ROC) and area under the ROC curve to predict thoracic aortic dissection. (A) Receiver operator characteristic curves for each of the primary models including Age + Sex, Age + Sex + CRF, Age + Sex + CRF + Dissection-PRS, Age + Sex + CRF + AscAoD, and Age + Sex + CRF + AscAoD + Dissection-PRS. (B) Corresponding area under the ROC curves with error bars demonstrating 95% confidence intervals for models utilized among all individuals. AscAoD = ascending thoracic aortic diameter; AUROC = area under the receiver operator characteristic curve; CI = 95% confidence interval; CRF = Clinical Risk Factors; Dissection-PRS = thoracic aortic dissection polygenic risk score.

### Incident thoracic aortic dissection analysis

As a sensitivity analysis we performed time-to-event analyses using a Cox proportional hazard model among individuals with incident dissection in the PMBB and AOU. In the PMBB, for every 1 sd increase in Dissection-PRS, the hazard ratio (HR) was 2.30 (95% CI 1.99 to 2.64, *P*<0.001) and 2.27 when adjusting for CRF (95% CI 1.96 to 2.62, *P*<0.001) [**Supplemental Figure 24A**]. When this analysis was restricted to individuals with measured AscAoD prior to dissection, and AscAoD was adjusted for with or without CRF, the effect of the Dissection-PRS remained significant (adjusted for AscAoD: HR=1.65, 95% CI 1.08 to 2.53, P=0.021; adjusted for AscAoD + CRF: HR=1.74, 95% CI 1.12 to 2.70, *P*=0.013) [**Supplemental Figure 24B**]. These incident analysis results were consistent among individuals in the AOU validation cohort where a 1 sd increase in Dissection-PRS yielded an HR of 1.36 (95% CI 1.10 to 1.67, P=0.005). Taken together, these results demonstrate that the Dissection-PRS is associated with incident thoracic aortic dissection across two diverse cohorts.

## Discussion

Identification of diameter-independent risk factors for dissection remains one of the key unresolved challenges in preventing premature deaths due to thoracic aortic disease. In this study, we performed a GWAS-by-subtraction that identified 43 genetic loci associated with diameter-independent thoracic aortic disease, 20 of which were previously unreported. From our summary statistics, we created a PRS that strongly associated with dissection in the PMBB independent of clinical variables including AscAoD and validated our findings in AOU. We then demonstrated that inclusion of the Dissection-PRS in a clinical risk prediction model improves model calibration and discrimination across diverse populations.

The explanation for the improvement in the strength of association with prevalent and incident dissection provided by the Dissection-PRS, as compared to the TAAD-PRS or AscAoD-PRS, is likely partially explained by the identification of novel variants associated with dissection using the GWAS-by-subtraction methodology. However, some of the improvement is also likely due to the subtle statistical reweighting of known variants such that the weights are substantially changed and reflected in the overall observed PRS effect. For example, the effect estimate of the *CDH13* risk locus in the TAAD GWAS was −0.12 while the effect estimate in our GWAS-by-subtraction was −0.14. The enhancement in PRS performance raises the prospect of a true dissection GWAS, rather than the present one inferred through latent values, and whether a PRS from such a GWAS would exceed what we have shown in this present study. In future work, it will also be interesting to attempt to de-convolve the Dissection-PRS into recognizable features (e.g., features of aortic geometry; properties of the aortic wall; etc.).

Determining which patients should receive prophylactic surgical intervention to prevent an acute dissection, and the timing of that intervention, represents an ongoing challenge in cardiovascular medicine and surgery. While the standard AscAoD size threshold for repair remains 5-5.5 cm,^12^ it is clear from retrospective data that this threshold is insufficient to identify at-risk individuals.^13^ Despite myriad attempts to develop algorithms to better predict dissection risk,^34–37^ there remains limited clinical implementation of risk factors other than AscAoD, rate of diameter growth, and family history. Genetic data is changing the way we interpret multi-factorial disease risk. Our results suggest that using common genetic risk factors aggregated in a PRS can contribute to improved assessment of dissection risk and may enhance patient selection for surgical intervention.

Importantly, the Dissection-PRS had a similar effect on dissection risk across diverse populations. This was true in incident analyses across both the primary and validation cohorts, as well as prevalent analyses in the primary cohort. We also observed robust prediction model discrimination improvement with the addition of the Dissection-PRS in both the EUR and AFR population groups. Extensive investigation has been performed on the lack of portability of PRS across diverse populations.^38,39^ The present multi-trait GWAS-by-subtraction is benefited by the diversity inherent in the previously published TAAD GWAS in the MVP and not penalized by subtracting out AoD using summary statistics from a GWAS largely performed in an EUR population. The use of multi-trait GWAS from more diverse parent GWAS data to derive equitable PRS may improve PRS portability as biobanks continue to work to overcome historical lack of diversity.

Determining if a prediction model that includes a PRS improves patient identification will require prospective investigation. For example, future studies may benefit from incorporating a model that includes a PRS for thoracic aortic dissection among patients at risk of dissection to identify those patients who could be considered for expedited repair at a lower diameter threshold. One way to address this is to prospectively enroll individuals at high volume aortic centers with dilated ascending thoracic aortas to have genetic testing and PRS calculation, and integrating the individual PRS into a model to predict dissection risk. Subsequently, individuals would be followed over time to determine the utility of such a model. As thoracic aortic dissection is a relatively rare event, coordinated efforts across many different aortic centers may be better powered to demonstrate an effect.

### Limitations

This study has several limitations. First, our GWAS-by-subtraction is not a true GWAS of dissection and should not be misconstrued as serving that purpose, although it will be interesting to compare these findings with future thoracic dissection GWAS as they become available. Second, the PMBB is enriched for cases of dissection and the robust association of the Dissection-PRS with prevalent and incident dissection may be overestimated in this cohort. Third, dissection is a relatively rare event and therefore any understanding of the true predictive capacity of the Dissection-PRS would be best evaluated prospectively. Fourth, for the TTE data in the PMBB, each TTE was interpreted by a cardiologist in a clinical echocardiography lab, which is accredited by the Intersocietal Commission for the Accreditation of Echocardiography Labs and staffed by cardiologists who are board-certified in echocardiography. However, each TTE was not interpreted by the same cardiologist and small differences in measurement and interpretation may exist between studies. Additionally, although we used the largest and most diverse GWAS studies available to perform our GWAS-by-subtraction, the AscAoD and DesAoD GWAS were performed in the UKB which is almost entirely composed of individuals genetically similar to the 1000G EUR reference population. *Conclusions*

Our findings suggest that our GWAS-by-subtraction allowed the construction of a Dissection-PRS that associates with prevalent dissection. Using this PRS, we can better predict individuals at increased risk of acute dissection. Future clinical implementation of our model requires prospective studies to investigate the benefit of identifying patients at increased risk of dissection and offering earlier surgical intervention.

## Data Availability

The summary statistics for the GWAS-by-subtraction will be made publicly available on Zenodo. The PRS weights will be submitted to the Polygenic Score Catalog. All other data may be made available upon reasonable request to the corresponding author.

## Supporting information

Supplemental Methods, Supplemental Results, Supplemental Tables 3-16, Supplemental Figures 1-24

## Abbreviations

AoD: Aortic diameter
AscAoD: Ascending aortic diameter
AUROC: Area under the receiver operator characteristic curve
CI: Confidence interval
CT: Computed tomography
CRF: Clinical Risk Factors
DesAoD: Descending aortic diameter
PRS: Polygenic risk score
ROC: Receiver operator characteristic curve
SEM: Structural equation modeling
TAAD: Thoracic aortic aneurysm and dissection
TTE: Transthoracic echocardiography

## Acknowledgements

We acknowledge Perline Demange, PhD, Andrew D. Grotzinger, PhD, Michael G. Nivard, PhD, and Elliot M. Tucker-Drob, PhD who assisted in the methodology of GWAS-by-subtraction and maintain the Genomic Structural Equation Modeling package in R. We acknowledge the Penn Medicine BioBank (PMBB) for providing data and thank the patient-participants of Penn Medicine who consented to participate in this research program. We would also like to thank the Penn Medicine BioBank team and Regeneron Genetics Center for providing genetic variant data for analysis. The PMBB is approved under IRB protocol# 813913 and supported by Perelman School of Medicine at University of Pennsylvania, a gift from the Smilow family, and the National Center for Advancing Translational Sciences of the National Institutes of Health under CTSA award number UL1TR001878. The summary statistics for the TAAD GWAS performed in the MVP were obtained through dbGaP, accession number phs001672.v11.p1. The authors thank the MVP staff, researchers, and volunteers who have contributed to MVP, and especially participants who previously served their country in the military and now generously agreed to enroll in the study. (See https://www.research.va.gov/mvp/ for more details). This research is based on data from the Million Veteran Program, Office of Research and Development, Veterans Health Administration, and was supported by the Veterans Administration (VA) MVP award #000. The All of Us Research Program is supported by the National Institutes of Health, Office of the Director: Regional Medical Centers: 1 OT2 OD026549; 1 OT2 OD026554; 1 OT2 OD026557; 1 OT2 OD026556; 1 OT2 OD026550; 1 OT2 OD 026552; 1 OT2 OD026553; 1 OT2 OD026548; 1 OT2 OD026551; 1 OT2 OD026555; IAA #: AOD 16037; Federally Qualified Health Centers: HHSN 263201600085U; Data and Research Center: 5 U2C OD023196; Biobank: 1 U24 OD023121; The Participant Center: U24 OD023176; Participant Technology Systems Center: 1 U24 OD023163; Communications and Engagement: 3 OT2 OD023205; 3 OT2 OD023206; and Community Partners: 1 OT2 OD025277; 3 OT2 OD025315; 1 OT2 OD025337; 1 OT2 OD025276. We gratefully acknowledge *All of Us* participants for their contributions, without whom this research would not have been possible. We also thank the National Institutes of Health’s *<u>All of Us</u>* <u>Research Program</u> for making available the participant data examined in this study.

## Sources of Funding

J.D. is supported by the American Heart Association (23POST1011251). The views expressed in this article are those of the authors and do not necessarily reflect the position or policy of the Department of Veterans Affairs or the United States government. W.R.W. is supported by NIH R01 HL171709, P41 EB029460, and R01 HL169378. D.M.M. and D.C.G. are supported by the NIH (R01HL109942), the John Ritter Foundation. M.G.L. received support from the Doris Duke Foundation (2023-0224) and Department of Veterans Affairs Biomedical Research and Development Award IK2-BX006551. J.P.P is supported by the NIH (K08HL159346).

## Disclosures

M.G.L. receives research support to his institution from MyOme outside of this work. S.M.D receives research support to his institution from RenalytixAI and in kind support from Novo Nordisk, both outside of this work. SMD is named as a co-inventor on a government-owned US Patent application related to the use of genetic risk prediction for venous thromboembolic disease filed by the US Department of Veterans Affairs in accordance with Federal regulatory requirements.

## Supplemental Material

### Supplement 1

- Supplemental table 1-2

### Supplement 2

- Supplemental Methods

- Supplemental Results

- Supplemental Authors

- Supplemental Tables 3-16

- Supplemental Figures 1-24

## Notes

### Competing Interest Statement

The authors have declared no competing interest.

### Author Declarations

The IRB of the University of Pennsylvania gave ethical approval for the Penn Medicine Biobank analysis in this work (Protocol #813913). The IRB of University of California San Francisco gave ethical approval for the All of Us analysis in this work (Protocol #22-37715).

## References

1. Gudbjartsson T, Ahlsson A, Geirsson A, Gunn J, Hjortdal V, Jeppsson A, Mennander A, Zindovic I, Olsson C. Acute type A aortic dissection – a review. Scandinavian Cardiovascular Journal. 2020;54:1–13. doi: 10.1080/14017431.2019.1660401

2. DeMartino RR, Sen I, Huang Y, Bower TC, Oderich GS, Pochettino A, Greason K, Kalra M, Johnstone J, Shuja F, et al. Population-Based Assessment of the Incidence of Aortic Dissection, Intramural Hematoma, and Penetrating Ulcer, and Its Associated Mortality From 1995 to 2015. Circ Cardiovasc Qual Outcomes. 2018;11:e004689. doi: 10.1161/CIRCOUTCOMES.118.004689

3. Clouse WD, Hallett JW, Jr., Schaff HV, Spittell PC, Rowland CM, Ilstrup DM, Melton LJ, 3rd. Acute aortic dissection: population-based incidence compared with degenerative aortic aneurysm rupture. Mayo Clin Proc. 2004;79:176–180. doi: 10.4065/79.2.176

4. Olsson C, Thelin S, Ståhle E, Ekbom A, Granath F. Thoracic aortic aneurysm and dissection: increasing prevalence and improved outcomes reported in a nationwide population-based study of more than 14,000 cases from 1987 to 2002. Circulation. 2006;114:2611–2618. doi: 10.1161/circulationaha.106.630400

5. Mody PS, Wang Y, Geirsson A, Kim N, Desai MM, Gupta A, Dodson JA, Krumholz HM. Trends in aortic dissection hospitalizations, interventions, and outcomes among medicare beneficiaries in the United States, 2000-2011. Circ Cardiovasc Qual Outcomes. 2014;7:920–928. doi: 10.1161/circoutcomes.114.001140

6. Pape Linda A, Awais M, Woznicki Elise M, Suzuki T, Trimarchi S, Evangelista A, Myrmel T, Larsen M, Harris Kevin M, Greason K, et al. Presentation, Diagnosis, and Outcomes of Acute Aortic Dissection. Journal of the American College of Cardiology. 2015;66:350–358. doi: 10.1016/j.jacc.2015.05.029

7. McClure RS, Ouzounian M, Boodhwani M, El-Hamamsy I, Chu MWA, Pozeg Z, Dagenais F, Sikdar KC, Appoo JJ. Cause of Death Following Surgery for Acute Type A Dissection: Evidence from the Canadian Thoracic Aortic Collaborative. Aorta (Stamford). 2017;5:33–41. doi: 10.12945/j.aorta.2017.16.034

8. Verstraeten A, Luyckx I, Loeys B. Aetiology and management of hereditary aortopathy. Nat Rev Cardiol. 2017;14:197–208. doi: 10.1038/nrcardio.2016.211

9. Melvinsdottir IH, Lund SH, Agnarsson BA, Sigvaldason K, Gudbjartsson T, Geirsson A. The incidence and mortality of acute thoracic aortic dissection: results from a whole nation study. Eur J Cardiothorac Surg. 2016;50:1111–1117. doi: 10.1093/ejcts/ezw235

10. Howard DP, Banerjee A, Fairhead JF, Perkins J, Silver LE, Rothwell PM, Oxford Vascular S. Population-based study of incidence and outcome of acute aortic dissection and premorbid risk factor control: 10-year results from the Oxford Vascular Study. Circulation. 2013;127:2031–2037. doi: 10.1161/CIRCULATIONAHA.112.000483

11. Evangelista A, Isselbacher EM, Bossone E, Gleason TG, Eusanio MD, Sechtem U, Ehrlich MP, Trimarchi S, Braverman AC, Myrmel T, et al. Insights From the International Registry of Acute Aortic Dissection: A 20-Year Experience of Collaborative Clinical Research. Circulation. 2018;137:1846–1860. doi: 10.1161/CIRCULATIONAHA.117.031264

12. Isselbacher EM, Preventza O, Hamilton Black J, 3rd, Augoustides JG, Beck AW, Bolen MA, Braverman AC, Bray BE, Brown-Zimmerman MM, Chen EP, et al. 2022 ACC/AHA Guideline for the Diagnosis and Management of Aortic Disease: A Report of the American Heart Association/American College of Cardiology Joint Committee on Clinical Practice Guidelines. Circulation. 2022;146:e334–e482. doi: 10.1161/CIR.0000000000001106

13. Pape LA, Tsai TT, Isselbacher EM, Oh JK, O’Gara P T, Evangelista A, Fattori R, Meinhardt G, Trimarchi S, Bossone E, et al. Aortic diameter >or = 5.5 cm is not a good predictor of type A aortic dissection: observations from the International Registry of Acute Aortic Dissection (IRAD). Circulation. 2007;116:1120–1127. doi: 10.1161/circulationaha.107.702720

14. Wolford BN, Hornsby WE, Guo D, Zhou W, Lin M, Farhat L, McNamara J, Driscoll A, Wu X, Schmidt EM, et al. Clinical Implications of Identifying Pathogenic Variants in Individuals With Thoracic Aortic Dissection. Circ Genom Precis Med. 2019;12:e002476. doi: 10.1161/CIRCGEN.118.002476

15. Renard M, Francis C, Ghosh R, Scott AF, Witmer PD, Ades LC, Andelfinger GU, Arnaud P, Boileau C, Callewaert BL, et al. Clinical Validity of Genes for Heritable Thoracic Aortic Aneurysm and Dissection. J Am Coll Cardiol. 2018;72:605–615. doi: 10.1016/j.jacc.2018.04.089

16. Pinard A, Jones GT, Milewicz DM. Genetics of Thoracic and Abdominal Aortic Diseases. Circ Res. 2019;124:588–606. doi: 10.1161/CIRCRESAHA.118.312436

17. Pirruccello JP, Chaffin MD, Chou EL, Fleming SJ, Lin H, Nekoui M, Khurshid S, Friedman SF, Bick AG, Arduini A, et al. Deep learning enables genetic analysis of the human thoracic aorta. Nat Genet. 2022;54:40–51. doi: 10.1038/s41588-021-00962-4

18. Tcheandjieu C, Xiao K, Tejeda H, Lynch JA, Ruotsalainen S, Bellomo T, Palnati M, Judy R, Klarin D, Kember RL, et al. High heritability of ascending aortic diameter and trans-ancestry prediction of thoracic aortic disease. Nat Genet. 2022;54:772–782. doi: 10.1038/s41588-022-01070-7

19. Klarin D, Devineni P, Sendamarai AK, Angueira AR, Graham SE, Shen YH, Levin MG, Pirruccello JP, Surakka I, Karnam PR, et al. Genome-wide association study of thoracic aortic aneurysm and dissection in the Million Veteran Program. Nat Genet. 2023;55:1106–1115. doi: 10.1038/s41588-023-01420-z

20. Verma A, Damrauer SM, Naseer N, Weaver J, Kripke CM, Guare L, Sirugo G, Kember RL, Drivas TG, Dudek SM, et al. The Penn Medicine BioBank: Towards a Genomics-Enabled Learning Healthcare System to Accelerate Precision Medicine in a Diverse Population. Journal of Personalized Medicine. 2022;12:1974.

21. Denny JC, Devaney SA, Gebo KA. The “All of Us” Research Program. Reply. N Engl J Med. 2019;381:1884–1885. doi: 10.1056/NEJMc1912496

22. Zhou Z, Cecchi AC, Prakash SK, Milewicz DM. Risk Factors for Thoracic Aortic Dissection. Genes (Basel). 2022;13. doi: 10.3390/genes13101814

23. Takada M, Yamagishi K, Tamakoshi A, Iso H, Group JS. Body Mass Index and Mortality From Aortic Aneurysm and Dissection. J Atheroscler Thromb. 2021;28:338–348. doi: 10.5551/jat.57232

24. Zafar MA, Li Y, Rizzo JA, Charilaou P, Saeyeldin A, Velasquez CA, Mansour AM, Bin Mahmood SU, Ma WG, Brownstein AJ, et al. Height alone, rather than body surface area, suffices for risk estimation in ascending aortic aneurysm. J Thorac Cardiovasc Surg. 2018;155:1938–1950. doi: 10.1016/j.jtcvs.2017.10.140

25. Pirruccello JP, Lin H, Khurshid S, Nekoui M, Weng LC, Vasan RS, Isselbacher EM, Benjamin EJ, Lubitz SA, Lindsay ME, Ellinor PT. Development of a Prediction Model for Ascending Aortic Diameter Among Asymptomatic Individuals. JAMA. 2022;328:1935–1944. doi: 10.1001/jama.2022.19701

26. Beeche C, Dib MJ, Zhao B, Azzo JD, Maynard H, Duda J, Gee J, Salman O, Penn Medicine B, Witschey WR, Chirinos JA. Three-dimensional aortic geometry: clinical correlates, prognostic value and genetic architecture. bioRxiv. 2024. doi: 10.1101/2024.05.09.593413

27. Demange PA, Malanchini M, Mallard TT, Biroli P, Cox SR, Grotzinger AD, Tucker-Drob EM, Abdellaoui A, Arseneault L, van Bergen E, et al. Investigating the genetic architecture of noncognitive skills using GWAS-by-subtraction. Nat Genet. 2021;53:35–44. doi: 10.1038/s41588-020-00754-2

28. St Pourcain B, Eaves LJ, Ring SM, Fisher SE, Medland S, Evans DM, Davey Smith G. Developmental Changes Within the Genetic Architecture of Social Communication Behavior: A Multivariate Study of Genetic Variance in Unrelated Individuals. Biol Psychiatry. 2018;83:598–606. doi: 10.1016/j.biopsych.2017.09.020

29. Grotzinger AD, Rhemtulla M, de Vlaming R, Ritchie SJ, Mallard TT, Hill WD, Ip HF, Marioni RE, McIntosh AM, Deary IJ, et al. Genomic structural equation modelling provides insights into the multivariate genetic architecture of complex traits. Nat Hum Behav. 2019;3:513–525. doi: 10.1038/s41562-019-0566-x

30. Ruan Y, Lin YF, Feng YA, Chen CY, Lam M, Guo Z, Stanley Global Asia I, He L, Sawa A, Martin AR, et al. Improving polygenic prediction in ancestrally diverse populations. Nat Genet. 2022;54:573–580. doi: 10.1038/s41588-022-01054-7

31. Lambert SA, Gil L, Jupp S, Ritchie SC, Xu Y, Buniello A, McMahon A, Abraham G, Chapman M, Parkinson H, et al. The Polygenic Score Catalog as an open database for reproducibility and systematic evaluation. Nature Genetics. 2021;53:420–425. doi: 10.1038/s41588-021-00783-5

32. Fairley S, Lowy-Gallego E, Perry E, Flicek P. The International Genome Sample Resource (IGSR) collection of open human genomic variation resources. Nucleic Acids Res. 2020;48:D941–D947. doi: 10.1093/nar/gkz836

33. Therneau TM, Grambsch PM. The Cox Model. In: Therneau TM, Grambsch PM, eds. Modeling Survival Data: Extending the Cox Model. New York, NY: Springer New York; 2000:39–77.

34. von Kodolitsch Y, Schwartz AG, Nienaber CA. Clinical prediction of acute aortic dissection. Arch Intern Med. 2000;160:2977–2982. doi: 10.1001/archinte.160.19.2977

35. Adriaans BP, Wildberger JE, Westenberg JJM, Lamb HJ, Schalla S. Predictive imaging for thoracic aortic dissection and rupture: moving beyond diameters. Eur Radiol. 2019;29:6396–6404. doi: 10.1007/s00330-019-06320-7

36. Heuts S, Adriaans BP, Rylski B, Mihl C, Bekkers S, Olsthoorn JR, Natour E, Bouman H, Berezowski M, Kosiorowska K, et al. Evaluating the diagnostic accuracy of maximal aortic diameter, length and volume for prediction of aortic dissection. Heart. 2020;106:892–897. doi: 10.1136/heartjnl-2019-316251

37. Wu J, Qiu J, Xie E, Jiang W, Zhao R, Qiu J, Zafar MA, Huang Y, Yu C. Predicting in-hospital rupture of type A aortic dissection using Random Forest. J Thorac Dis. 2019;11:4634–4646. doi: 10.21037/jtd.2019.10.82

38. Popejoy AB, Fullerton SM. Genomics is failing on diversity. Nature. 2016;538:161–164. doi: 10.1038/538161a

39. Ding Y, Hou K, Xu Z, Pimplaskar A, Petter E, Boulier K, Prive F, Vilhjalmsson BJ, Olde Loohuis LM, Pasaniuc B. Polygenic scoring accuracy varies across the genetic ancestry continuum. Nature. 2023;618:774–781. doi: 10.1038/s41586-023-06079-4

40. Purcell S, Neale B, Todd-Brown K, Thomas L, Ferreira MA, Bender D, Maller J, Sklar P, de Bakker PI, Daly MJ, Sham PC. PLINK: a tool set for whole-genome association and population-based linkage analyses. Am J Hum Genet. 2007;81:559–575. doi: 10.1086/519795

41. Genomes Project C, Abecasis GR, Auton A, Brooks LD, DePristo MA, Durbin RM, Handsaker RE, Kang HM, Marth GT, McVean GA. An integrated map of genetic variation from 1,092 human genomes. Nature. 2012;491:56–65. doi: 10.1038/nature11632

42. Pruim RJ, Welch RP, Sanna S, Teslovich TM, Chines PS, Gliedt TP, Boehnke M, Abecasis GR, Willer CJ. LocusZoom: regional visualization of genome-wide association scan results. Bioinformatics. 2010;26:2336–2337. doi: 10.1093/bioinformatics/btq419

43. Lawrence M, Huber W, Pages H, Aboyoun P, Carlson M, Gentleman R, Morgan MT, Carey VJ. Software for computing and annotating genomic ranges. PLoS Comput Biol. 2013;9:e1003118. doi: 10.1371/journal.pcbi.1003118

44. de Leeuw CA, Mooij JM, Heskes T, Posthuma D. MAGMA: generalized gene-set analysis of GWAS data. PLoS Comput Biol. 2015;11:e1004219. doi: 10.1371/journal.pcbi.1004219

45. Consortium GT. The GTEx Consortium atlas of genetic regulatory effects across human tissues. Science. 2020;369:1318–1330. doi: 10.1126/science.aaz1776

46. Wang G, Sarkar A, Carbonetto P, Stephens M. A simple new approach to variable selection in regression, with application to genetic fine mapping. J R Stat Soc Series B Stat Methodol. 2020;82:1273–1300. doi: 10.1111/rssb.12388

47. Wallace C. A more accurate method for colocalisation analysis allowing for multiple causal variants. PLoS Genet. 2021;17:e1009440. doi: 10.1371/journal.pgen.1009440

48. Dewey FE, Gusarova V, O’Dushlaine C, Gottesman O, Trejos J, Hunt C, Van Hout CV, Habegger L, Buckler D, Lai KM, et al. Inactivating Variants in ANGPTL4 and Risk of Coronary Artery Disease. N Engl J Med. 2016;374:1123–1133. doi: 10.1056/NEJMoa1510926

49. Verma A, Damrauer SM, Naseer N, Weaver J, Kripke CM, Guare L, Sirugo G, Kember RL, Drivas TG, Dudek SM, et al. The Penn Medicine BioBank: Towards a Genomics-Enabled Learning Healthcare System to Accelerate Precision Medicine in a Diverse Population. J Pers Med. 2022;12. doi: 10.3390/jpm12121974

50. Loh P-R, Danecek P, Palamara PF, Fuchsberger C, A Reshef Y, K Finucane H, Schoenherr S, Forer L, McCarthy S, Abecasis GR, et al. Reference-based phasing using the Haplotype Reference Consortium panel. Nature Genetics. 2016;48:1443–1448. doi: 10.1038/ng.3679

51. Das S, Forer L, Schönherr S, Sidore C, Locke AE, Kwong A, Vrieze SI, Chew EY, Levy S, McGue M, et al. Next-generation genotype imputation service and methods. Nature Genetics. 2016;48:1284–1287. doi: 10.1038/ng.3656

52. Price AL, Patterson NJ, Plenge RM, Weinblatt ME, Shadick NA, Reich D. Principal components analysis corrects for stratification in genome-wide association studies. Nature Genetics. 2006;38:904–909. doi: 10.1038/ng1847

53. Karczewski KJ, Francioli LC, Tiao G, Cummings BB, Alfoldi J, Wang Q, Collins RL, Laricchia KM, Ganna A, Birnbaum DP, et al. The mutational constraint spectrum quantified from variation in 141,456 humans. Nature. 2020;581:434–443. doi: 10.1038/s41586-020-2308-7

54. Ioannidis NM, Rothstein JH, Pejaver V, Middha S, McDonnell SK, Baheti S, Musolf A, Li Q, Holzinger E, Karyadi D, et al. REVEL: An Ensemble Method for Predicting the Pathogenicity of Rare Missense Variants. Am J Hum Genet. 2016;99:877–885. doi: 10.1016/j.ajhg.2016.08.016

55. Richards S, Aziz N, Bale S, Bick D, Das S, Gastier-Foster J, Grody WW, Hegde M, Lyon E, Spector E, et al. Standards and guidelines for the interpretation of sequence variants: a joint consensus recommendation of the American College of Medical Genetics and Genomics and the Association for Molecular Pathology. Genet Med. 2015;17:405–424. doi: 10.1038/gim.2015.30

56. All of Us Research Program Genomics I. Genomic data in the All of Us Research Program. Nature. 2024;627:340–346. doi: 10.1038/s41586-023-06957-x

57. Venner E, Patterson K, Kalra D, Wheeler MM, Chen YJ, Kalla SE, Yuan B, Karnes JH, Walker K, Smith JD, et al. The frequency of pathogenic variation in the All of Us cohort reveals ancestry-driven disparities. Commun Biol. 2024;7:174. doi: 10.1038/s42003-023-05708-y

58. Khan A, Turchin MC, Patki A, Srinivasasainagendra V, Shang N, Nadukuru R, Jones AC, Malolepsza E, Dikilitas O, Kullo IJ, et al. Genome-wide polygenic score to predict chronic kidney disease across ancestries. Nat Med. 2022;28:1412–1420. doi: 10.1038/s41591-022-01869-1

59. Marcot BG, Hanea AM. What is an optimal value of k in k-fold cross-validation in discrete Bayesian network analysis? Computational Statistics. 2021;36:2009–2031. doi: 10.1007/s00180-020-00999-9

60. Kuhn MW, H. Tidymodels: a collection of packages for modeling and machine learning using tidyverse principles. 2020.

61. Wilimitis D, Walsh CG. Practical Considerations and Applied Examples of Cross-Validation for Model Development and Evaluation in Health Care: Tutorial. JMIR AI. 2023;2:e49023. doi: 10.2196/49023

62. Kruschke JK, Liddell TM. The Bayesian New Statistics: Hypothesis testing, estimation, meta-analysis, and power analysis from a Bayesian perspective. Psychon Bull Rev. 2018;25:178–206. doi: 10.3758/s13423-016-1221-4

63. Kruschke JK. Rejecting or Accepting Parameter Values in Bayesian Estimation. Advances in Methods and Practices in Psychological Science. 2018;1:270–280. doi: 10.1177/2515245918771304

64. Kuhn M. tidyposterior: Bayesian Analysis to Compare Models using Resampling Statistics. 2022.

65. Harrell F. Statistically Efficient Ways to Quantify Added Predictive Value of New Measurements. In: Statistical Thinking. 2018.

66. Harrell JFE. Regression Modeling Strategies : With Applications to Linear Models, Logistic and Ordinal Regression, and Survival Analysis. In: Springer Series in Statistics,. Cham: Springer International Publishing: Imprint: Springer,; 2015:1 online resource (XXV, 582 pages 157 illustrations, 553 illustrations in color.

67. Fronczek J, Polok K, de Lange DW, Jung C, Beil M, Rhodes A, Fjølner J, Górka J, Andersen FH, Artigas A, et al. Relationship between the Clinical Frailty Scale and short-term mortality in patients ≥ 80 years old acutely admitted to the ICU: a prospective cohort study. Critical Care. 2021;25:231. doi: 10.1186/s13054-021-03632-3

68. DeLong ER, DeLong DM, Clarke-Pearson DL. Comparing the areas under two or more correlated receiver operating characteristic curves: a nonparametric approach. Biometrics. 1988;44:837–845.

69. Akashi M, Higashi T, Masuda S, Komori T, Furuse M. A coronary artery disease-associated gene product, JCAD/KIAA1462, is a novel component of endothelial cell-cell junctions. Biochem Biophys Res Commun. 2011;413:224–229. doi: 10.1016/j.bbrc.2011.08.073

70. Douglas G, Mehta V, Al Haj Zen A, Akoumianakis I, Goel A, Rashbrook VS, Trelfa L, Donovan L, Drydale E, Chuaiphichai S, et al. A key role for the novel coronary artery disease gene JCAD in atherosclerosis via shear stress mechanotransduction. Cardiovasc Res. 2020;116:1863–1874. doi: 10.1093/cvr/cvz263

71. Guo B, Greenwood PL, Cafe LM, Zhou G, Zhang W, Dalrymple BP. Transcriptome analysis of cattle muscle identifies potential markers for skeletal muscle growth rate and major cell types. BMC Genomics. 2015;16:177. doi: 10.1186/s12864-015-1403-x

