## Supplemental Methods, Supplemental Results, Supplemental Tables 3-16, Supplemental Figures 1-24 for "Predicting thoracic aortic dissection in a diverse biobank using a polygenic risk score"

*Lead loci identification and gene prioritization*

Genomic risk loci were identified using the MRC-IEU “ld_clump_local” command in the MRCIEU/ieugwasr package in R (https://github.com/MRCIEU/ieugwasr) that relies on the “clump” command in PLINK,^40^ and the 1000 Genomes Project^41^ Phase 3 reference panel (*P* < 5 × 10^−8^; window 500 kb. *r*^2^ = 0001). Independent loci were represented in a Manhattan plot and loci unique from the original ascending aortic diameter (AscAoD), descending aortic diameter (DesAoD), and thoracic aortic aneurysm and dissection (TAAD) summary statistics were identified visually. A quantile-quantile plot was also generated and $\lambda$gc calculated to determine the degree of genomic inflation. Individual lead loci regional association plots were compared to equivalent regions identified in the separate parent GWAS of TAAD, AscAoD, and DesAoD using the locuszoom package (version 0.3.1).^42^ Gene prioritization was performed at each lead locus combining the following metrics: nearest gene, deleterious coding variants in moderate or high linkage disequilibrium (LD) with the lead variant at each locus, and output from the Multi-marker Analysis of GenoMic Annotation (MAGMA). The nearest gene to each loci was identified using the nearest command in the Genomic Ranges package in R.^43^ The nearest gene was then compared to the constituent trait GWAS to determine if any variant in the 500kb flanking the lead locus is genome-wide significant. Next, we determined which of these variants was in high (*r*^2^ > 0.8) or moderate (0.8 > *r*^2^ > 0.3) linkage disequilibrium (LD) with coding variants from the assigned nearest gene using the LDLinkR (v1.4.0) package in R . Finally, MAGMA^44^ was performed to validate lead loci gene prioritization using aggregated variants within each gene to determine which genes were most likely involved the trait of interest.

*Aortic tissue eQTL colocalization*

We performed two-step colocalization with aortic tissue expression quantitative trait loci (eQTLs) from Genotype-Tissue Expression (GTEx) v8 at select loci.^45^ First, we performed select lead loci fine-mapping using the sum of single effects (SuSiE)^46^ to derive credible sets at the specific loci of interest. We then performed SuSiE Colocalization,^47^ which relaxes the single variant assumption at lead loci, to determine the posterior probability of GWAS-by-subtraction and aortic tissue eQTL colocalization. Colocalization results are reported in standard posterior probability form where PP.H0 = no signal in either dataset, PP.H1 = signal in the first dataset, PP.H2 = signal in the second dataset, PP.H3 = colocalization at locus but different lead variants, or PP.H4 = colocalization with shared lead variants.

*PMBB Genetic Data*

Whole exome sequencing was performed as previously described by Regeneron Genomics Center (RGC).^48^ Individual patient DNA samples were processed and sequenced on the Illumina NovaSeq 6000 (Albany, NY, USA). WeCall variant caller was employed for sequence alignment (GRCh38), variant identification, and genotype assignment.^49^ Quality control exclusions included sex errors, high rates of heterozygosity (D-statistic > 0.4), low sequence coverage, and genetically identified sample duplicates. Single nucleotide variants (SNVs) were filtered for a read depth $\geq$ 7 and were retained if they either had at least one heterozygous variant genotype with an allele balance ratio $\geq$ 0.15, or a homozygous variant genotype. Insertion-deletion variants (INDELs) were filtered for a read depth $\geq$ 10 and either a heterozygous variant genotype with an allele balance $\geq$ 0.20, or a homozygous variant genotype.

Genotype array analysis was performed as previously described by RGC using a genotyping array chip with 654,027 genetic markers with the addition of ancestry-specific informative markers.^20^ Participant samples were genotyped on Illumina Global Screening Array v.2.0 (GSAv2). Sample level quality control was performed and subsequently imputation was performed using Eagle v2.4.1^50^ and Minimac4 version 1.0.0^51^ software. All autosomes were imputed with TOPMed version R2 on a GRCh38 reference panel. Quality control after imputation included removal of palindromic variants, biallelic variant check, sex check, genotype and sample call rate filtering (>99%), minor allele frequency filtering (MAF > 1%), Hardy-Weinberg equilibrium test (P-value > 1x10^-8^), and imputation score check (R2 > 0.7).^20^ Genetic principal components (PCs) to adjust for population structure and inform genetically inferred ancestry was performed using EIGENSOFT version 7.2.0.^52^

*PMBB HTAAD Variant Calls*

Of the more than 30 genes have been associated with hereditary thoracic aortic aneurysm and dissection (HTAAD), we selected the 11 genes that have definitive or strong association with HTAAD based on National Institutes of Health Clinical Genome Resource (ClinGen) Aortopathy Working Group, including *ACTA2, COL3A1, FBN1, LOX, MYH11, MYLK, PRKG1, SMAD3, TGFB2, TGFBR1, and TGFBR2*.^15^ Variants were initially filtered for minor allele frequency (MAF) < 0.001 in the gnomAD database,^53^ and a Rare Exome Variant Ensemble Learner (REVEL)^54^ score ≥ 0.5 for nonsynonymous variants. Subsequently, pathogenic (P) or likely pathogenic (LP) variants were adjudicated based on American College of Medical Genetics classification framework which relies on observed penetrance of given variants, segregation of a specific variant with cases of disease within a family, and presence of the variant in cases observed in unrelated individuals.^16,55^

*PMBB population descriptors*

Individuals of similar genetic ancestry were assembled into population groups based on genetic similarity to reference superpopulations from the 1000 Genomes Project (release 3).^41^ The primary analytic cohort included a European population group (EUR) consisting of 29,947 individuals (69%) genetically similar to the 1000G European reference population and an African population group (AFR) consisting of 11,111 individuals (25%) genetically similar to the 1000G African reference population; no other population group reached a sufficient size for stratified analysis. Analyses accounted for study-level population stratification by ancestry group, controlling for age, sex, and the first five genetic PCs.

*All of Us genetic data*

At the time of analysis, whole genome sequencing (WGS) had been completed in 245,149 *All of Us* (AOU) participants (release “R7”). Sequencing and sample quality control in AOU has been detailed previously.^56,57^ In brief, sequencing was performed with Illumina NovaSeq 6000 to an average depth of 30x. Subsequently the sequencing data was processed with Illumina DRAGEN v3.4.12 for mapping, alignment to GRCh38, duplicate marking, and variant calling. Their internal quality control processes ensure that released data have mean coverage ≥ 30x, ≥90% of the genome at ≥20x, ≥8x10^10^ bases at Q30, and <1% contamination.^56^

*AOU Primary Outcomes and Statistical Analysis*

Thoracic aortic dissection was identified in *All of Us* based on SNOMED code 233994002 (“Dissection of thoracic aorta”). Prevalent dissection, occurring prior to enrollment, was modeled with logistic regression including age at enrollment, sex, genetic PCs 1-5, and the Dissection-PRS. Incident dissection, occurring after enrollment, was modeled with Cox proportional hazards model including the same covariates as above. Cox models that additionally accounted for systolic and diastolic blood pressure, height, weight, body mass index, body surface area, and heart rate were also produced.

*Polygenic Risk Score Creation*

We constructed a PRS from a UKB-derived GWAS of AscAoD performed among 38,694 participants. This PRS contained 1,117,325 variants. We constructed a multi-population PRS for TAAD from a MVP-derived GWAS of TAAD among a diverse population of 461,669 MVP participants (8,626 cases, 453,043 disease-free controls).^19^ This MVP-derived PRS contained 1,274,933 variants. We also constructed a Dissection-PRS from our GWAS-by-subtraction summary statistics presented above. This score contained 1,086,099 variants. For the UKB-derived AscAoD PRS and the Dissection-PRS, genetic variants and their associated weights were selected using PRS-CS-auto using default settings.^30^ For the MVP-derived multi-population TAAD PRS, genetic variants and their associated weights were selected using PRS-CSx-auto using default settings.^30^

*Polygenic risk score residualization in AOU*

The polygenic score weights were applied in 1000G and AOU. In 1000G, following the approach of Khan, *et al*, the first 20 PCs of ancestry were used to predict the polygenic score (model 1). The squared residuals were then predicted again by the first 20 PCs of ancestry (model 2).^58^ Model 1 was then applied in AOU and subtracted from the polygenic score, yielding PC-residualized values. Model 2 was then applied in AOU and the PC-residualized values from Model 1 were divided by the ancestry-adjusted residual standard deviations from Model 2, yielding a polygenic score adjusted for ancestral differences in mean and variance. The final score units are, approximately, standard deviations of the score in 1000G.

*Prediction model calibration and discrimination testing with cross-validation*

To limit over-fitting, models were trained and tested using 5 repeats of 10-fold cross-validation. Cross-validation is a standard resampling method that is easily applied to predictive modeling to minimize bias; 10-fold cross-validation is commonly accepted.^59,60^ Repeated cross-validations provide increasingly conservative bias for model outcomes.^61^ Our repeated cross-validation split the data evenly 10 times, holding out one of 10 folds for model assessment while the data across the remaining nine folds are used to fit the model. As this entire process is repeated five times, the result is 50 separate model fits/tests. Final resampling estimates of performance average each cross-validated replicate with 95% confidence intervals. The cross-validated replicates allow robust comparisons across models using different metrics including log-loss (calibration) and area under the receiver operator characteristic curve (AUROC).

*Assessing model calibration improvement with addition of Dissection-PRS*

Cross-validated log-loss and area under the receiver operator characteristic curve (AUROC) with 95% confidence intervals (CIs) was calculated and compared across all models. To determine the probability of significant differences in calibration between models with or without the Dissection-PRS, we used Bayesian analysis of 5,000 iterations of log-less test between models. Bayesian hypothesis testing prioritizes precision of estimation.^62^ Instead of testing for the ability to reject the null hypothesis that two models of interest are not different, the Bayesian framework provides a direct comparison of different models based on credible intervals, or the range of values that encompass 95% of most likely observed outcomes. Model mean estimates and credible interval comparisons are performed by assigning a region of practical equivalence (ROPE) around one model mean. ROPE is calculated using $\delta= \pm0.1$ where $\delta={(\mu}_{1}-\mu_{2})/\sqrt{(\sigma_{1}^{2}-\sigma_{2}^{2})/2}$ where μ = mean value and σ = standard deviation.^62,63^ For each Bayesian analysis, the addition of the Dissection-PRS to a base model was evaluated.

The Bayesian approach was performed using analysis of variance (ANOVA) function in the *tidyposterior* package in R (v1.0.0) that employs a random model to account for resampling by assuming that individual resamples only effect the model by changing the intercept.^64^ Cross-model comparisons were reported using plots demonstrating model posterior distributions with mean model difference and 95% credible intervals, and a table reporting the probability of practical difference or equivalence based on the calculated ROPE.

*Frequentist analyses*

As a sensitivity analysis, frequentist statistical testing was employed to assess model calibration and discrimination. For model calibration, we used the likelihood ratio $\chi$^2^ test, a simple way to determine model improvement with the addition of a covariate.^65^ The likelihood ratio $\chi$^2^ test can also be used to identify the fraction of new information (FNI) provided by an added covariate,^66,67^ and is calculated by the likelihood ratio of two nested models.^65^ A null result is due to the likelihood ratio being <1 suggesting that the model is not improved by the addition of a covariate. The FNI is then derived by calculating the adequacy index (AI) of the base model, and then calculating FNI = 1-AI. For each likelihood ratio $\chi$^2^ test, the addition of the Dissection-PRS to a base model was evaluated once more. To perform frequentist analysis of model discrimination, we determined base model AUROC improvement with the Dissection-PRS by comparing the AUROC between the two nested models using the non-parametric DeLong method.^68^

**Supplemental Results**

*GWAS-by-subtraction gene prioritization*

Of the lead loci, 20 were unreported at genome-wide significance in the constituent GWAS for either TAAD, AscAoD, or DesAoD (**Figure 1B**).^17,19^ At several of these loci, gene prioritization analyses identified genes which encode proteins known to affect cell signaling pathways, extracellular matrix proteins, or structural proteins (**Supplemental Figure 3, Supplemental Table 2**). For example, we identified a novel association at the *JCAD* locus where gene prioritization techniques consistently prioritized JCAD as the causal gene (**Supplemental Figure 3**), we observed colocalization with aortic tissue eQTLs (**Supplemental Table 3**), and there appeared to be sub-genome-wide significant signal in the parent TAAD GWAS (**Supplemental Figure 4**). The JCAD protein is a junctional cadherin that regulates vascular endothelial homeostasis.^69^ In animal models, *JCAD*-knockout mice demonstrate significantly less atherosclerosis along the ascending aorta and aortic arch, and decreased pro-inflammatory adhesion molecules in areas of disturbed aortic blood flow.^70^ As another example, we identified a novel locus near *SOX8* (**Supplemental Figure 5**) that also colocalized with aortic tissue eQTLs supporting the prioritization of *SOX8* (**Supplemental Table 3**). Little is known regarding the function of SOX8 in aortic tissue, however there is evidence that it is involved in angiogenesis in animal studies.^71^ Taken together, these data suggest the GWAS-by-subtraction methodology identified novel loci associated with thoracic aortic disease.

**Supplemental Authors**

Penn Medicine BioBank Banner Author List and Contribution Statements

PMBB Leadership Team

Daniel J. Rader, M.D., Marylyn D. Ritchie, Ph.D.

Contribution: All authors contributed to securing funding, study design and oversight. All authors reviewed the final version of the manuscript.

Patient Recruitment and Regulatory Oversight

JoEllen Weaver, Nawar Naseer, Ph.D., M.P.H., Afiya Poindexter, Khadijah Hu-Sain, Yi-An Ko, Ph.D.

Contributions: JW manages patient recruitment and regulatory oversight of study. NN manages participant engagement, assists with regulatory oversight, and researcher access. AP, KH, YK perform recruitment and enrollment of study participants.

Lab Operations

JoEllen Weaver, Meghan Livingstone, Fred Vadivieso, Stephanie DerOhannessian, Teo Tran, Julia Stephanowski, Monica Zielinski, Ned Haubein, Joseph Dunn

Contribution: JW, ML, FV, SD conduct oversight of lab operations. ML, FV, AK, SD, TT, JS, MZ perform sample processing. NH, JD are responsible for sample tracking and the laboratory information management system.

Clinical Informatics

Anurag Verma, Ph.D., Colleen Morse Kripke, M.S. DPT, MSA, Marjorie Risman, M.S., Renae Judy, B.S.

Contribution: All authors contributed to the development and validation of clinical phenotypes used to identify study subjects and (when applicable) controls.

Genome Informatics

Anurag Verma Ph.D., Shefali S. Verma, Ph.D., Yuki Bradford, M.S., Scott Dudek, M.S., Theodore Drivas, M.D., Ph.D.

Contribution: A.V., S.S.V. are responsible for the analysis, design, and infrastructure needed to quality control genotype and exome data. Y.B. performs the analysis. T.D. and A.V. provides variant and gene annotations and their functional interpretation of variants.

**Tables, Figures, and Figure Legends**

**Supplemental Table 3: Sum of single effects (SuSiE) colocalization results at select novel lead loci in the GWAS-by-subtraction results and aortic GTEx eQTLs.**

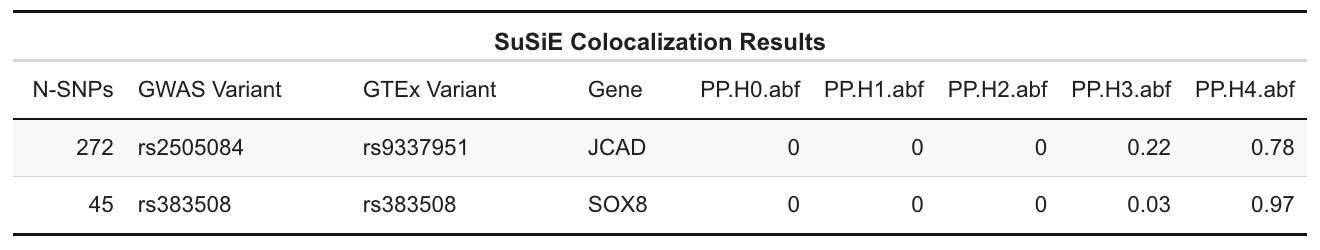

**Supplemental Table 4: Clinical characteristics of Penn Medicine Biobank individuals with and without thoracic aortic dissection who have at least 1 trans-thoracic echocardiography or computed tomography measurement of ascending aortic diameter.**

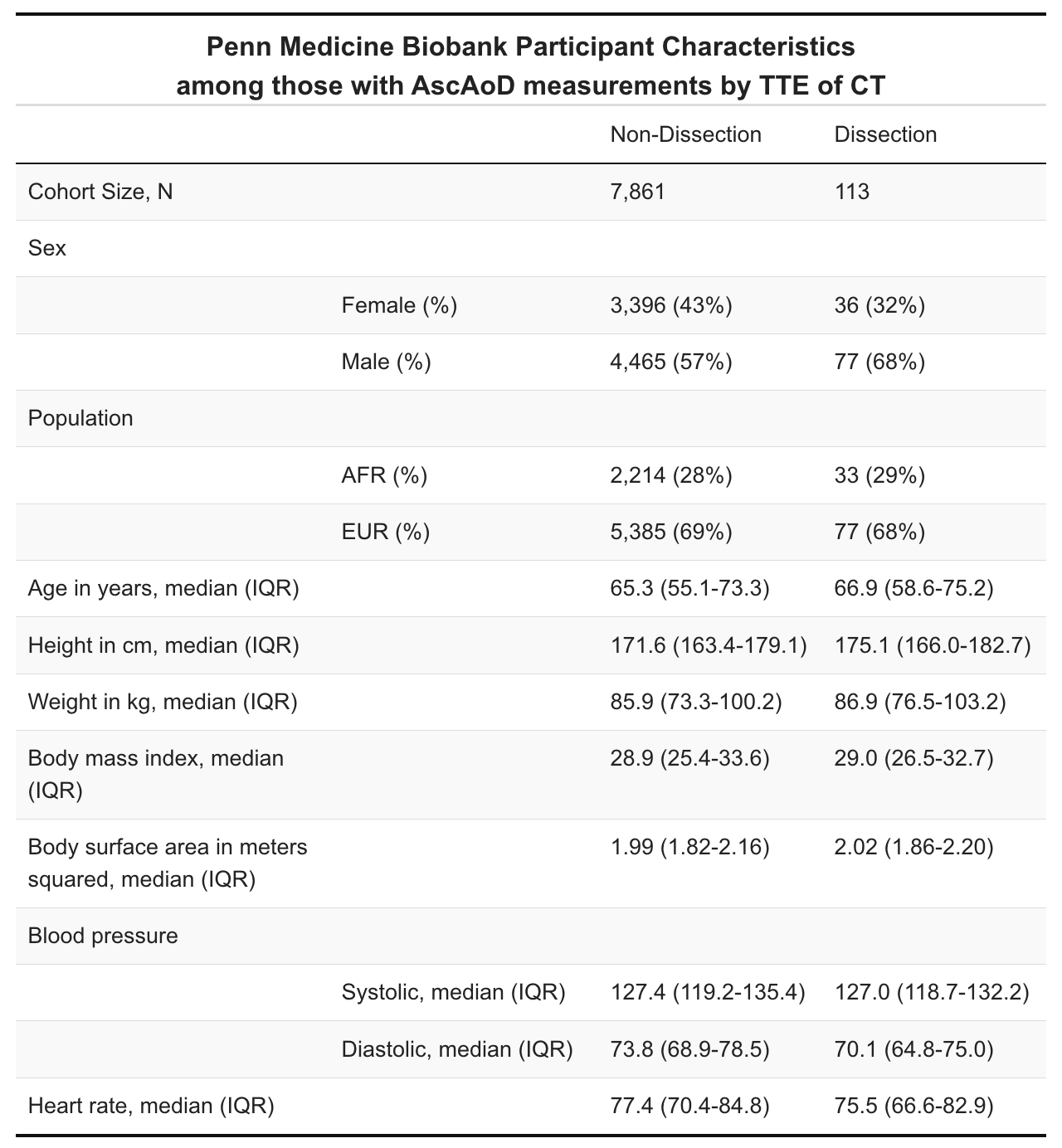

**Supplemental Table 5: Probability of cross-model practical differences in mean log-loss based on calculated region of practical equivalence between models with and without the Dissection-PRS. The reference model is listed in the first column, and to each reference model the Dissection-PRS was added to compare model log-loss change. The values in the three columns on the right are probabilities of either a practically negative, equivalent, or positive difference in log-loss between the two models being compared.**

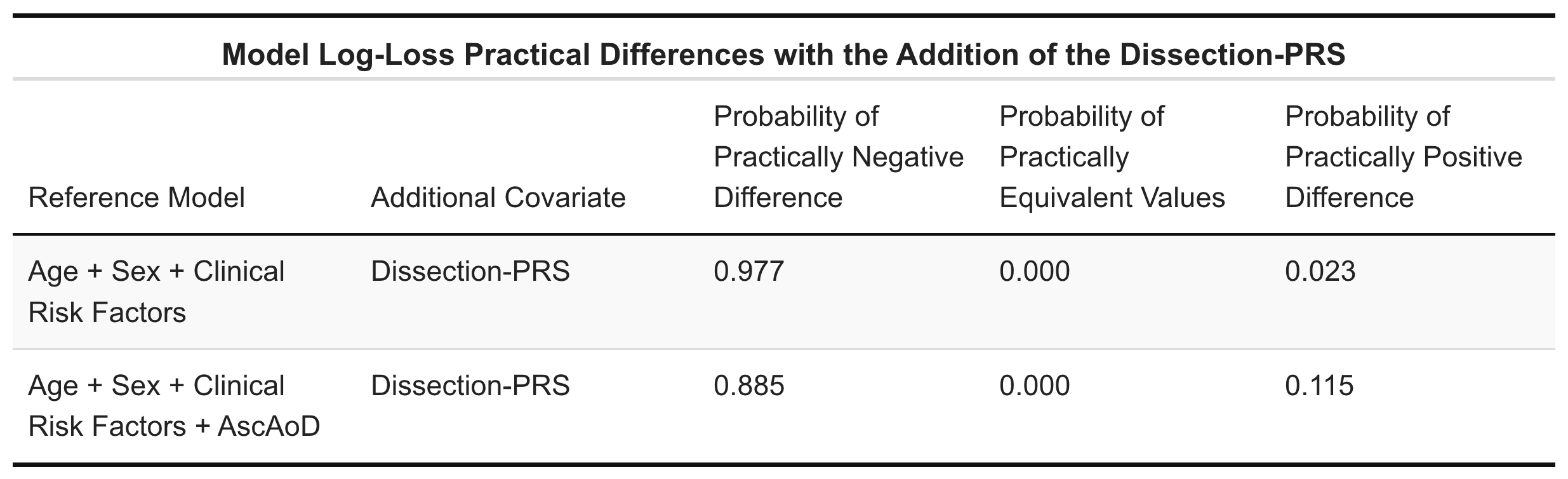

AscAoD = Ascending aortic diameter; Dissection-PRS = Thoracic aortic dissection polygenic risk score**.**

**Supplemental Table 6: Likelihood ratio test differences between models with and without the Dissection-PRS. The reference model is listed in the first column, and to each reference model the Dissection-PRS was added to compare model likelihood ratio change.**

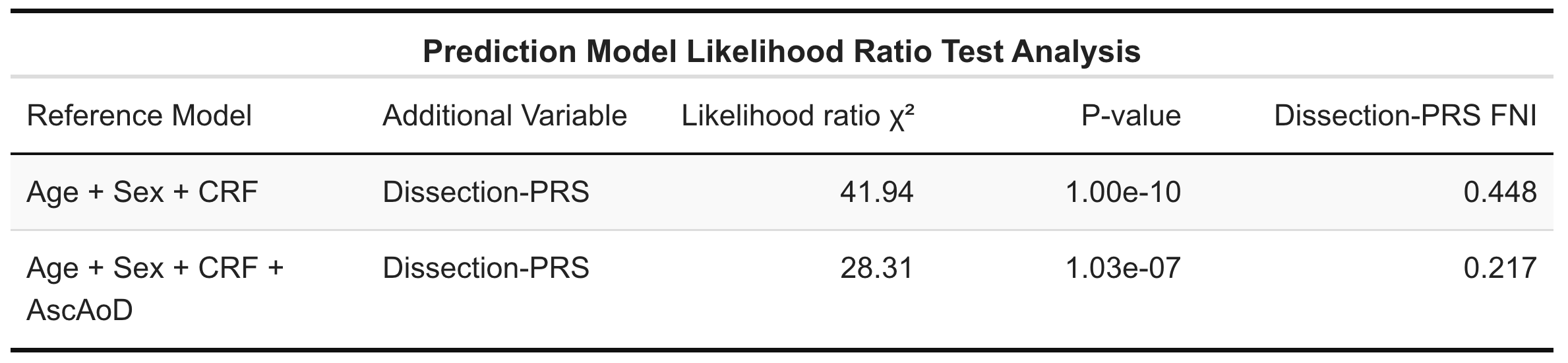

AscAoD = Ascending aortic diameter; CRF = Clinical Risk Factors; Dissection-PRS = Thoracic aortic dissection polygenic risk score; FNI = Fraction of new information.

**Supplemental Table 7: Probability of cross-model practical differences in mean log-loss based on calculated region of practical equivalence between models with and without the Dissection-PRS among females in the Penn Medicine Biobank. The reference model is listed in the first column, and to each reference model the Dissection-PRS was added to compare model log-loss change. The values in the three columns on the right are probabilities of either a practically negative, equivalent, or positive difference in log-loss between the two models being compared.**

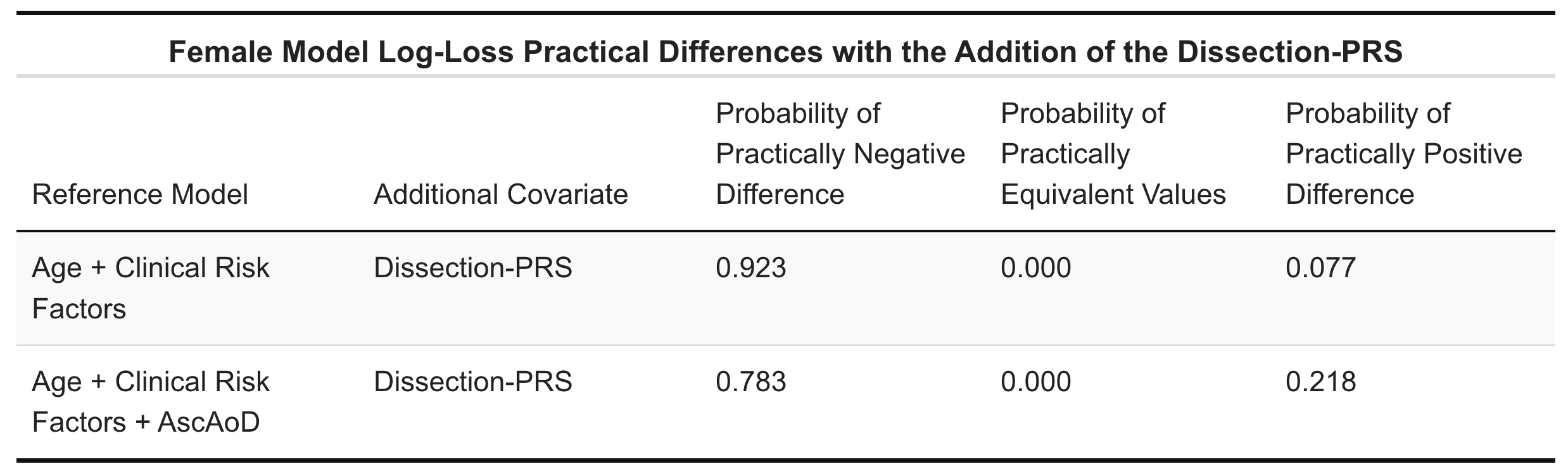

AscAoD = Ascending aortic diameter; Dissection-PRS = Thoracic aortic dissection polygenic risk score**.**

**Supplemental Table 8: Probability of cross-model practical differences in mean log-loss based on calculated region of practical equivalence between models with and without the Dissection-PRS among males in the Penn Medicine Biobank. The reference model is listed in the first column, and to each reference model the Dissection-PRS was added to compare model log-loss change. The values in the three columns on the right are probabilities of either a practically negative, equivalent, or positive difference in log-loss between the two models being compared.**

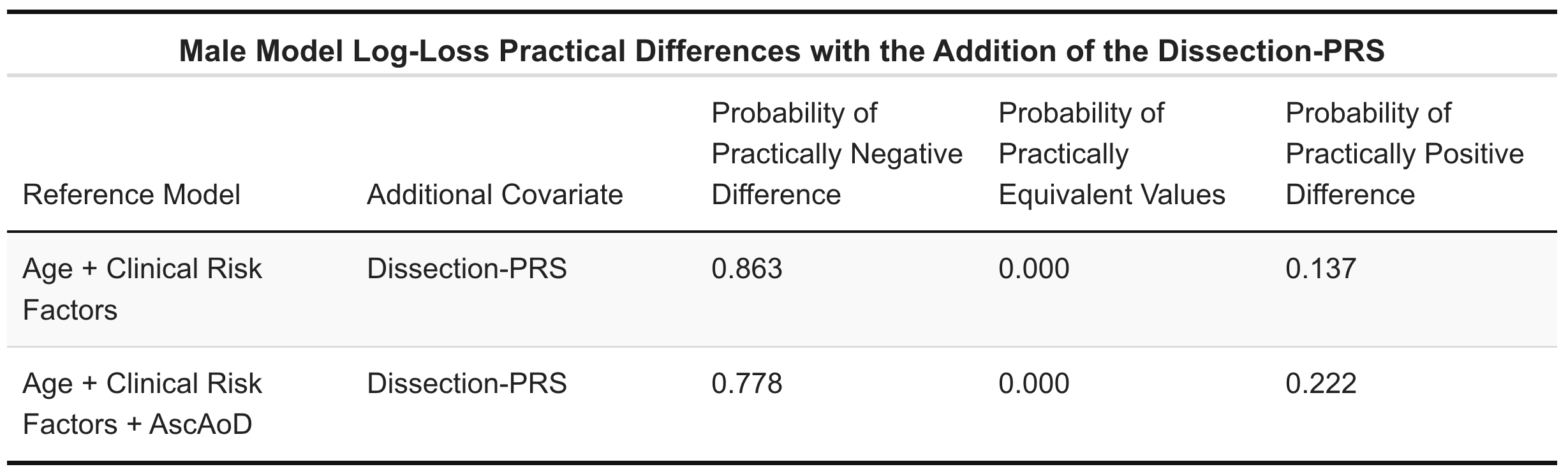

AscAoD = Ascending aortic diameter; Dissection-PRS = Thoracic aortic dissection polygenic risk score**.**

**Supplemental Table 9: Probability of cross-model practical differences in mean log-loss based on calculated region of practical equivalence between models with and without the Dissection-PRS among individuals genetically similar to the 1000G EUR reference population in the Penn Medicine Biobank. The reference model is listed in the first column, and to each reference model the Dissection-PRS was added to compare model log-loss change. The values in the three columns on the right are probabilities of either a practically negative, equivalent, or positive difference in log-loss between the two models being compared.**

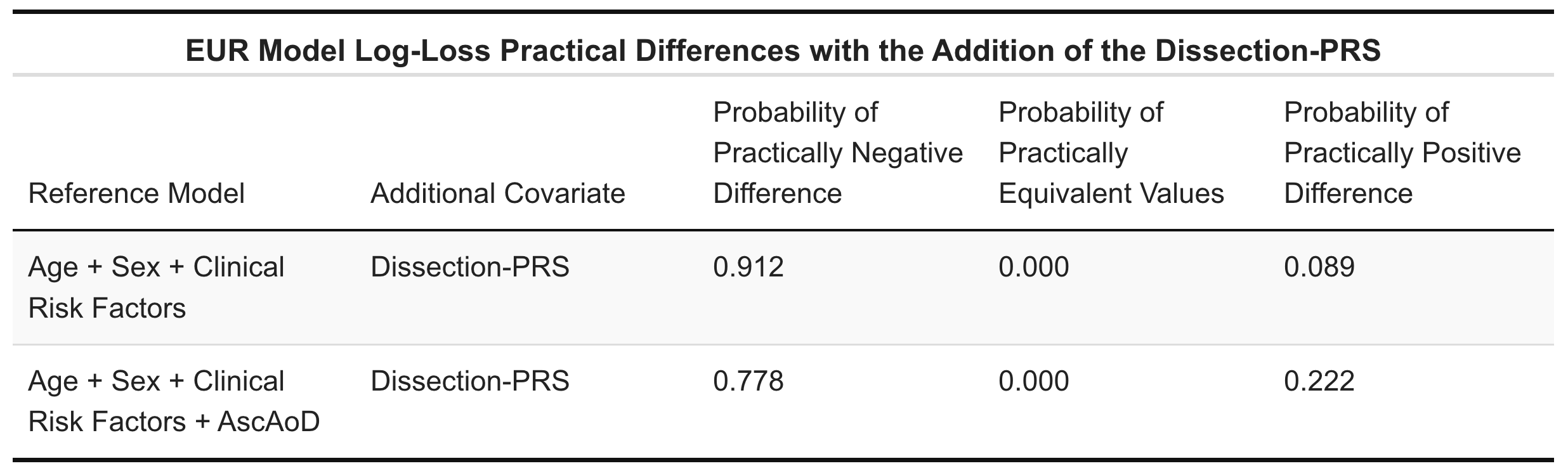

AscAoD = Ascending aortic diameter; Dissection-PRS = Thoracic aortic dissection polygenic risk score**.**

**Supplemental Table 10: Probability of cross-model practical differences in mean log-loss based on calculated region of practical equivalence between models with and without the Dissection-PRS among individuals genetically similar to the 1000G AFR reference population in the Penn Medicine Biobank. The reference model is listed in the first column, and to each reference model the Dissection-PRS was added to compare model log-loss change. The values in the three columns on the right are probabilities of either a practically negative, equivalent, or positive difference in log-loss between the two models being compared.**

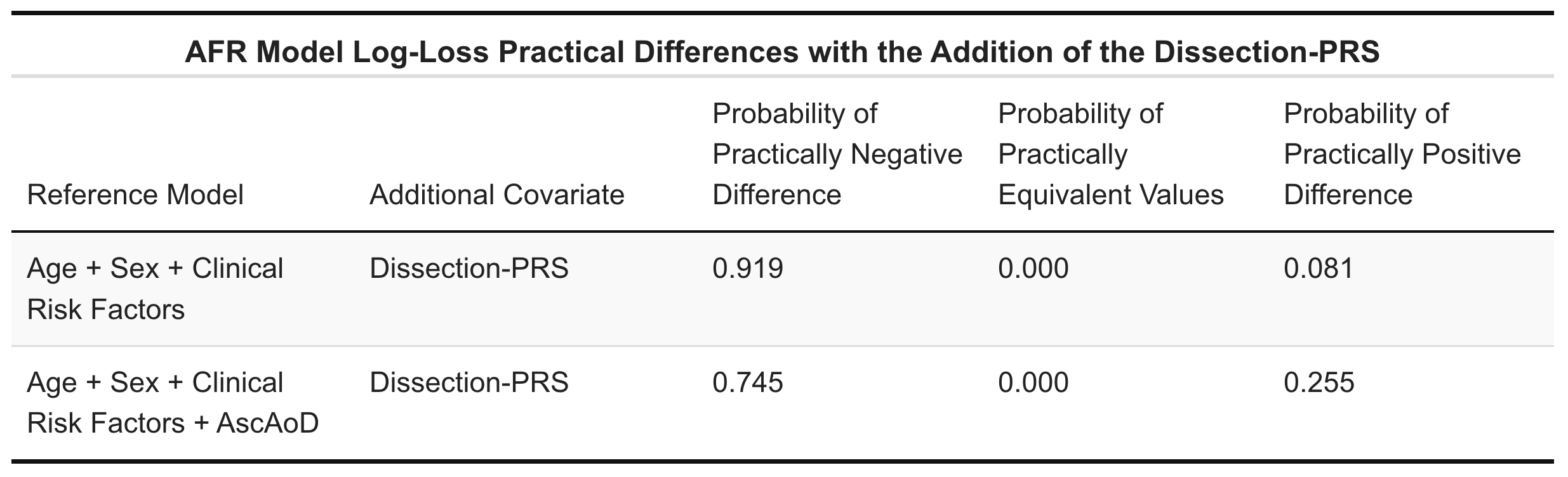

AscAoD = Ascending aortic diameter; Dissection-PRS = Thoracic aortic dissection polygenic risk score**.**

**Supplemental Table 11: Probability of cross-model practical differences in mean AUROC between models with and without the Dissection-PRS based on a calculated region of practical equivalence. The reference model is listed in the first column, and to each reference model the Dissection-PRS was added to compare model AUROC change. The values in the three columns on the right are probabilities of either a practically negative, equivalent, or positive difference in log-loss between the two models being compared.**

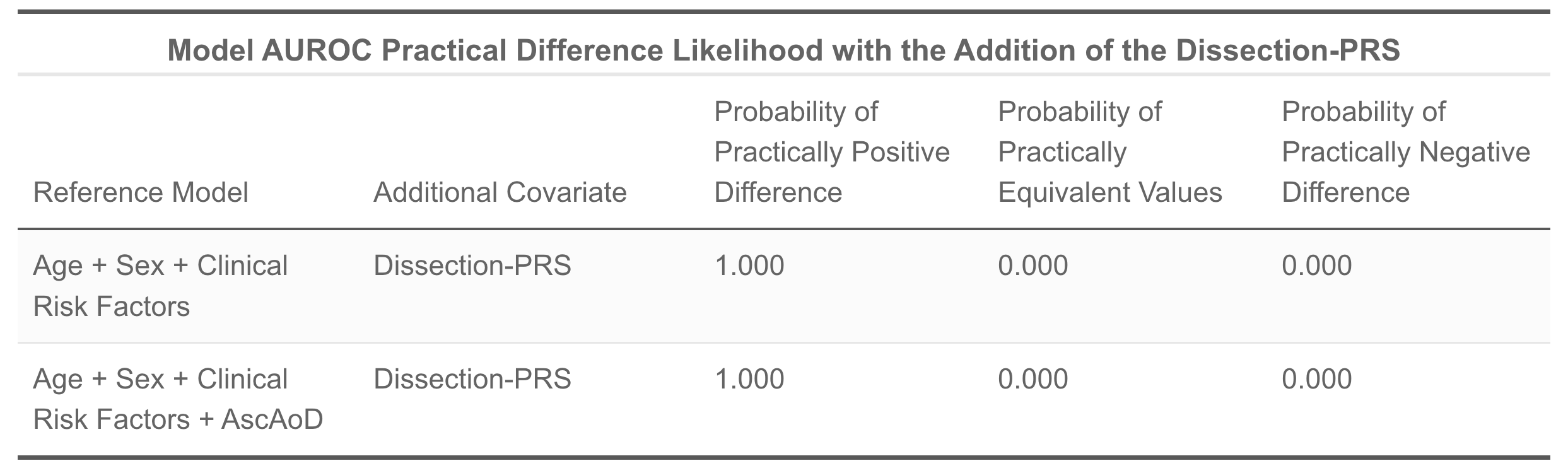

AscAoD = Ascending aortic diameter; AUROC = Are under the receiver operator characteristic curve; Dissection-PRS = Thoracic aortic dissection polygenic risk score**.**

**Supplemental Table 12: Area under the receiver operator characteristic curve differences between models with and without the Dissection-PRS using frequentist statistical analysis with the DeLong method.**

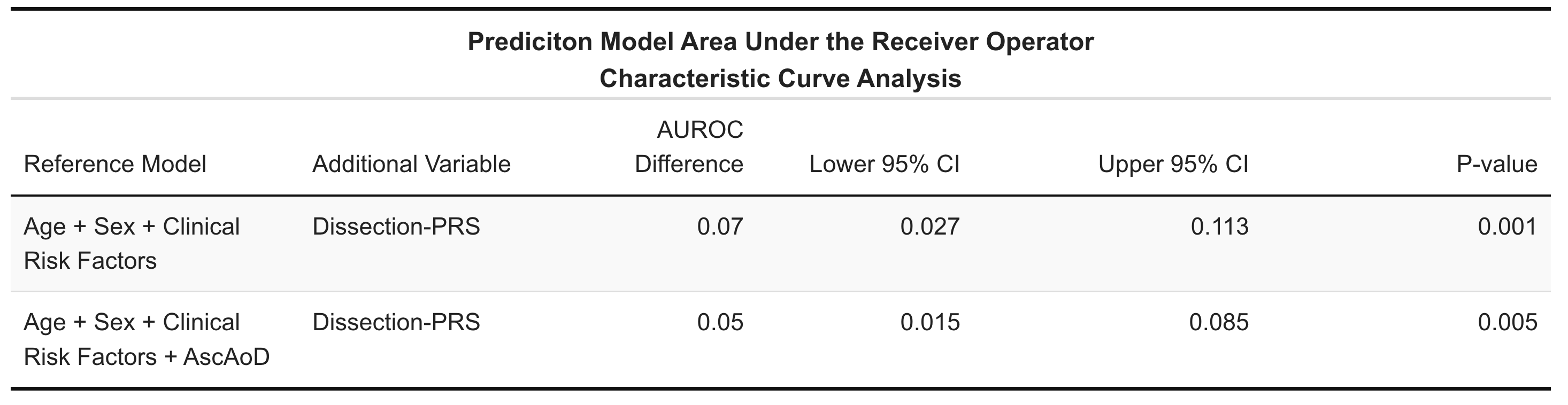

AscAoD = Ascending aortic diameter; Dissection-PRS = Thoracic aortic dissection polygenic risk score**.**

**Supplemental Table 13: Probability of cross-model practical differences in mean AUROC based on calculated region of practical equivalence between models with and without the Dissection-PRS among females in the Penn Medicine Biobank. The reference model is listed in the first column, and to each reference model the Dissection-PRS was added to compare model AUROC change. The values in the three columns on the right are probabilities of either a practically negative, equivalent, or positive difference in log-loss between the two models being compared.**

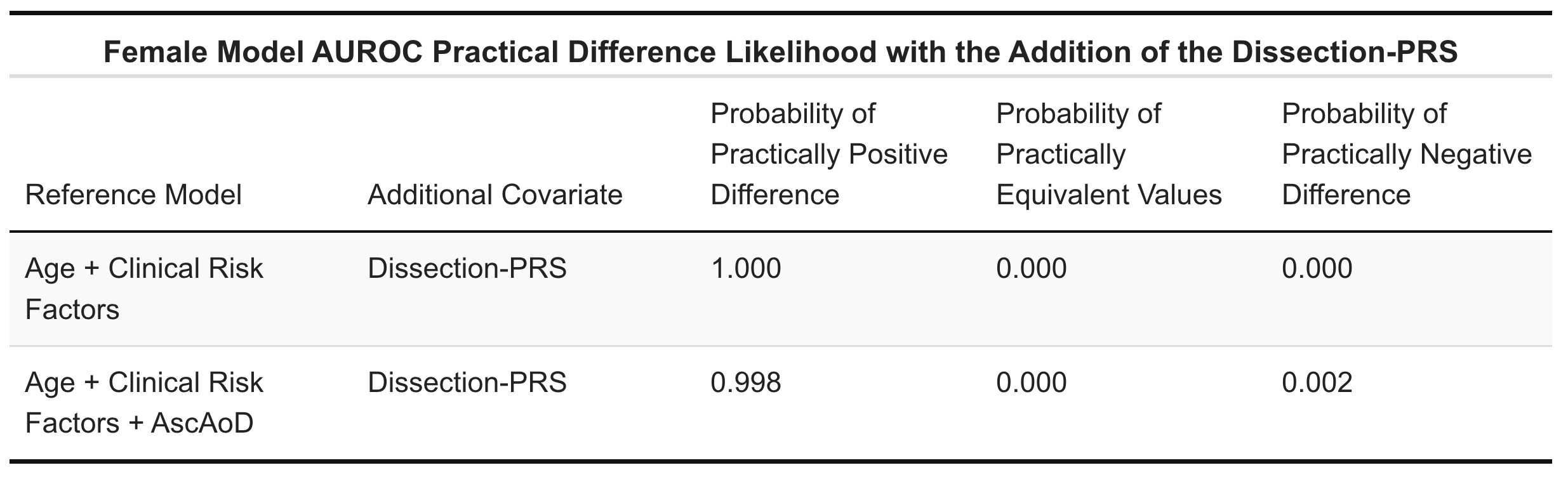

AscAoD = Ascending aortic diameter; AUROC = Are under the receiver operator characteristic curve; Dissection-PRS = Thoracic aortic dissection polygenic risk score**.**

**Supplemental Table 14: Probability of cross-model practical differences in mean AUROC based on calculated region of practical equivalence between models with and without the Dissection-PRS among males in the Penn Medicine Biobank. The reference model is listed in the first column, and to each reference model the Dissection-PRS was added to compare model AUROC change. The values in the three columns on the right are probabilities of either a practically negative, equivalent, or positive difference in log-loss between the two models being compared.**

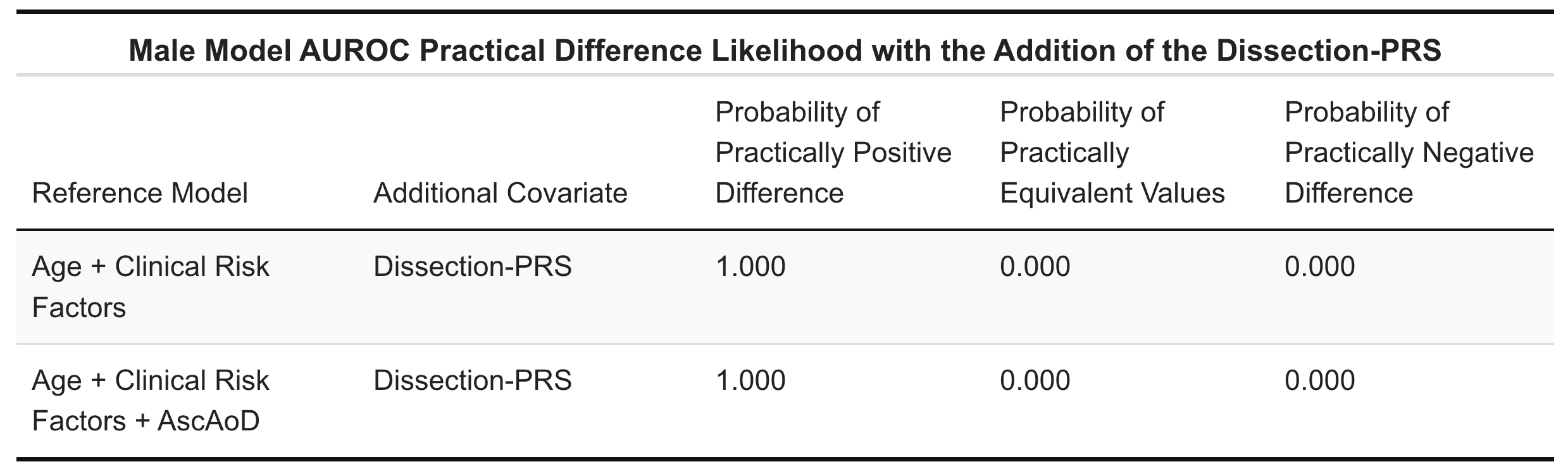

AscAoD = Ascending aortic diameter; AUROC = Are under the receiver operator characteristic curve; Dissection-PRS = Thoracic aortic dissection polygenic risk score**.**

**Supplemental Table 15: Probability of cross-model practical differences in mean AUROC based on calculated region of practical equivalence between models with and without the Dissection-PRS among individuals genetically similar to the 1000G EUR reference population in the Penn Medicine Biobank. The reference model is listed in the first column, and to each reference model the Dissection-PRS was added to compare model AUROC change. The values in the three columns on the right are probabilities of either a practically negative, equivalent, or positive difference in log-loss between the two models being compared.**

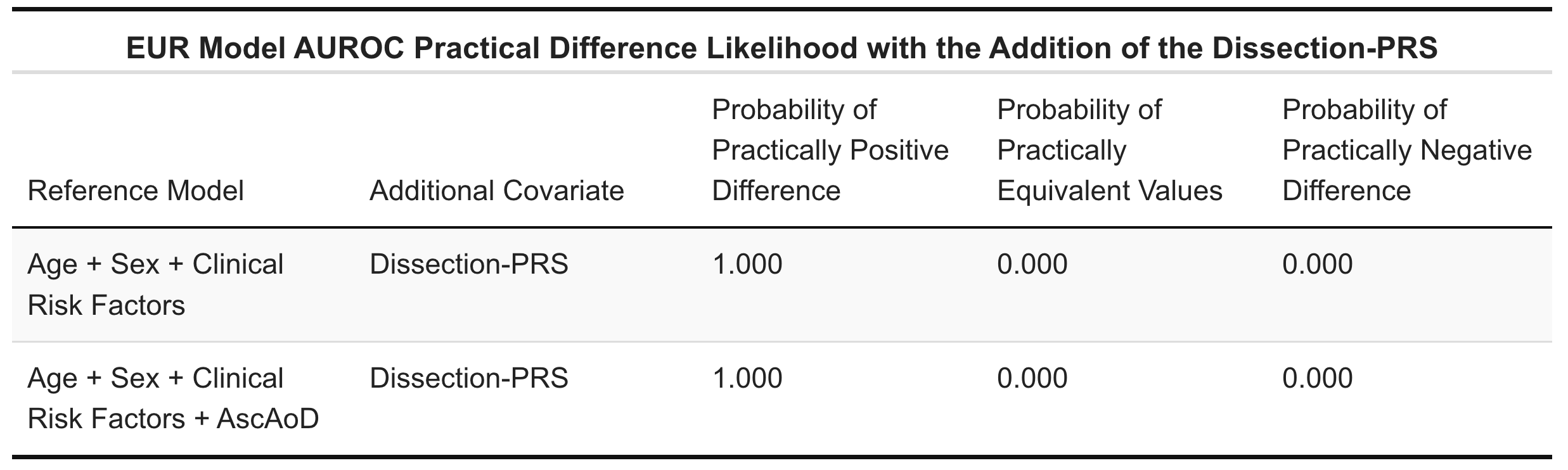

AscAoD = Ascending aortic diameter; AUROC = Are under the receiver operator characteristic curve; Dissection-PRS = Thoracic aortic dissection polygenic risk score**.**

**Supplemental Table 16: Probability of cross-model practical differences in mean AUROC based on calculated region of practical equivalence between models with and without the Dissection-PRS among individuals genetically similar to the 1000G AFR reference population in the Penn Medicine Biobank. The reference model is listed in the first column, and to each reference model the Dissection-PRS was added to compare model AUROC change. The values in the three columns on the right are probabilities of either a practically negative, equivalent, or positive difference in log-loss between the two models being compared.**

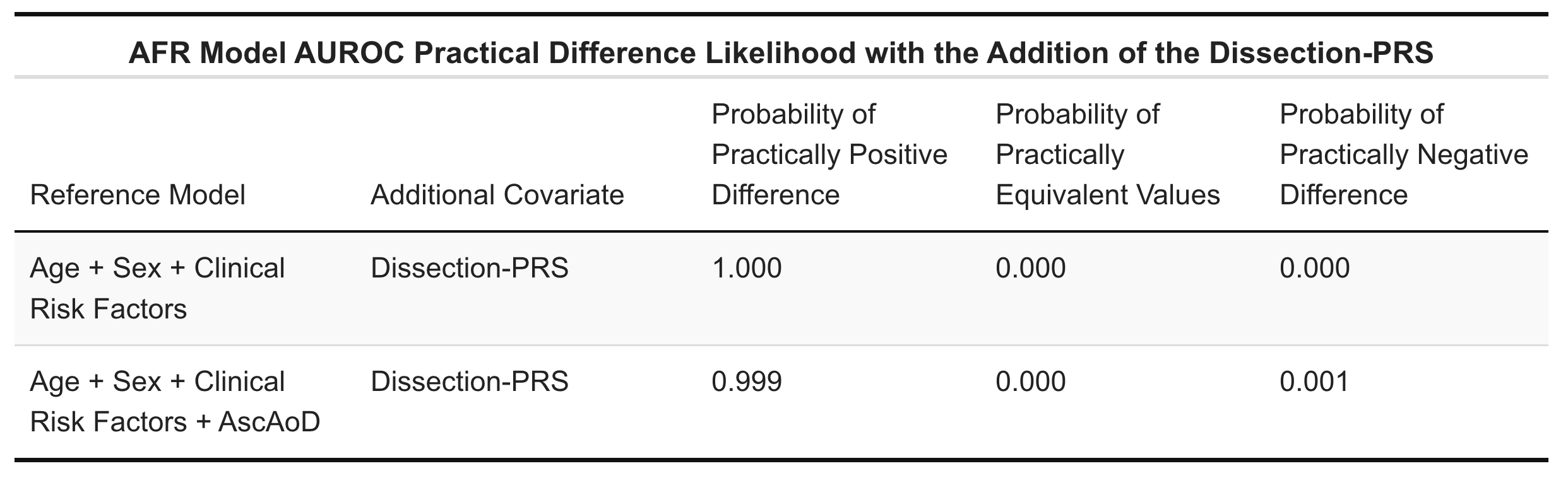

AscAoD = Ascending aortic diameter; AUROC = Are under the receiver operator characteristic curve; Dissection-PRS = Thoracic aortic dissection polygenic risk score**.**

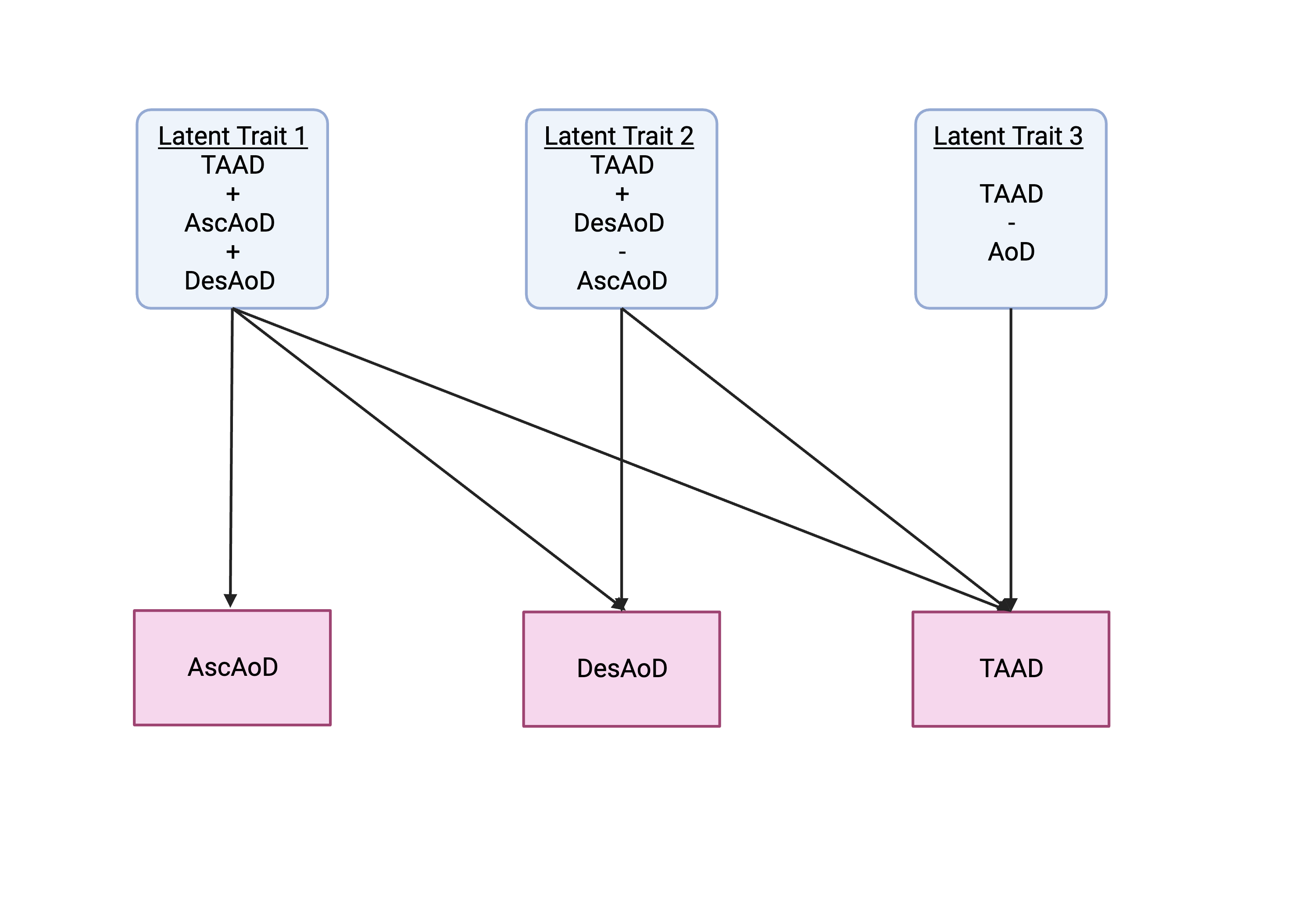
**Supplemental Figure 1: Cholesky decompensation model of genome-wide association study-by-subtraction of the novel latent TAAD-minus-AoD trait.** Multivariate Cholesky decompensation model with standardized paths of genetic influence. Measured traits with previously published GWAS are light red and latent traits are light blue. AscAoD = Ascending aortic diameter; AoD = Ascending and descending thoracic aortic diameter; DesAoD = Descending aortic diameter; TAAD = Thoracic aortic aneurysm and dissection.

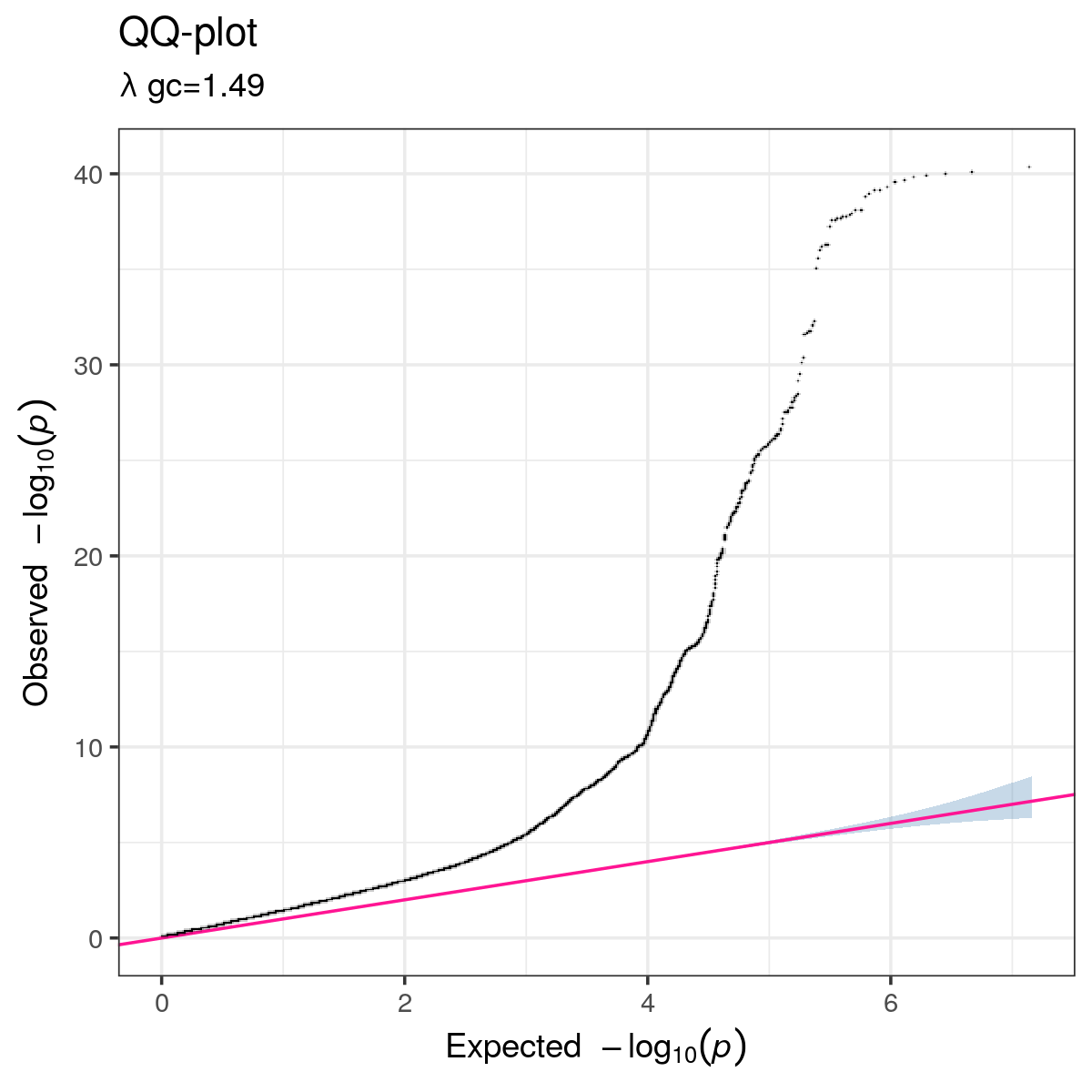

**Supplemental Figure 2: Quantile-quantile plot of *P*-value distribution.** The expected logistic regression association P-values versus the observed distribution of *P*-values for diameter-independent dissection association are displayed. No systematic inflation was observed (λGC = 1.49). All *P*-values were two-sided.

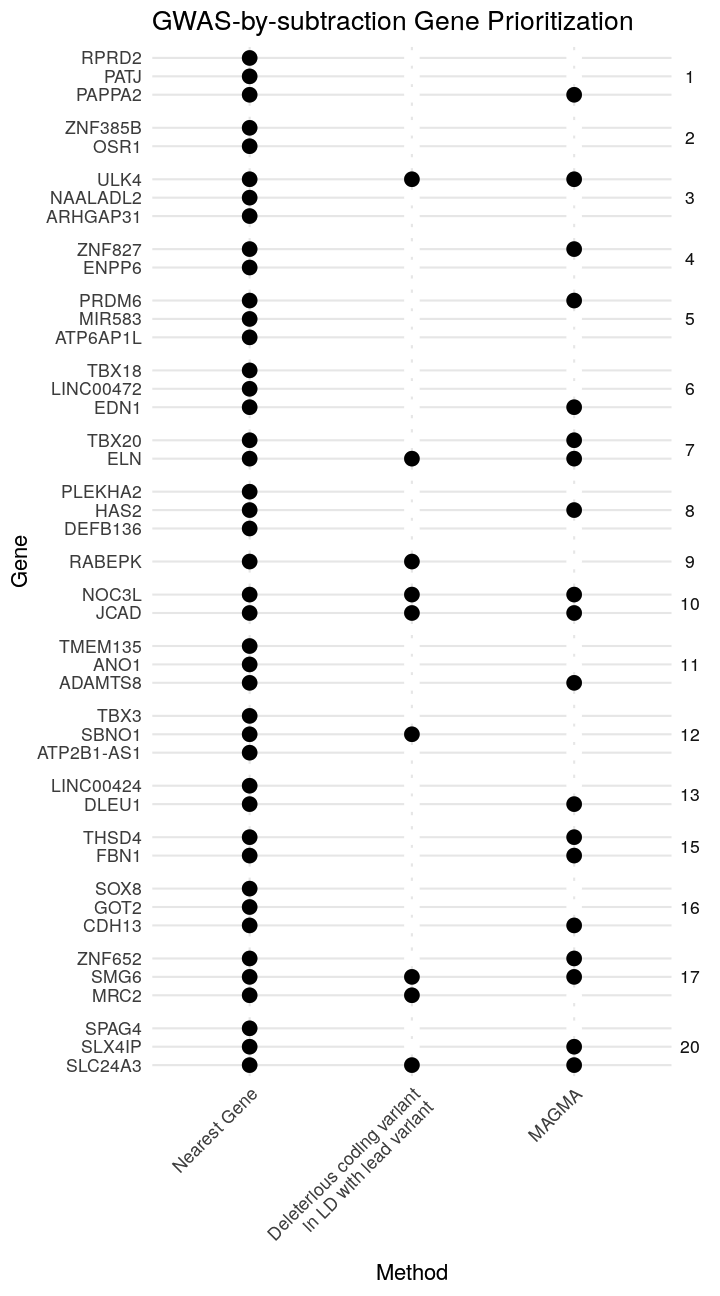

**Supplemental Figure 3: Genome-wide significant loci gene prioritization stratified by method with associated gene.** Lead variants identified in the GWAS-by-subtraction were mapped to the nearest gene. Gene prioritization was validated by identifiying genes in moderate (*R*^2^>0.3) or high (*R*^2^>0.8) linkage disequilibrium (LD) with coding variants in the prioritized genes, and using Multi-marker Analysis of GenoMic Annotation (MAGMA).

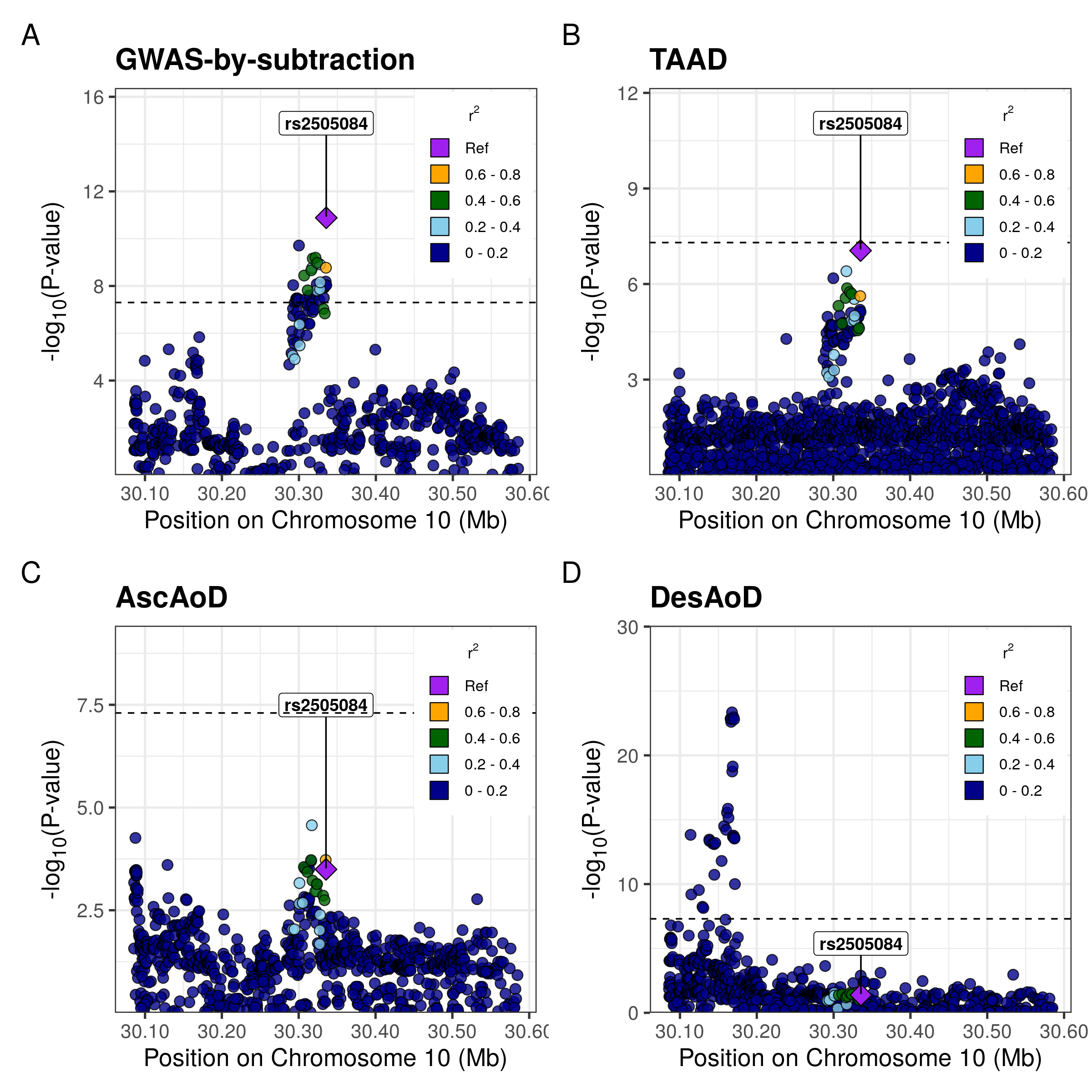

**Supplemental Figure 4: Regional Association Plots of the GWAS-by-subtraction Dissection GWAS as well as parent GWAS at the *JCAD* locus.** Regional association plots for independent genetic variants associated with dissection (A) at *P*<5x10^-8^ as well as the parent (B) TAAD, (C) AscAoD, and (D) DesAoD GWAS. Points are colored by linkage disequilibrium with index variant from the GWAS-by-subtraction estimated from the 1000G Phase 3 referece panel.

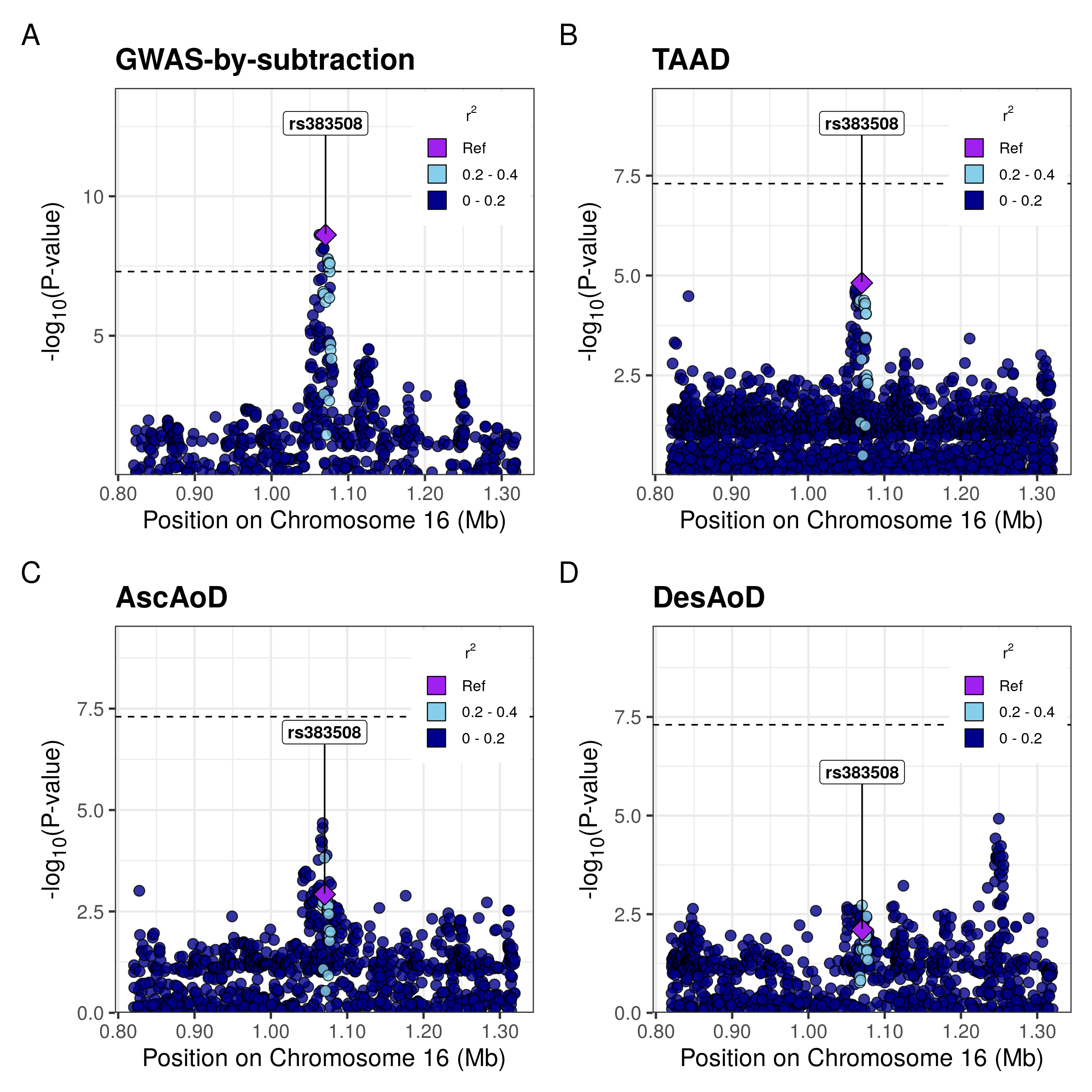

**Supplemental Figure 5: Regional Association Plots of the GWAS-by-subtraction Dissection GWAS as well as parent GWAS at the *SOX8* locus.** Regional association plots for independent genetic variants associated with dissection (A) at *P*<5x10^-8^ as well as the parent (B) TAAD, (C) AscAoD, and (D) DesAoD GWAS. Points are colored by linkage disequilibrium with index variant from the GWAS-by-subtraction estimated from the 1000G Phase 3 referece panel.

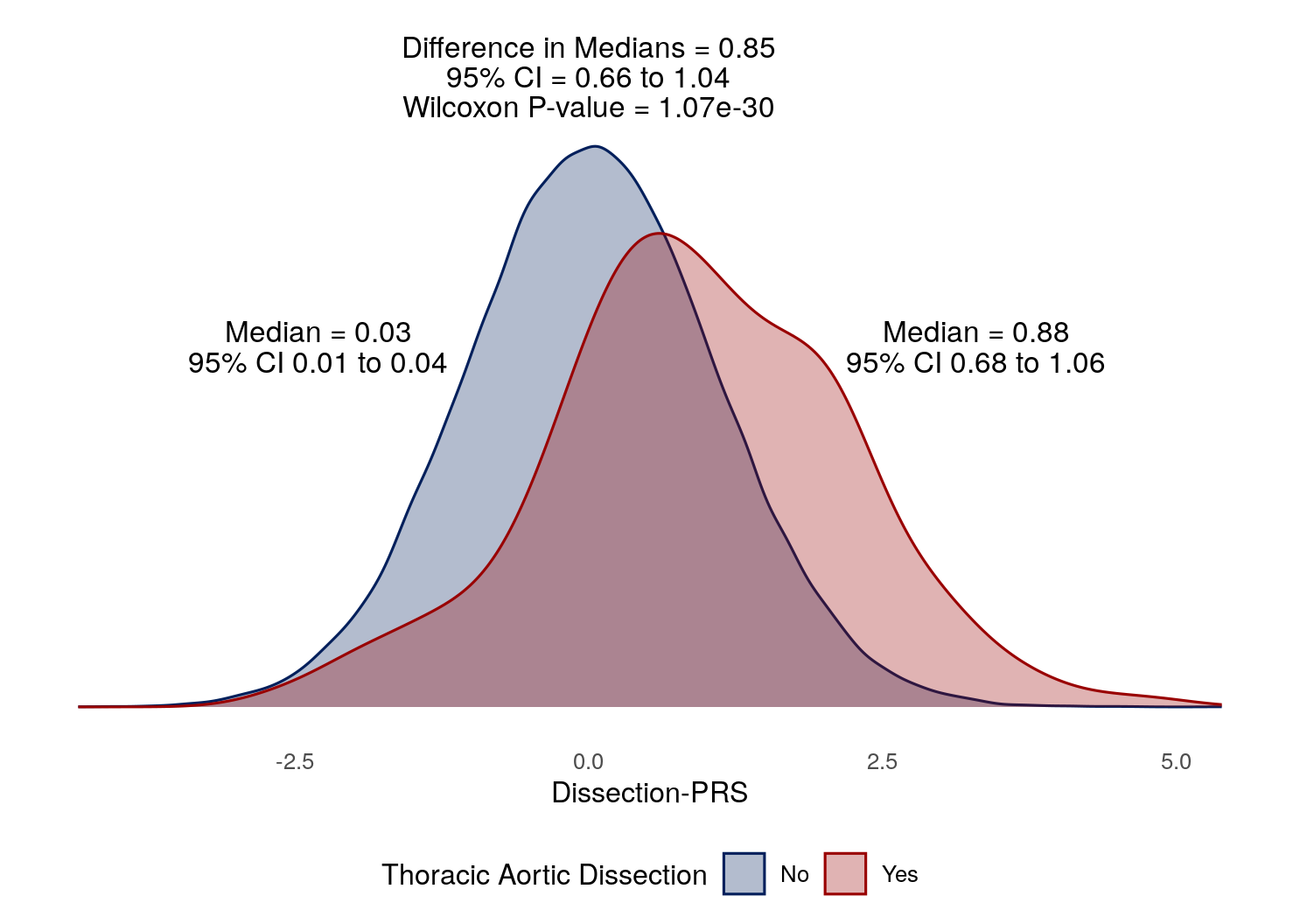

**Supplemental Figure 6: Difference in median Dissection-PRS between individuals with and without a diagnosis of thoracic aortic dissection**. Density plot of Dissection-PRS value of individuals with (red) and without (blue) a diagnosis of dissection. Difference in median PRS values above the density plots. CI = confidence interval.

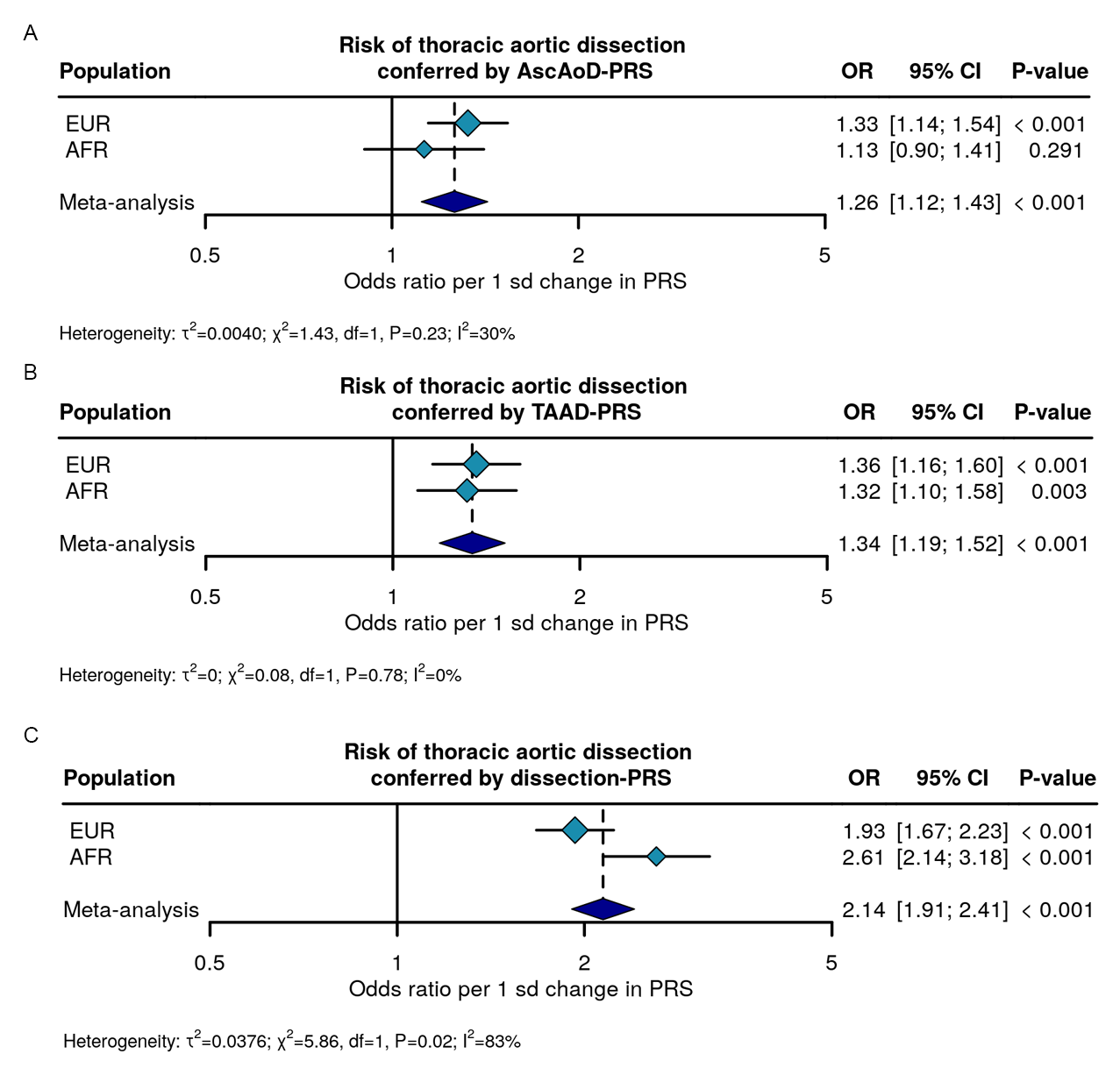

**Supplemental Figure 7: Association of different PRS with prevalent thoracic aortic dissection in the Penn Medicine Biobank stratified by genetically similar population group.** Multivariable logistic regression analysis of the AscAoD-PRS, TAAD-PRS, and Dissection-PRS among individuals in the PMBB stratified by genetically similar population group according to 1000G reference populations, adjusting for age, sex, and the first five genetic principal components. AscAoD-PRS = Ascending aortic diameter polygenic risk score; CI = confidence interval; OR = Odds ratio; TAAD-PRS = thoracic aortic aneurysm and dissection polygenic risk score; Dissection-PRS = thoracic aortic dissection polygenic risk score.

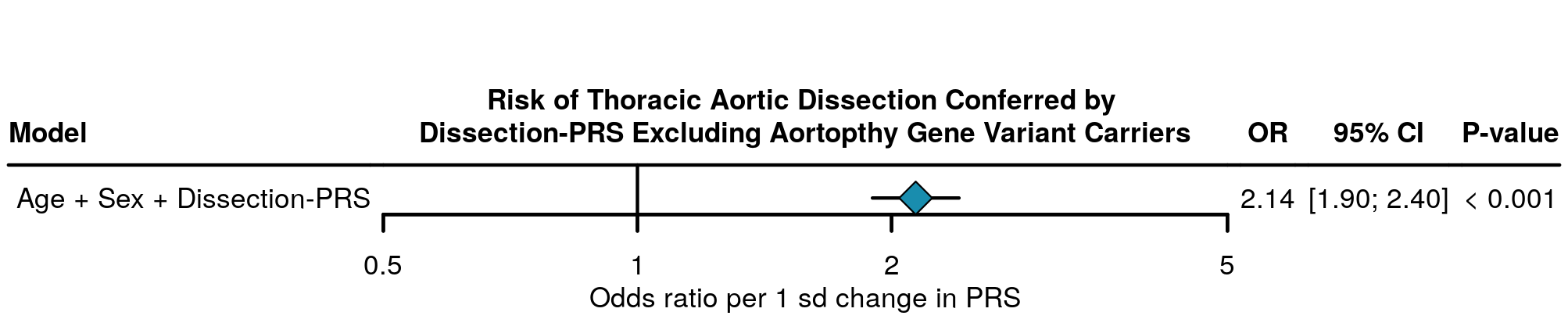

**Supplemental Figure 8: Association of the Dissection-PRS with prevalent thoracic aortic dissection in the Penn Medicine Biobank excluding individuals carrying a pathogenic or likely pathogenic variant in an aortopathy gene.** Multivariable logistic regression analysis of the risk for thoracic aortic dissection conferred by the Dissection-PRS adjusting for age, sex, and the first five genetic principal components. CI = confidence interval; OR = Odds ratio; Dissection-PRS = thoracic aortic dissection polygenic risk score.

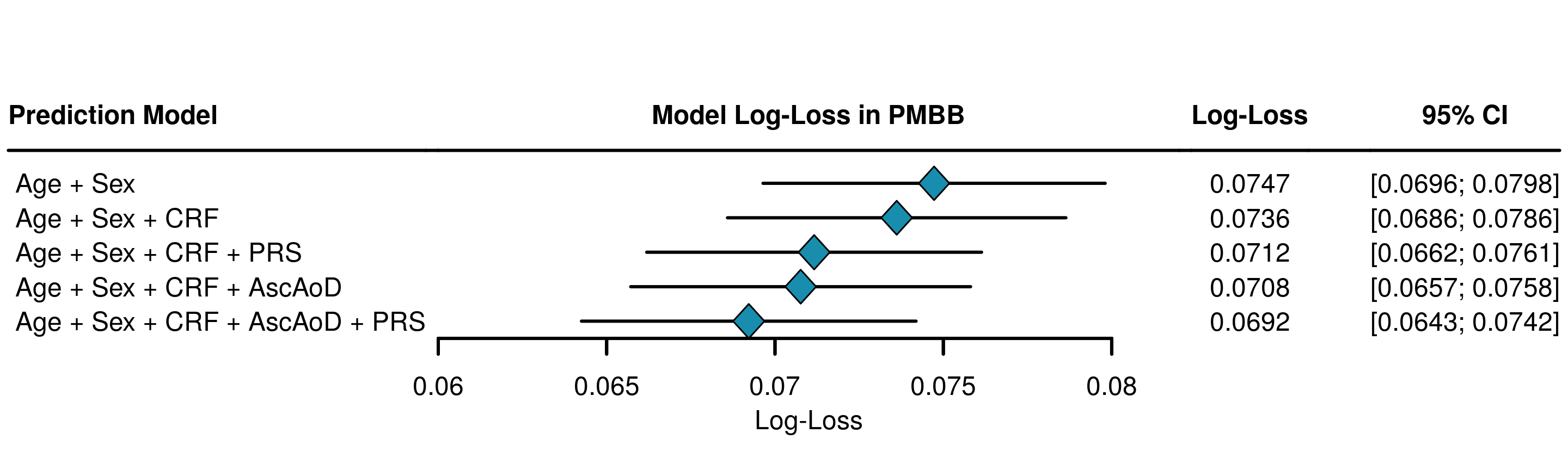

**Supplemental Figure 9: Log-loss comparison of logistic regression models to predict thoracic aortic dissection in the Penn Medicine Biobank.** Five-times 10-fold cross-validation analysis of model log-loss for each of Age + Sex, Age + Sex + Clinical Risk Factors, Age + Sex + Clinical Risk Factors + Dissection-PRS, Age + Sex + Clinical Risk Factors + AscAoD, and Age + Sex + Clinical Risk Factors + AscAoD + Dissection-PRS. Corresponding error bars demonstrating 95% confidence intervals for models utilized among all individuals with AscAoD measured by TTE. AscAoD = ascending thoracic aortic diameter; CI = 95% confidence interval; CRF = Clinical risk factors; PRS = thoracic aortic dissection polygenic risk score.

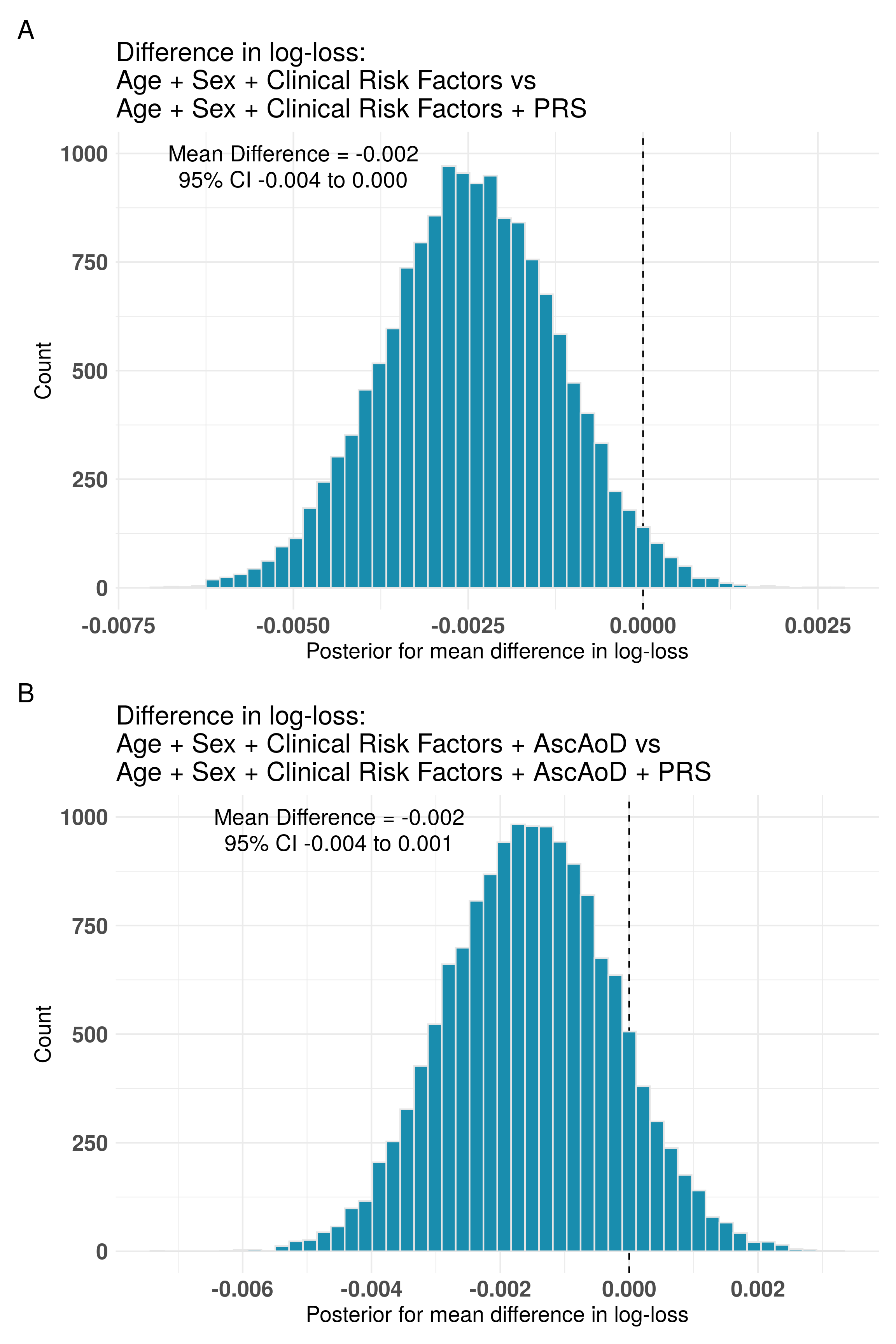

**Supplemental Figure 10: Cross-model Bayesian comparison of difference in log-loss between models with and without the inclusion of the Dissection-PRS among individuals in PMBB.** Posterior distribution for the difference in log-loss between (A) Age + Sex + Clinical Risk Factors and Age + Sex + Clinical Risk Factors + Dissection-PRS; and (B) Age + Sex + Clinical Risk Factors + AscAoD and Age + Sex + Clinical Risk Factors + AscAoD + Dissection-PRS. Between-model mean difference and 95% credible intervals in each plot. AscAoD = ascending thoracic aortic diameter; PRS = thoracic aortic dissection polygenic risk score; CI = credible interval.

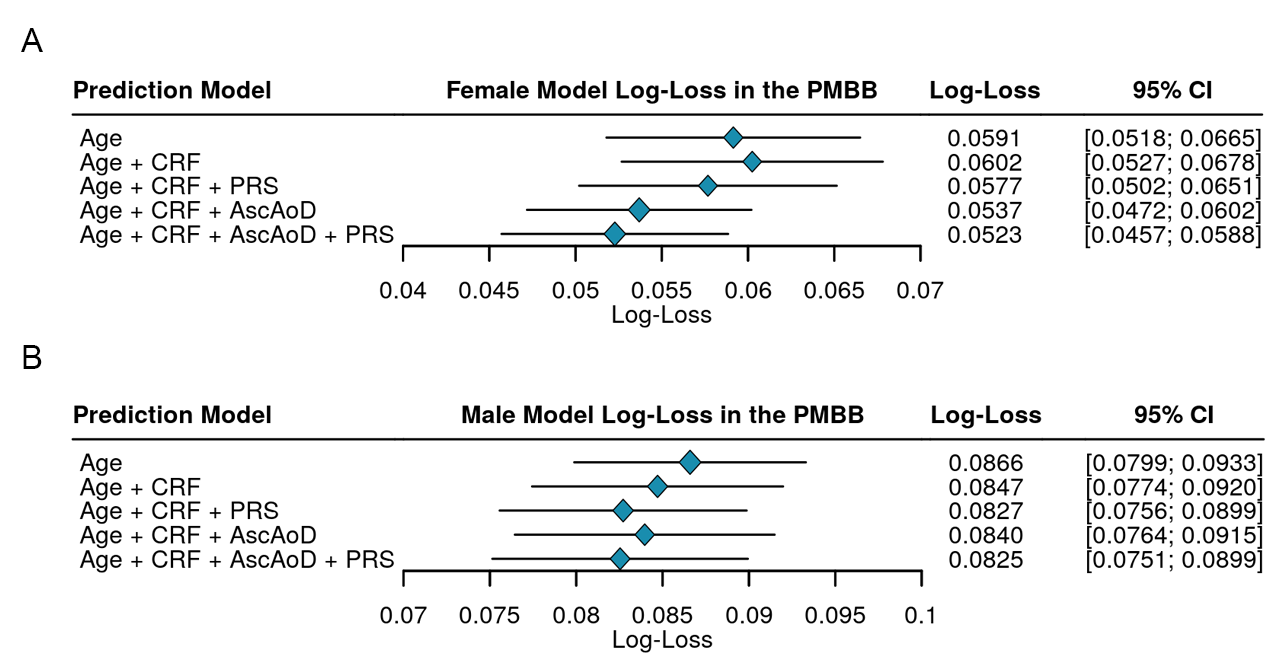

**Supplemental Figure 11: Log-loss comparison of logistic regression models to predict thoracic aortic dissection stratified by sex.** Female cohort (A) and male cohort (B) five-times 10-fold cross-validation analysis of model log-loss for each of five-times 10-fold cross-validation analysis of model log-loss for each of Age, Age + Clinical Risk Factors, Age + Clinical Risk Factors + Dissection-PRS, Age + Clinical Risk Factors + AscAoD, and Age + Clinical Risk Factors + AscAoD + Dissection-PRS. Corresponding error bars demonstrating 95% confidence intervals for models utilized among all individuals with AscAoD measured by TTE. AscAoD = ascending thoracic aortic diameter; CI = 95% confidence interval; CRF = Clinical risk factors; PRS = thoracic aortic dissection polygenic risk score.

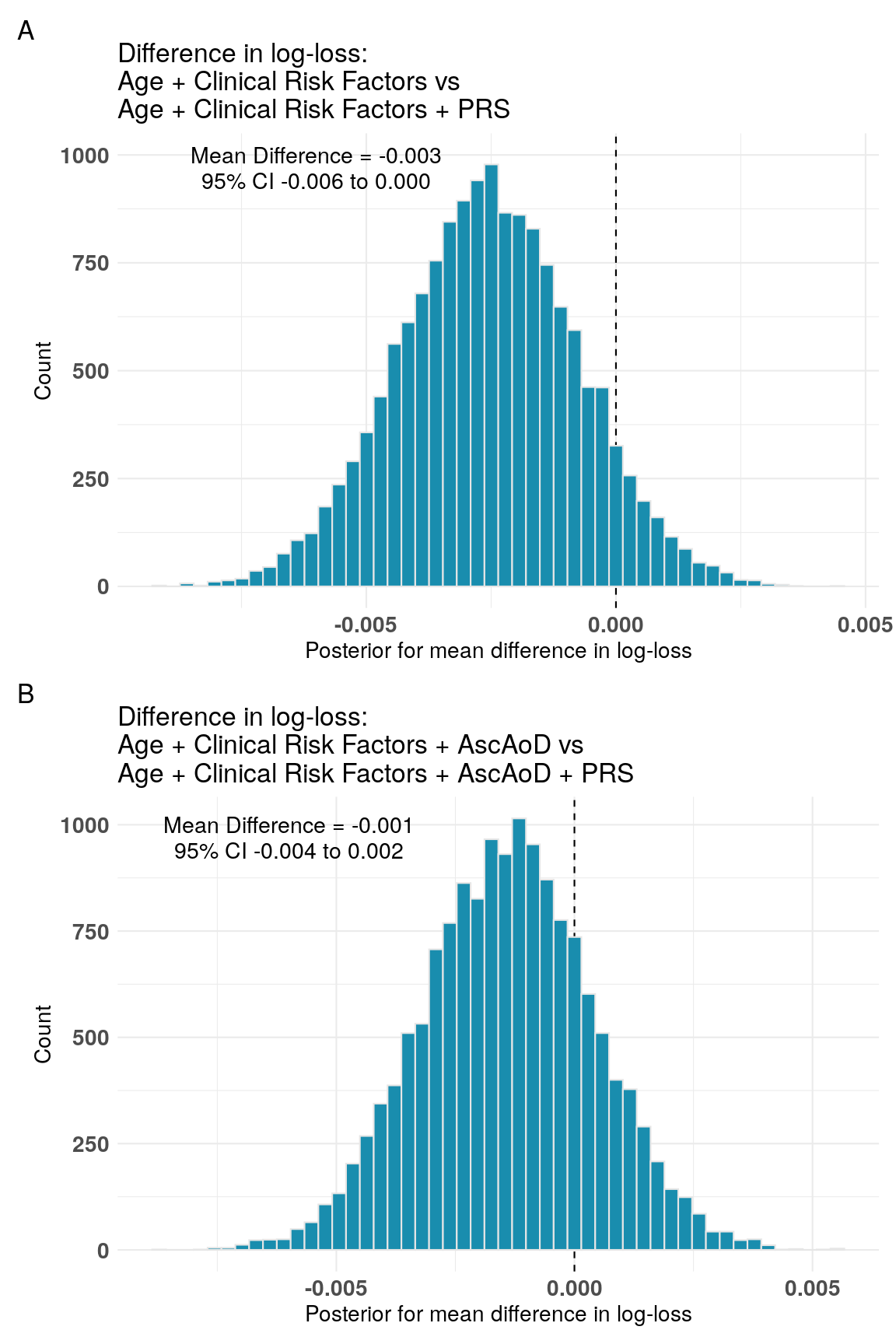

**Supplemental Figure 12: Cross-model Bayesian comparison of difference in log-loss between models with and without the inclusion of the Dissection-PRS among females in PMBB.** Posterior distribution for the difference in log-loss between (A) Age + Clinical Risk Factors and Age + Clinical Risk Factors + Dissection-PRS; and (B) Age + Clinical Risk Factors + AscAoD and Age + Clinical Risk Factors + AscAoD + Dissection-PRS. Between-model mean difference and 95% credible intervals in each plot. AscAoD = ascending thoracic aortic diameter; PRS = thoracic aortic dissection polygenic risk score; CI = credible interval.

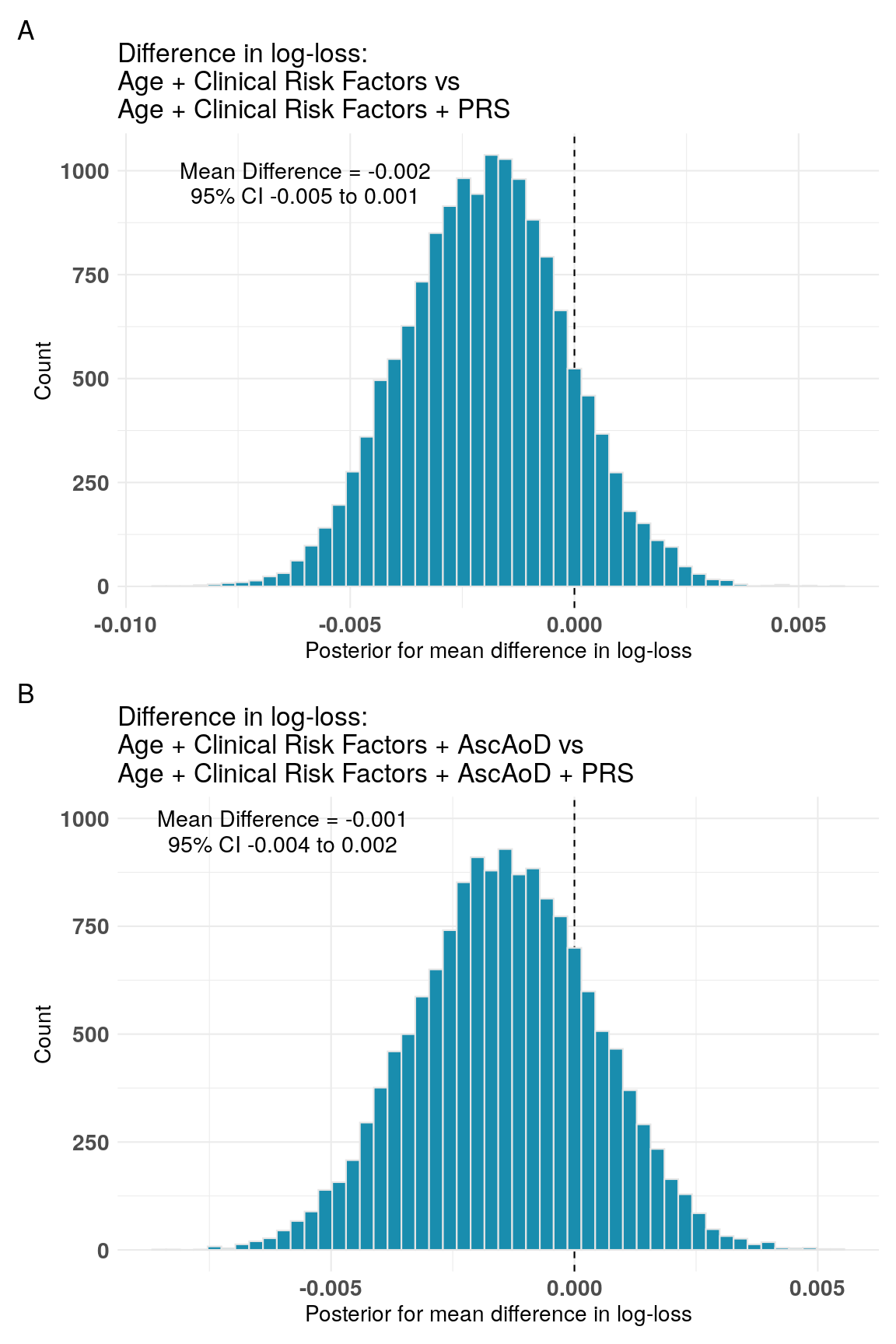

**Supplemental Figure 13: Cross-model Bayesian comparison of difference in log-loss between models with and without the inclusion of the Dissection-PRS among males in PMBB.** Posterior distribution for the difference in log-loss between (A) Age + Clinical Risk Factors and Age + Clinical Risk Factors + Dissection-PRS; and (B) Age + Clinical Risk Factors + AscAoD and Age + Clinical Risk Factors + AscAoD + Dissection-PRS. Between-model mean difference and 95% credible intervals in each plot. AscAoD = ascending thoracic aortic diameter; PRS = thoracic aortic dissection polygenic risk score; CI = credible interval.

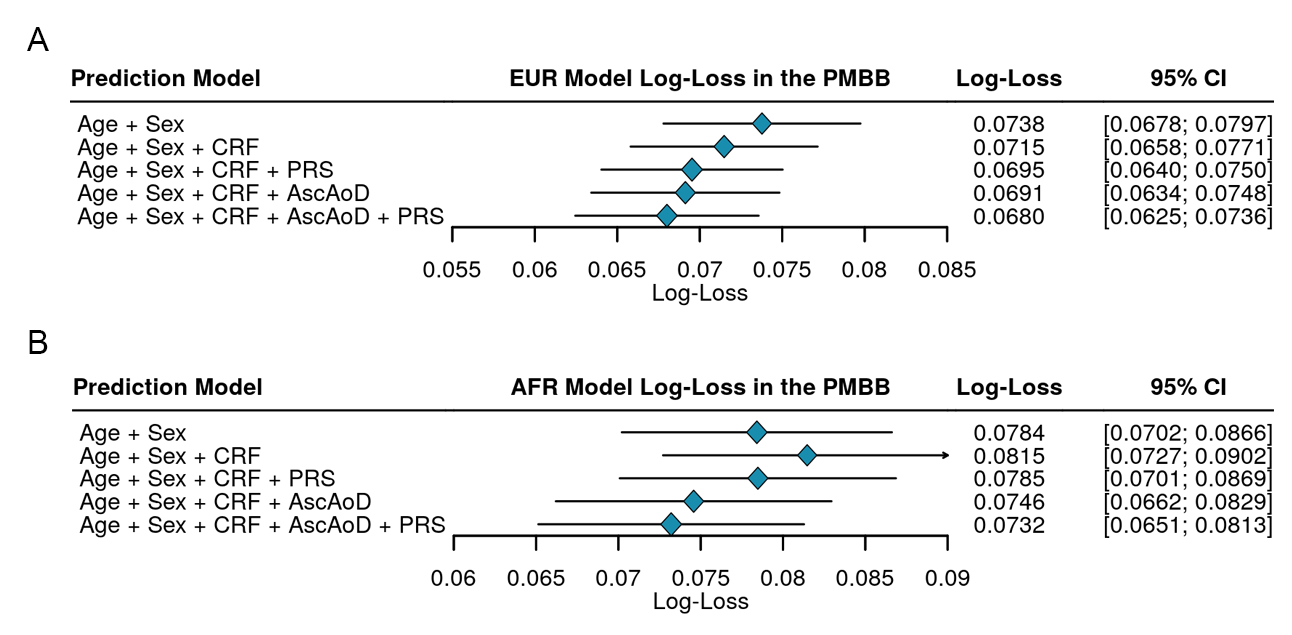

**Supplemental Figure 14: Log-loss comparison of logistic regression models to predict thoracic aortic dissection stratified by population group.** EUR cohort (A) and AFR cohort (B) five-times 10-fold cross-validation analysis of model log-loss for each of Age + Sex, Age + Sex + Clinical Risk Factors, Age + Sex + Clinical Risk Factors + Dissection-PRS, Age + Sex + Clinical Risk Factors + AscAoD, and Age + Sex + Clinical Risk Factors + AscAoD + Dissection-PRS. Corresponding error bars demonstrating 95% confidence intervals for models utilized among all individuals with AscAoD measured by TTE or computed tomography. AscAoD = ascending thoracic aortic diameter; CI = 95% confidence interval; CRF = Clinical risk factors; PRS = thoracic aortic dissection polygenic risk score.

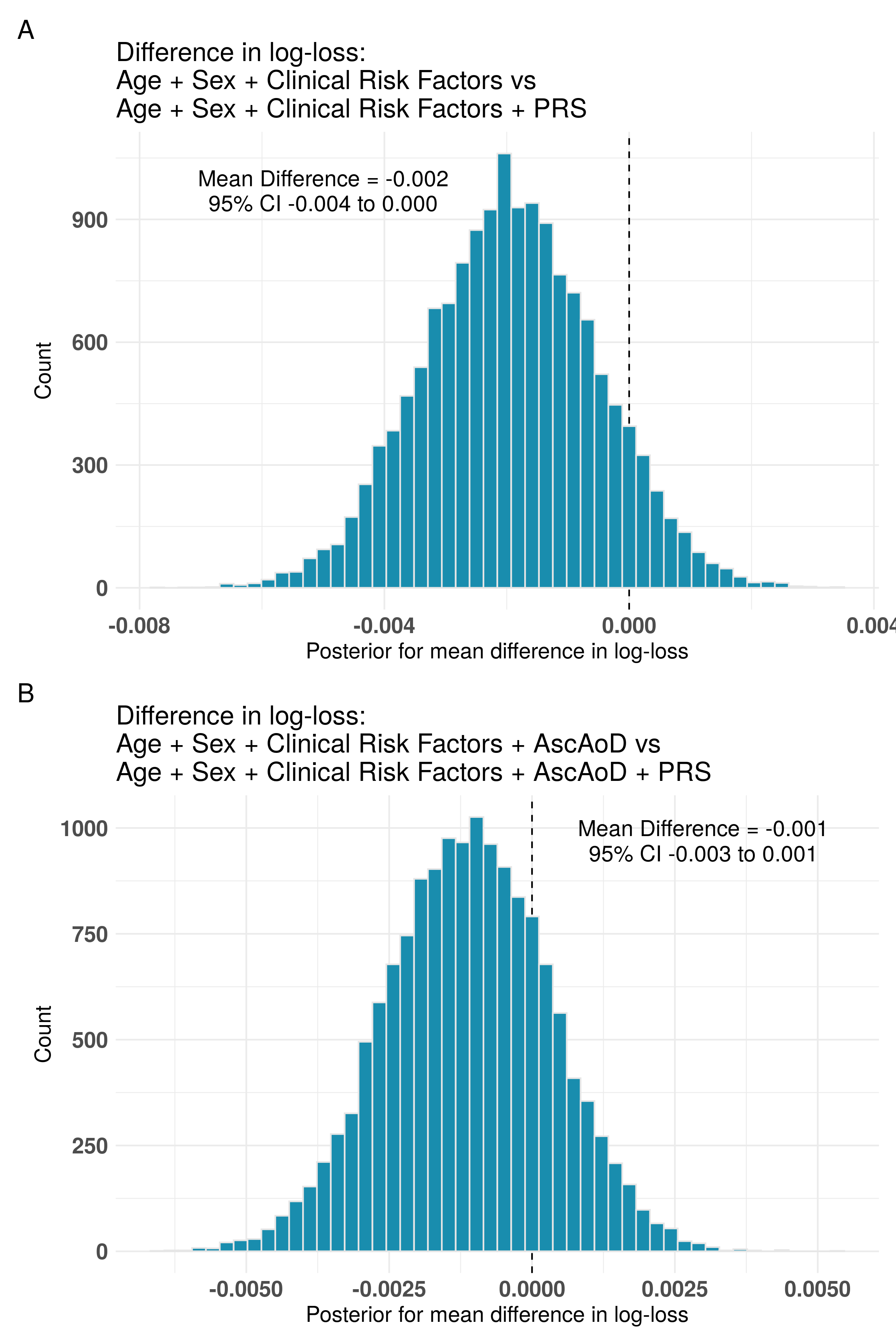

**Supplemental Figure 15: Cross-model Bayesian comparison of difference in log-loss between models with and without the inclusion of the Dissection-PRS among individuals genetically similar to the 1000G EUR reference population in PMBB.** Posterior distribution for the difference in log-loss between (A) Age + Sex + Clinical Risk Factors and Age + Sex + Clinical Risk Factors + Dissection-PRS; and (B) Age + Sex + Clinical Risk Factors + AscAoD and Age + Sex + Clinical Risk Factors + AscAoD + Dissection-PRS. Between-model mean difference and 95% credible intervals in each plot. AscAoD = ascending thoracic aortic diameter; PRS = thoracic aortic dissection polygenic risk score; CI = credible interval.

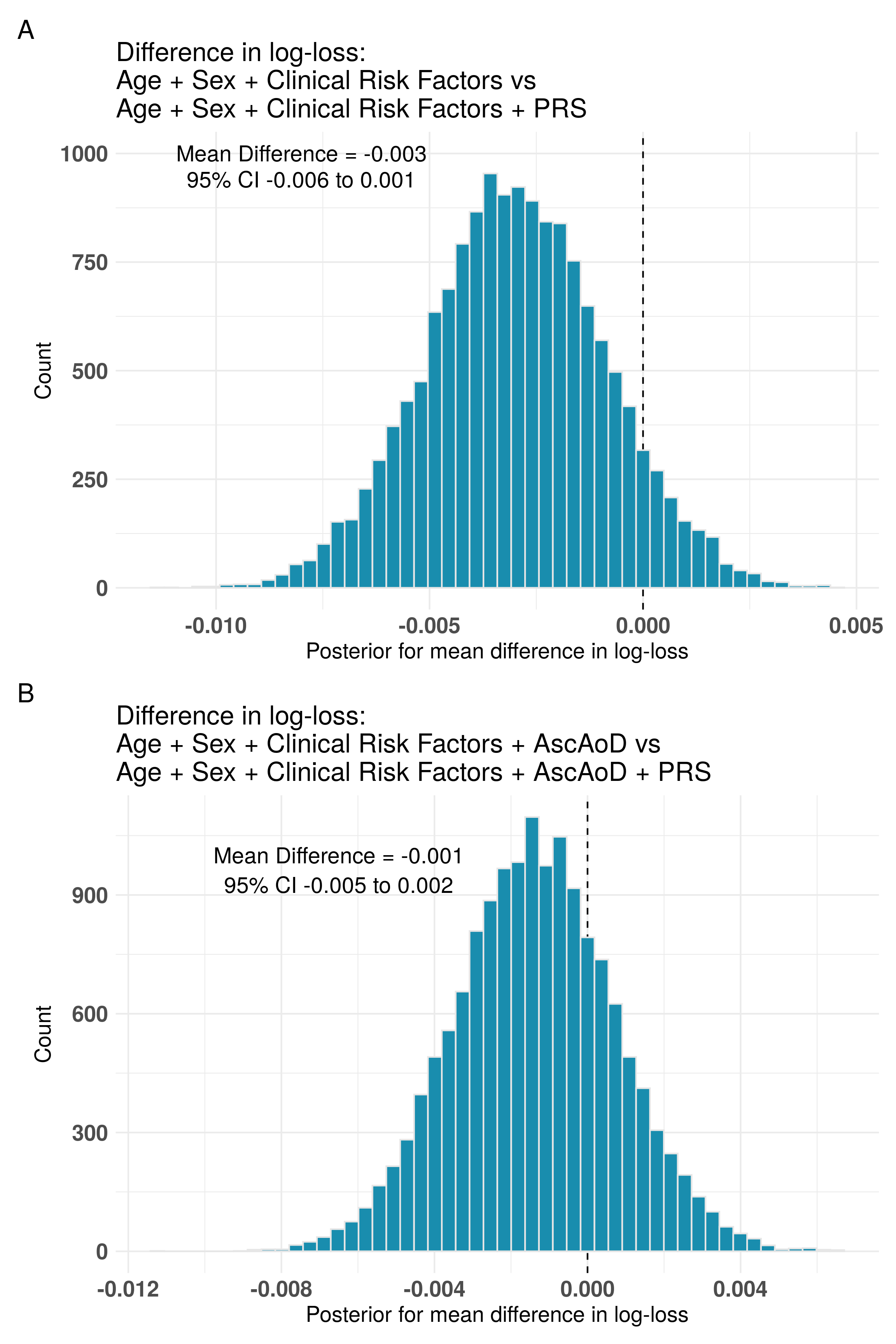

**Supplemental Figure 16: Cross-model Bayesian comparison of difference in log-loss between models with and without the inclusion of the Dissection-PRS among individuals genetically similar to the 1000G AFR reference population in PMBB.** Posterior distribution for the difference in log-loss between (A) Age + Sex + Clinical Risk Factors and Age + Sex + Clinical Risk Factors + Dissection-PRS; and (B) Age + Sex + Clinical Risk Factors + AscAoD and Age + Sex + Clinical Risk Factors + AscAoD + Dissection-PRS. Between-model mean difference and 95% credible intervals in each plot. AscAoD = ascending thoracic aortic diameter; PRS = thoracic aortic dissection polygenic risk score; CI = credible interval.

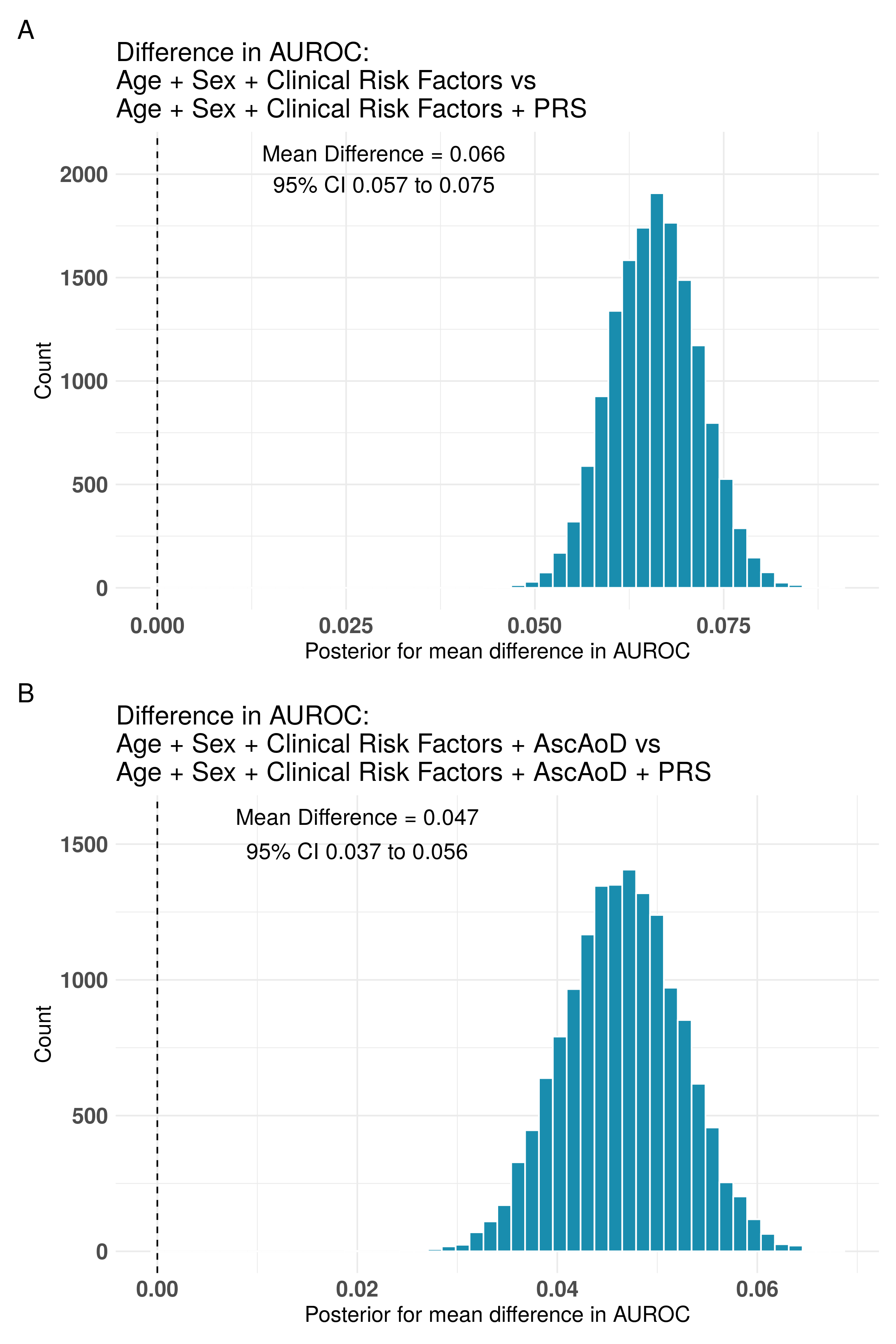

**Supplemental Figure 17: Cross-model Bayesian comparison of difference in area under the receiver operator characteristic curve (AUROC) between models with and without the inclusion of the Dissection-PRS among individuals in PMBB.** Cross-validated posterior distribution for the difference in AUROC between (A) Age + Sex + Clinical Risk Factors and Age + Sex + Clinical Risk Factors + Dissection-PRS; and (B) Age + Sex + Clinical Risk Factors + AscAoD and Age + Sex + Clinical Risk Factors + AscAoD + Dissection-PRS. Between-model mean difference and 95% credible intervals in each plot. AscAoD = ascending thoracic aortic diameter; AUROC = Area under the receiver operator characteristic curve; PRS = Thoracic aortic dissection polygenic risk score; CI = credible interval.

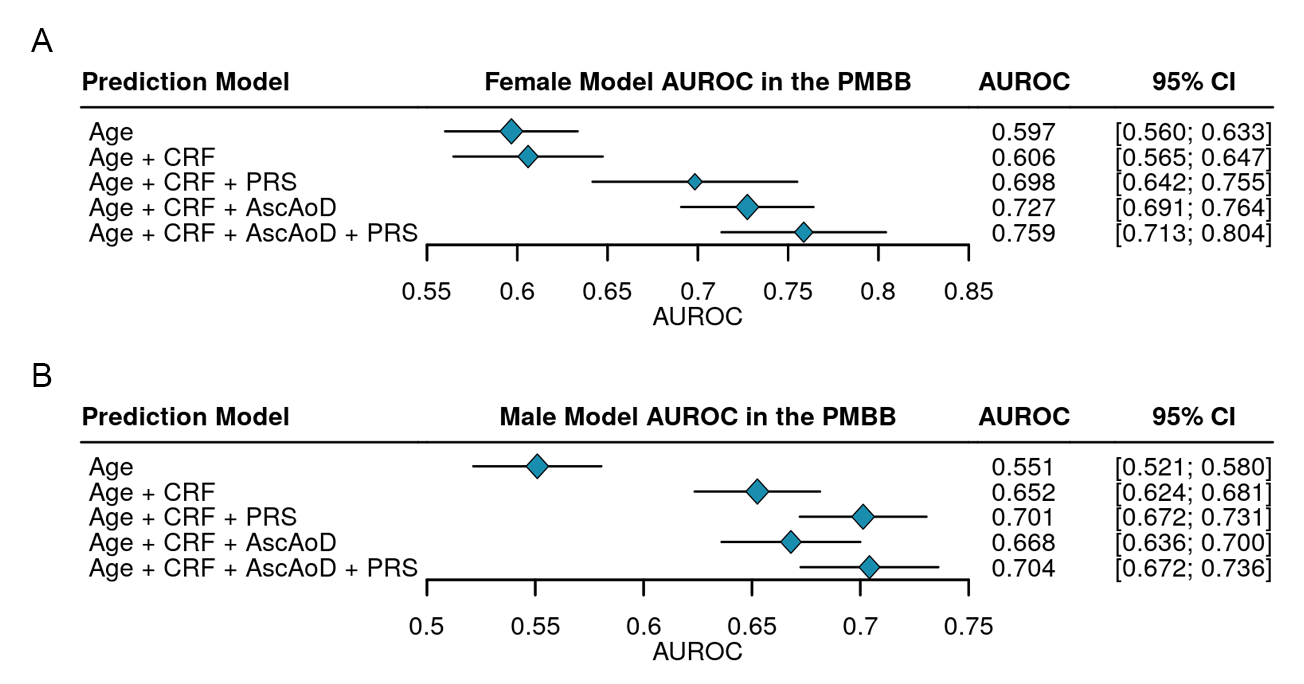

**Supplemental Figure 18: Area under the receiver operator curve comparison of logistic regression models to predict thoracic aortic dissection stratified by sex.** For both the PMBB female cohort (A) and the male cohort (B), five-times 10-fold cross-validation analysis of model AUROC for each of Age, Age + Clinical Risk Factors, Age + Clinical Risk Factors + Dissection-PRS, Age + Clinical Risk Factors + AscAoD, and Age + Clinical Risk Factors + AscAoD + Dissection-PRS. Corresponding error bars demonstrating 95% confidence intervals for models utilized among all individuals with AscAoD measured by TTE. AscAoD = ascending thoracic aortic diameter; CI = 95% confidence interval; CRF = Clinical risk factors; PRS = thoracic aortic dissection polygenic risk score.

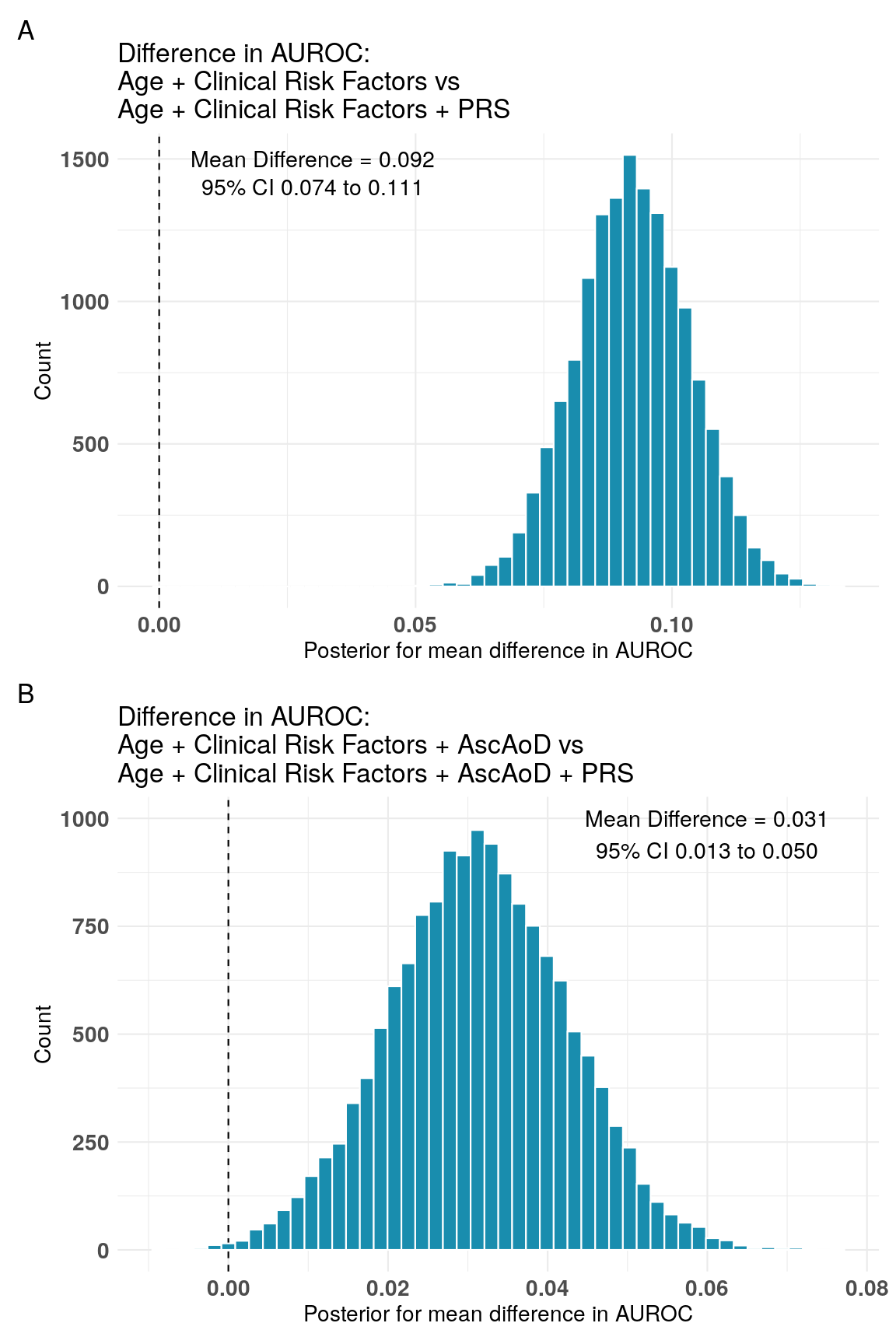

**Supplemental Figure 19: Cross-model Bayesian comparison of difference in area under the receiver operator characteristic curve between models with and without the inclusion of the Dissection-PRS among females in the PMBB.** Posterior distribution for the difference in log-loss between (A) Age + Clinical Risk Factors and Age + Clinical Risk Factors + Dissection-PRS; and (B) Age + Clinical Risk Factors + AscAoD and Age + Clinical Risk Factors + AscAoD + Dissection-PRS. Between-model mean difference and 95% credible intervals in each plot. AscAoD = ascending thoracic aortic diameter; PRS = thoracic aortic dissection polygenic risk score; CI = credible interval.

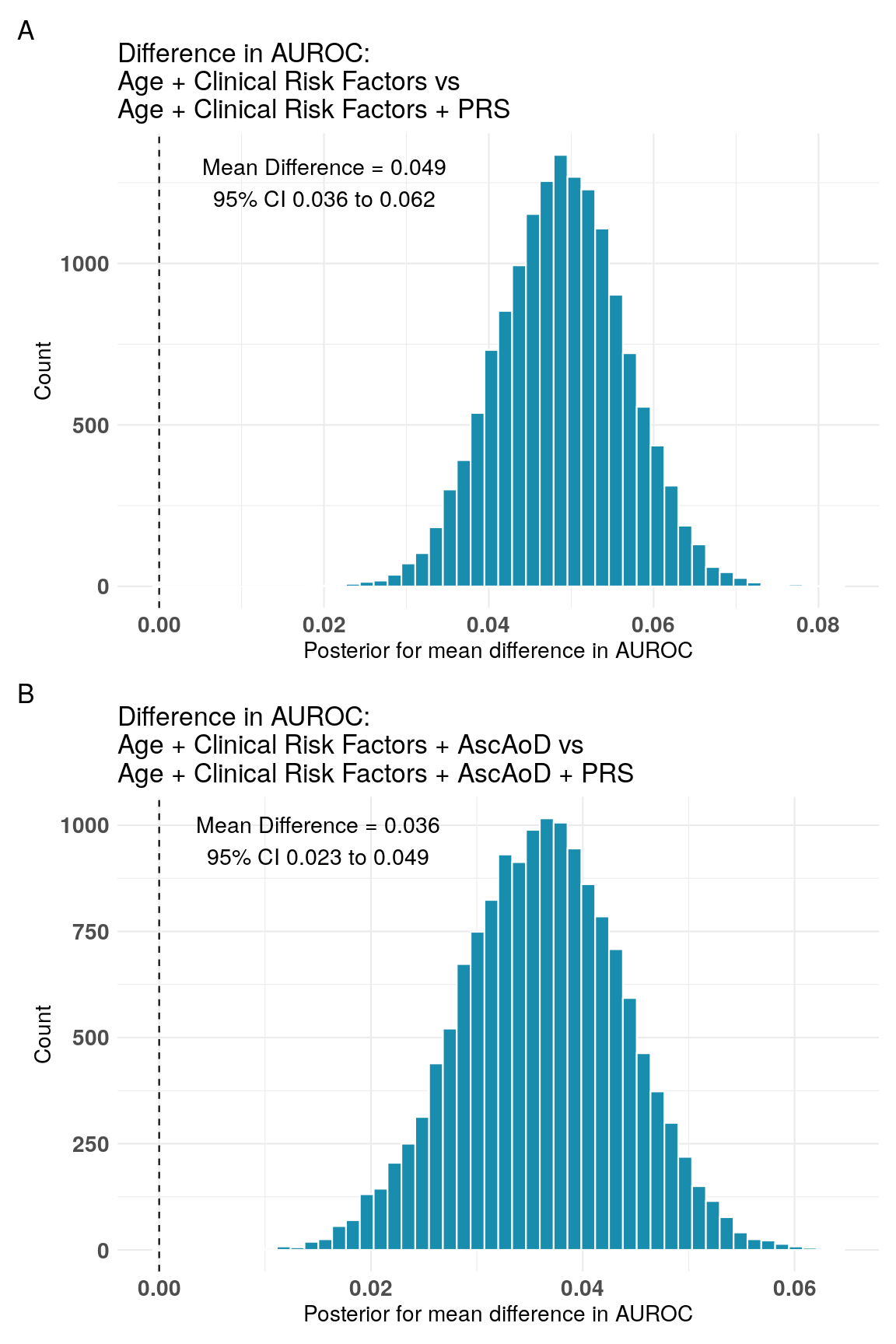

**Supplemental Figure 20: Cross-model Bayesian comparison of difference in area under the receiver operator characteristic curve between models with and without the inclusion of the Dissection-PRS among males in the PMBB.** Posterior distribution for the difference in log-loss between (A) Age + Clinical Risk Factors and Age + Clinical Risk Factors + Dissection-PRS; and (B) Age + Clinical Risk Factors + AscAoD and Age + Clinical Risk Factors + AscAoD + Dissection-PRS. Between-model mean difference and 95% credible intervals in each plot. AscAoD = ascending thoracic aortic diameter; PRS = thoracic aortic dissection polygenic risk score; CI = credible interval.

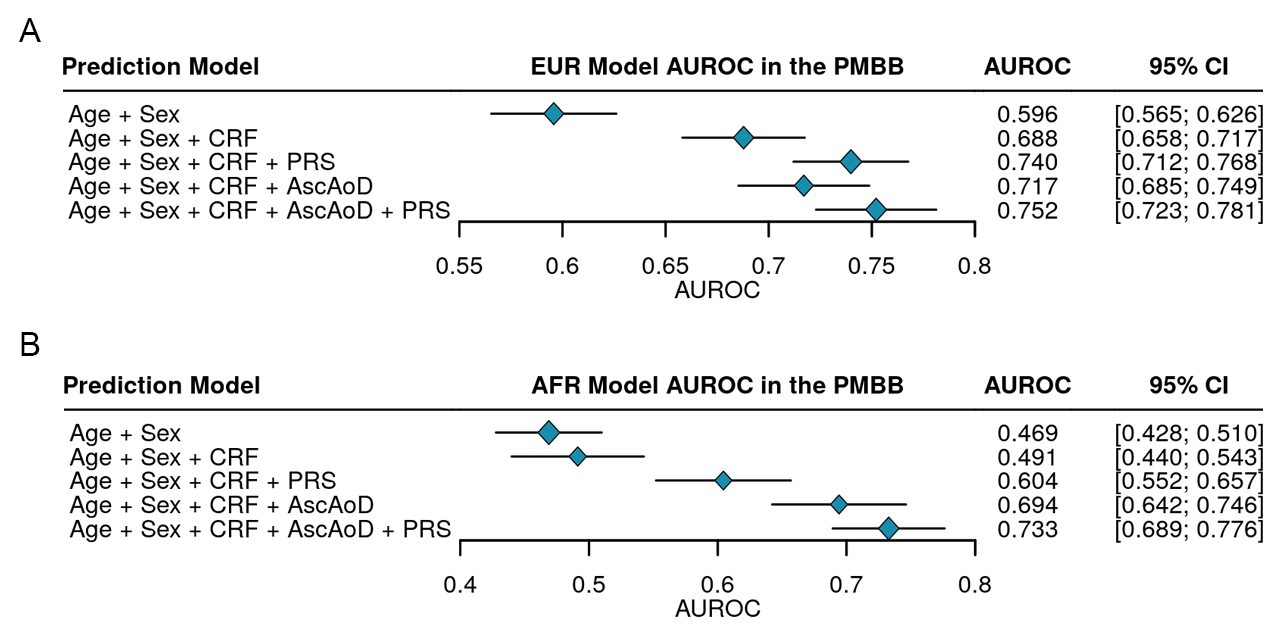
**Supplemental Figure 21: Area under the receiver operator curve comparison of logistic regression models to predict thoracic aortic dissection stratified by population group.** For both the PMBB EUR cohort (A) and the AFR cohort (B), five-times 10-fold cross-validation analysis of model AUROC for each of Age + Sex, Age + Sex + Clinical Risk Factors, Age + Sex + Clinical Risk Factors + Dissection-PRS, Age + Sex + Clinical Risk Factors + AscAoD, and Age + Sex + Clinical Risk Factors + AscAoD + Dissection-PRS. Corresponding error bars demonstrating 95% confidence intervals for models utilized among all individuals with AscAoD measured by TTE. AscAoD = ascending thoracic aortic diameter; CI = 95% confidence interval; CRF = Clinical risk factors; PRS = thoracic aortic dissection polygenic risk score.

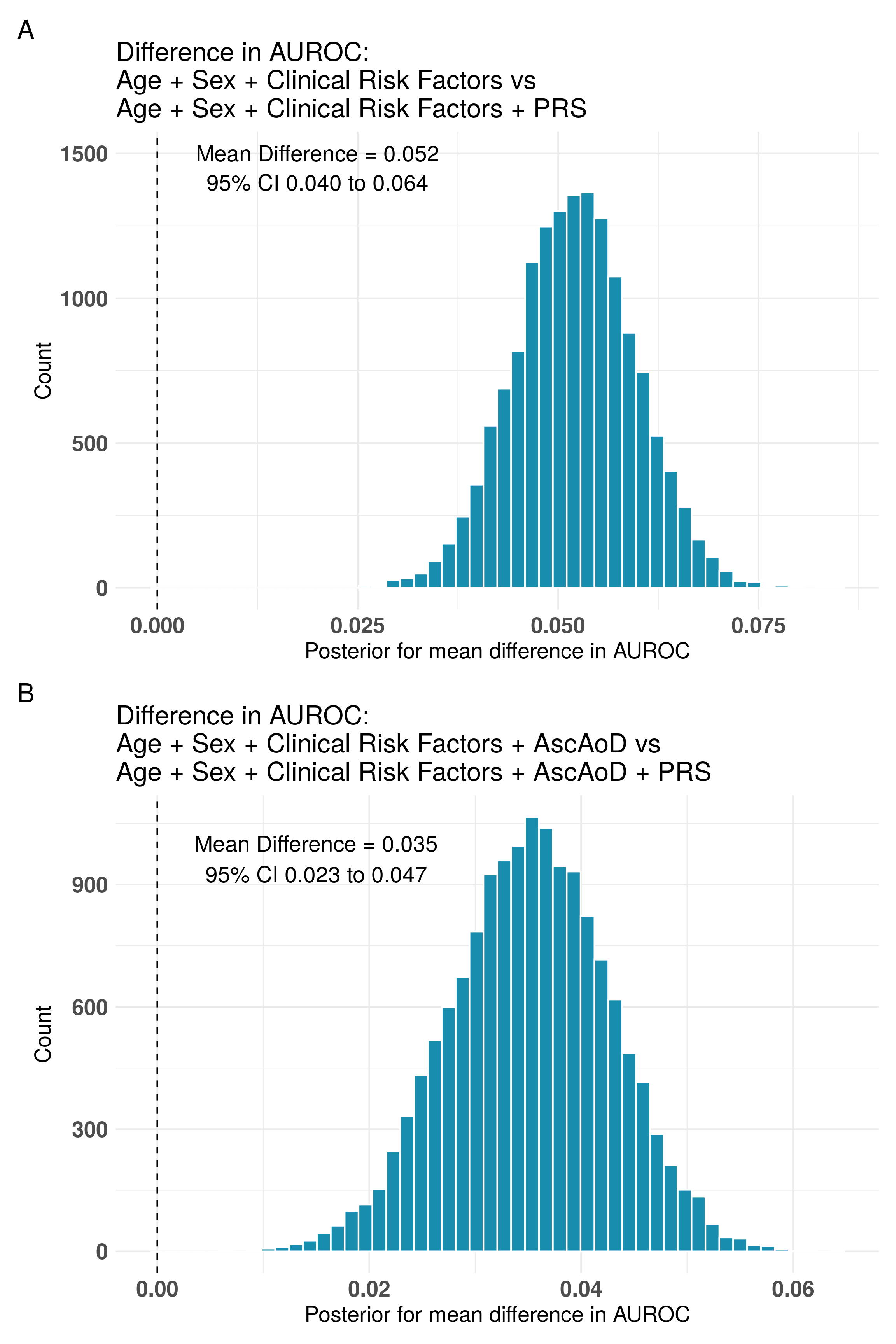

**Supplemental Figure 22: Cross-model Bayesian comparison of difference in area under the receiver operator characteristic curve between models with and without the inclusion of the Dissection-PRS among individuals genetically similar to the 1000G EUR reference population in the PMBB.** Posterior distribution for the difference in log-loss between (A) Age + Sex + Clinical Risk Factors and Age + Sex + Clinical Risk Factors + Dissection-PRS; and (B) Age + Sex + Clinical Risk Factors + AscAoD and Age + Sex + Clinical Risk Factors + AscAoD + Dissection-PRS. Between-model mean difference and 95% credible intervals in each plot. AscAoD = ascending thoracic aortic diameter; PRS = thoracic aortic dissection polygenic risk score; CI = credible interval.

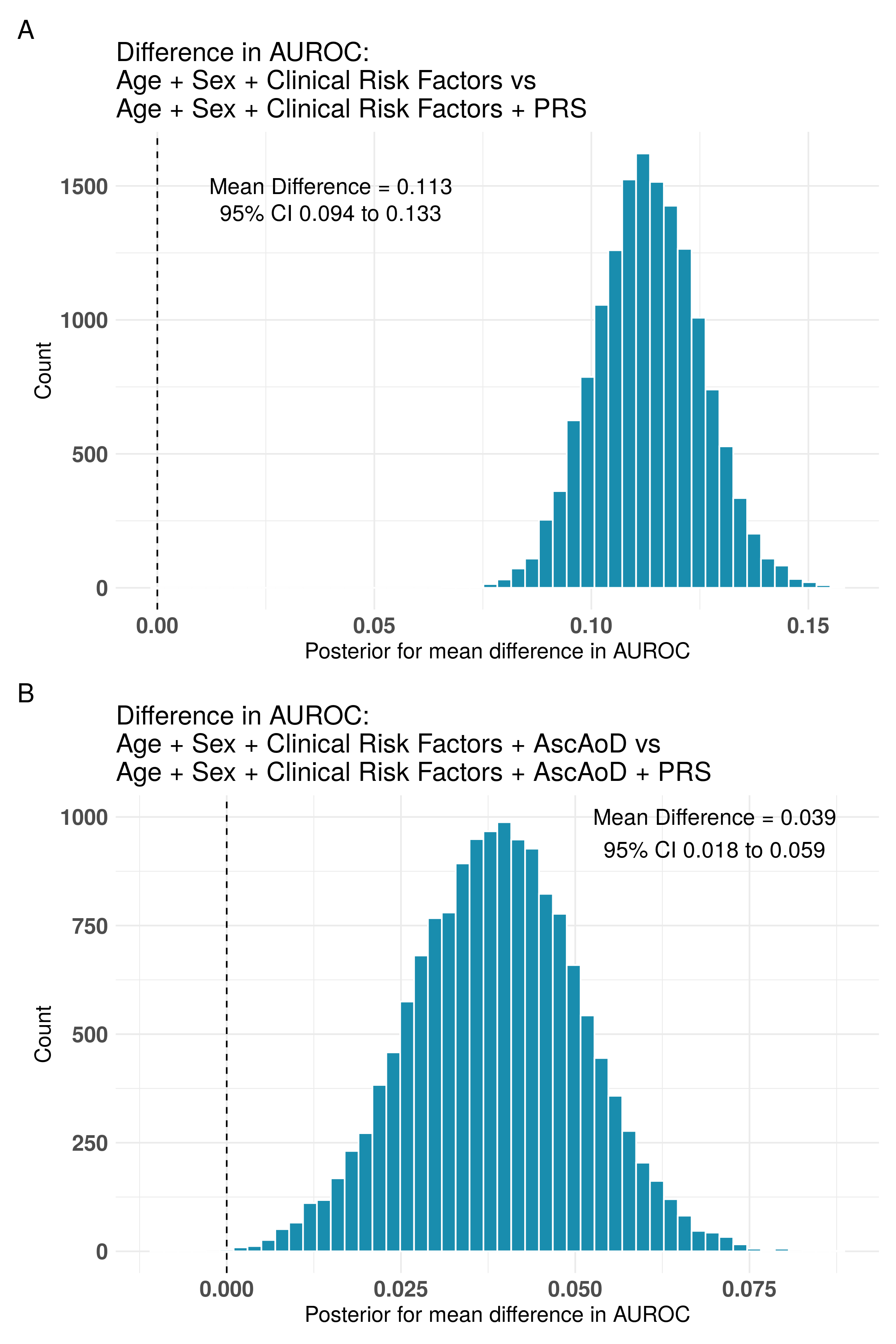

**Supplemental Figure 23: Cross-model Bayesian comparison of difference in area under the receiver operator characteristic curve between models with and without the inclusion of the Dissection-PRS among individuals genetically similar to the 1000G AFR reference population in the PMBB.** Posterior distribution for the difference in log-loss between (A) Age + Sex + Clinical Risk Factors and Age + Sex + Clinical Risk Factors + Dissection-PRS; and (B) Age + Sex + Clinical Risk Factors + AscAoD and Age + Sex + Clinical Risk Factors + AscAoD + Dissection-PRS. Between-model mean difference and 95% credible intervals in each plot. AscAoD = ascending thoracic aortic diameter; PRS = thoracic aortic dissection polygenic risk score; CI = credible interval.

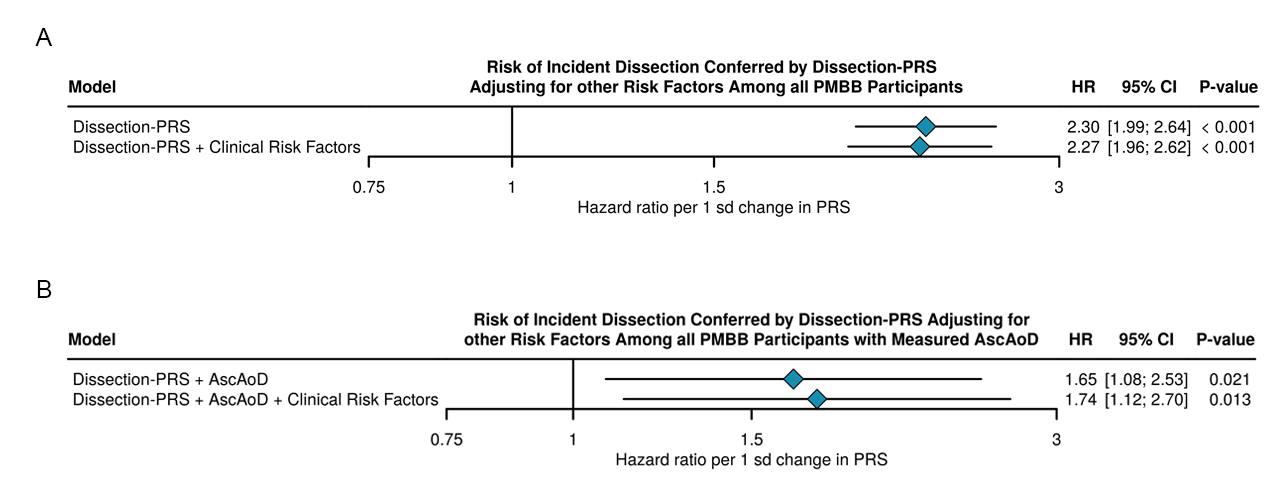

**Supplemental Figure 24: Cox proportional hazard ratio analysis of the association between the Dissection-PRS and incident dissection in different multivariable models.** Cox proportional hazard analysis of the effect of Dissection-PRS on incident dissection adjusting for (A) age + sex + genetic PCs 1-5 or age + sex + genetic PCs 1-5 + clinical risk factors among all PMBB individuals without prevalent dissection at the time or enrollment; or (B) age + sex + genetic PCs 1-5 + measured AscAoD or age + sex + genetic PCs 1-5 + measured AscAoD + clinical risk factors among all PMBB individuals without prevalent dissection at the time or enrollment and with at least one measurement of AscAoD prior to experiencing a dissection. AscAoD = Ascending aortic diameter measured by TTE; CI = confidence interval; HR = Hazard ratio; sd = standard deviation; Dissection-PRS = thoracic aortic dissection polygenic risk score.
